# Short-term and Mid-term Blood Pressure Variability and long-term Mortality: evidence from the Third National Health and Nutrition Examination Study

**DOI:** 10.1101/2023.12.18.23300161

**Authors:** David Steinsaltz, Hamish Patten, D W Bester, David H. Rehkopf

**Author notes:** Authors for communication: David Steinsaltz, Hamish Patten and David H. Rehkopf.

## Abstract

In considering the impact of blood-pressure variability on health outcomes, two methodological challenges arise: The presence of multiple timescales of variability that may act independently and interactively, and the fairly large stochastic uncertainty that is inevitable in estimates of individual variability based on modest numbers of observations. Here we present an application of Bayesian hierarchical modeling to the problem of estimating the effect of blood pressure (BP) variability on all-cause and cardiovascular mortality with two timescales – short-term variation among multiple measures at one visit, and medium-term variation between the measures at two visits several months apart. We use data from the Third National Health and Nutrition Examination Survey linked with up to 27 years of mortality follow-up. We find that medium-term systolic BP variability had a very significant predictive value for CV and all-cause mortality, around one-third as large as the well-established impact of mean systolic BP. Medium-term diastolic variability had an additional, though smaller, predictive effect. Short-term variability, on the other hand, had little or no measurable predictive value. The medium-term variability effect persisted when controlling for Framingham risk score.

## Introduction

While the association between hypertension and mortality risk was first noted in actuarial studies in the first half of the twentieth century(1,2) it was not until the 1960s that studies such as Framingham and the Seven Countries Study(3,4) demonstrated the link between hypertension and CVD risk. These observational studies in turn led to trials that showed treatment of hypertension could reduce risk.(5) In the United States, guidelines on the management of hypertension have been produced by the Joint National Commission since 1976 and are now in their eighth edition.(6) Screening for hypertension among adults for the prevention of cardiovascular disease is the only action with the highest “A” level evidence based rating from the U.S. Preventative Services Task Force (USPSTF). Despite this, the USPSTF notes that there is still a substantial research gap in understanding “white coat hypertension” and “masked hypertension”, which describe the variability of measured blood pressure depending on context. A better understanding of the clinical course associated with these variations is important not only for understanding errors in our current attributions of blood pressure to long-term health outcomes, but also potentially for identifying new mechanisms by which blood pressure may impact cardiovascular outcomes.

While most studies to date of blood pressure and cardiovascular risk have focused on mean blood pressure, there is a growing literature investigating the impacts of variation in blood pressure on cardiovascular-related outcomes.(7,8) This literature can be divided into studies that examine beat to beat variability, very short term (within an hour), short term (within 24 hours), mid-term or visit-to-visit (day to day), long term (< 5 years) and very-long term (>= 5 years) blood pressure variability(9). Work is still emerging on the clinical significance of these different types of blood pressure variation as they related to cardiovascular disease mortality. While these time windows of variation are sometimes framed as errors in measurement(10), emerging evidence suggests that visit-to-visit and 24-hour variation may both predict cardiovascular outcomes. Prior work showed that the standard deviation of visit-to-visit systolic blood pressure was associated with all-cause mortality over a 14-year follow-up.(11) Frattola and colleagues found that increased 24-hour blood pressure variability was related to end-organ damage in 73 hypertensive subjects followed for an average of 7.4 years.(12) Findings relating 24-hour or awake blood pressure variability to end-organ damage or the risk of cardiovascular events have been found in samples of hypertensive subjects(13), older individuals(14,15), and in the general population.(16) Visit-to-visit systolic blood pressure variability has likewise been associated with risk of stroke.(17,18) Importantly, visit-to-visit variability is likely to be systematically different between individuals, rather than reflecting merely random variation.(11)

These studies have focused primarily on blood pressure variability measured beyond the course of a single office visit. For incorporation of blood pressure variability into routine practice, it may also be useful to measure variation in the course of a patient’s visit for incorporation into risk stratification. However, the significance of the very short-term (VST) (measurements taken within an hour) variations in blood pressure that might be recorded in one visit is unclear. Previous work by Muntner and colleagues using a quantile analysis did not find associations between within visit systolic and diastolic blood pressure variability and mortality.(19) In this study, we consider the prognostic significance of the very short-term variations in blood pressure using the same large, U.S. nationally representative cohort of adults – but using a different analytic approach that better accounts for the randomness of individual variation to deal with regression dilution bias. In addition, we examine cardiovascular mortality specifically, and include a longer follow-up for mortality. We also take advantage of the availability of two sets of measurements (home and clinic) several months apart to estimate the effect of visit-to-visit variability, and explicitly compare the contribution of each of these different types of variability to cardiovascular mortality.

A key difficulty in using variability measures as covariates is that it takes a large number of measurements to accurately measure variability. Regression based on a small number of measurements per individual will be subject to bias in estimation of regression parameters.

While corrections for this “regression dilution” are possible, they make it difficult to combine data from individuals with different numbers of measurements, or multiple predictors that are each measured with uncertainty. Thus, while Rothwell et al. (2010) carefully separated out individuals with different numbers of blood pressure observations in their analysis, they produced multiple inconsistent parameter estimates. Here we apply a novel Bayesian methodology that correctly accounts for measurement error, and is fully flexible with regard to the different covariates that can be combined.

## METHODS

### Study population

Data were from the Third National Health and Nutrition Examination Survey (NHANES III), a survey and examination of a sample of the civilian, non-institutionalized US population conducted by the National Center for Health Statistics from 1988-1994.(20) We chose to focus on this older version of NHANES in order to have baseline blood pressure measurements with sufficient follow-up time to examine longer-term impacts on mortality. We used data from the National Death Index (NDI) mortality linkage through December of 2015.

### Measurement of blood pressure

The examinations included an in-home examination and a mobile clinic examination. The in-home measures were taken by trained interviewers, and the mobile clinic measures were taken by physicians who were specifically trained for measurement practices for the survey. In the home examination portion of this survey, blood pressure was measured antecubitally three times for each subject, with one minute between recordings.(21) A second set of three measurements was taken at the mobile examination clinic, also with one minute between recordings. There were generally a few months between these sets of measurements. Physicians taking the measurements were instructed that subjects should sit still for a minimum of 5 minutes prior to the measurement being taken. All BP measurements were recorded to the nearest even number. There were inconsistencies in the home data collection that became apparent on examination of the reported BP measurements, several of which to our knowledge have not been reported in previous analyses of the NHANES III data. These were: 1) Preference for 0 and 8 as final digits 2) Dependent repetition, and 3) Missing or implausible measurements (a preference specifically for 0 end digit in other NHANES waves was previously reported).(22,23) Details of how these were accounted for within the statistical procedure – as well as by exclusion of questionable or unusable data – are in sections 1.3 and 2.2 of the appendices. We also excluded the small group of subjects with ethnicity “Other”, as we wished to have robust estimation of baseline mortality within each ethnic group. This left 14654 individuals with two complete sets of usable BP measurements who were included in our primary analysis.

### Measurement of other variables

Our primary focus was to determine whether variability of BP provides additional information for predicting future mortality beyond what is already embedded in standard measures of risk. As the standard measure of risk, we use the 1998 version of the Framingham Risk Score (FRS). Out of the usable NHANES population 5063 were excluded from FRS calculation because their age was below 30 or above 74. An additional 583 had no FRS score because of missing cholesterol values or pre-existing CHD, leaving 9008 individuals in what we refer to as the “FRS population”. While we focus analyses on this healthier middle-aged population, we also include findings on the general population.

### Statistical models

The data allow us to examine two versions of very short term (VST) BP variability – in the home and in the mobile examination clinic. We could also estimate a measure of longer term (LT) variability by the difference between the two sets of measurements. We denote the three variability parameters for each individual by

*σ_C,i_=Clinic standard deviation*
*σ_H,i_=Home standard deviation*
*Δ_i_=(μ||C, i−μ_H,i_)/ 2,*
*M_i_=(μ||C, i+ μ_H,i_)/ 2*
*where μ_C,i_*=*Clinic mean; μ_H,i_*=*Home mean*.

with each of these parameters calculated for systolic and diastolic BP measurements.

In our model we assume that each individual has an overall BP mean *M_i_* and a Clinic— Home difference *Δ_i_*. These are assumed normally distributed in the population and independent. There are also individual home and clinic variance covariates (SD^2^), which are assumed to have independent inverse gamma distributions, consistent with the observation that the empirical SDs of the three clinic and three home measurements have relatively small Pearson correlations, the largest being r=0.269 between clinic systolic SD and mean. The overall means and the absolute differences between the means also have manageably small correlations, the largest, the only minor exceptions being r=0.347, between overall mean systolic BP and |Δ| for diastolic BP. Details about the empirical BP distribution are discussed in section 1.2 of Appendix A.

The main method we apply is a Bayesian hierarchical proportional hazards model, stratified by gender and race-ethnicity. This approach has the advantage of not suffering from regression-dilution bias, which occurs when there are random errors in the covariate of interest. This noisy-covariate problem is particularly acute when the covariate is a measure of variability based on a small number of observations. The model-fitting has been carried out using Markov Chain Monte Carlo (MCMC) on the Stan platform(24), whereby the exact algorithm used was the No-U-Turn Sampler (NUTS) Hamiltonian Monte Carlo (HMC). A generally similar method was originally proposed for BP variability in NHANES in (25), and has since been applied elsewhere to modeling similar data.(26)

As the statistical challenge is to estimate the effect of BP variability on long-term survival while accounting appropriately for covariate uncertainty — variability is only crudely ascertained from such a small number of measurements — it is most important to have a roughly accurate but stable estimate of the uncertainty in individual BP variance. This inspires our empirical Bayes approach(27), where we first calculate a maximum likelihood estimate of the hyperparameters that define the distribution of individual BP parameters (means and variances) and then hold these fixed as a prior distribution for Bayesian analysis of the survival data.

We define *Yij*, where *i*=1,…,N for the number of individuals and *j*=1,2,3, for the number of BP measurements for subject *i*. Then *Y _ij_*=*μ_i_* +*σ _i_ ε_ij_*, where *μ_i_* and *σ _i_* represent the mean and standard deviation respectively, for the *i*-th individual, and *ε _ij_* are independent standard normal random variables. Blood pressure interacts with mortality as in the standard Cox proportional hazards model, so that individual *i* has mortality rate *μ_i_* (*t*) at age *t* given by

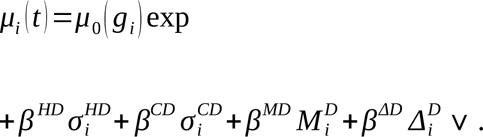

or, in the case of applying the FRS-1998 score

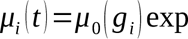

Here the superscripts H and C label *Home* or *Clinic* BP measurements, S and D tell whether these are systolic or diastolic BP, and *g_i_*is the demographic group – one of six possible combinations of sex (male/female) and race-ethnicity (White/Black/Mexican-American – these being the racial-ethnic categories available in the NHANES data, except for a small “Other” category that we have excluded from the analysis). For each demographic group we estimate a separate Gompertz baseline mortality *μ*_0_ ( *g*)=*B e^θg t^*. To better understand the value of these variability covariates in augmenting the predictive power of more traditional covariates, we also consider a version of the model with an extra term *β^FRS^ FRS*, where *FRS_i_* is the Framingham Risk Score (1998 version) for individual *i*. As the mean systolic BP is a substantial component of the FRS, this version of the model drops the mean-BP terms 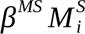 and 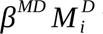 from the linear predictor. As FRS scores could be computed for only 9008 subjects, we performed the original analysis on this subpopulation as well, to distinguish differences due to the changed model from those due to the changed population.

It is important to recognize that the individual covariates – 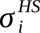, etc. – that enter into the individual mortality rate above are not observed quantities. They influence the observed BP measurements, and in the course of our MCMC simulations posterior distributions are generated for each individual, representing an inference about the range of possible values that these unknown quantities might have, based on the totality of observations. In order to improve the numerical stability and interpretability of the results, these inferred covariates are centered and normalized. Each inferred covariate *x_i_* appears in the mortality formula as 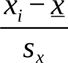 where *s_x_* is the posterior standard deviation for the covariate *x* and *<u>x</u>* is a centering parameter chosen to bring the average model mortality over the population – and averaged over the complete posterior distribution – as near as possible to the observed mortality at each age.

Our statistical approach proceeded through the following steps (further details are provided in Appendix B):

1. Estimate the population distribution of meanBP (M), medium-term BP variability (Δ, the absolute difference between home and clinic means), and very short-term BP variability ( *σ _H_* and *σ _C_*, the home and clinic SD respectively), separately for systolic and diastolic BP. This is an empirical Bayes approach to determining the highest level prior distribution. Fixing these at the outset substantially accelerates the convergence of the algorithm, relative to a completely hierarchical approach that would be updating the top-level parameters at every round.
2. Treat each individual’s inherent value of these 8 quantities as unknown samples from the population distribution, modified by the evidence of their 6 BP measurements.
3. In the framework of a Markov-chain Monte Carlo (MCMC) algorithm these unknown quantities are multiply imputed, to infer parameter values (8 different β parameters, corresponding to the 8 covariates, and 12 Gompertz mortality parameters, two for each of two sexes and three ethnic groups) that take account of this individual-level uncertainty.
4. The resulting posterior parameters are rescaled to the SD of the posterior samples and interpreted for their statistical significance and impact on mortality. Note that β represents the impact on mortality risk of changes in a latent covariate such as an individual’s SD for systolic BP, that is not exactly known from this data set, but which could in principle be measured to arbitrary precision, by increasing the number of BP measurements.

When presenting the outputs of our Bayesian model we adapt standard Bayesian terminology to something closer to the nomenclature customary in epidemiology. We give for each parameter estimate a central 90% credible interval, meaning that the parameter has a 5% chance each of being below or above the interval. If the interval excludes 0 that is meaningful evidence that the parameter is nonzero. The posterior probability that a given parameter is on the side of zero given by the alternative – for us, generally, the positive side – is the “Bayes factor”. It is conventional to say that strong evidence for the research hypothesis is provided by a Bayes factor >20 and decisive evidence by a Bayes factor >150.(28) We present instead a quantity more comparable to the usual one-tailed p-values, which is p=min(1,BF)/(1+BF), which may be understood as an estimate of the probability that the parameter is not on the side of the median estimate; thus P<0.10 (BF>19) may be seen as significant evidence that the covariate demonstrates a real effect, and P<0.013 (BF>150) as strong evidence. Results below 0.001 are reported simply as <0.001, because of the limits of precision in these simulation-based computations.

Each covariate is normalized by division by the observed posterior SD for its corresponding covariate. This makes all the coefficients comparable in scale. Thus, *β*=0.18, for example, always means that an increase of 1 SD in an individual’s value of this covariate corresponds to a 20% increase in mortality risk (relative to the average for the same age, sex, and racial-ethnic group).

## RESULTS

Table 1 presents the results of the association between mean, inter-visit precision, home precision and clinic precision for both systolic and diastolic BP with cardiovascular mortality and all-cause mortality in the subset of the NHANES population with the 1998 Framingham Risk Scores. We first describe the findings for cardiovascular mortality, our primary outcome of interest. Both systolic mean BP (M) and systolic inter-visit difference (|Δ^S^|) had highly significant effects, with the coefficient for the mean being about three times as large. No other coefficient was found to be relevant to a high degree of statistical confidence, though diastolic inter-visit difference (|Δ^D^|) had a comparable-sized coefficient estimate, and a Bayesian p-value 0.053. The mean (normalized) parameter estimate for systolic mean is 0.348, and for systolic Delta is 0.116. To appreciate the influence of these parameters, we note that the population mean of mean SBP is 125.9, and the SD is 18.3; thus individuals with mean SBP around 144 – approximately one-sixth of the survey population is above this level – would be expected to have 42% higher CV mortality than others of the same age, sex, and racial-ethnic category (risk ratio 1.42). The corresponding mortality increase for individuals whose intervisit SBP semidifference (|Δ|) is around 4.7 points above the population mean of 5.2 is 12%.

**Table 1.**
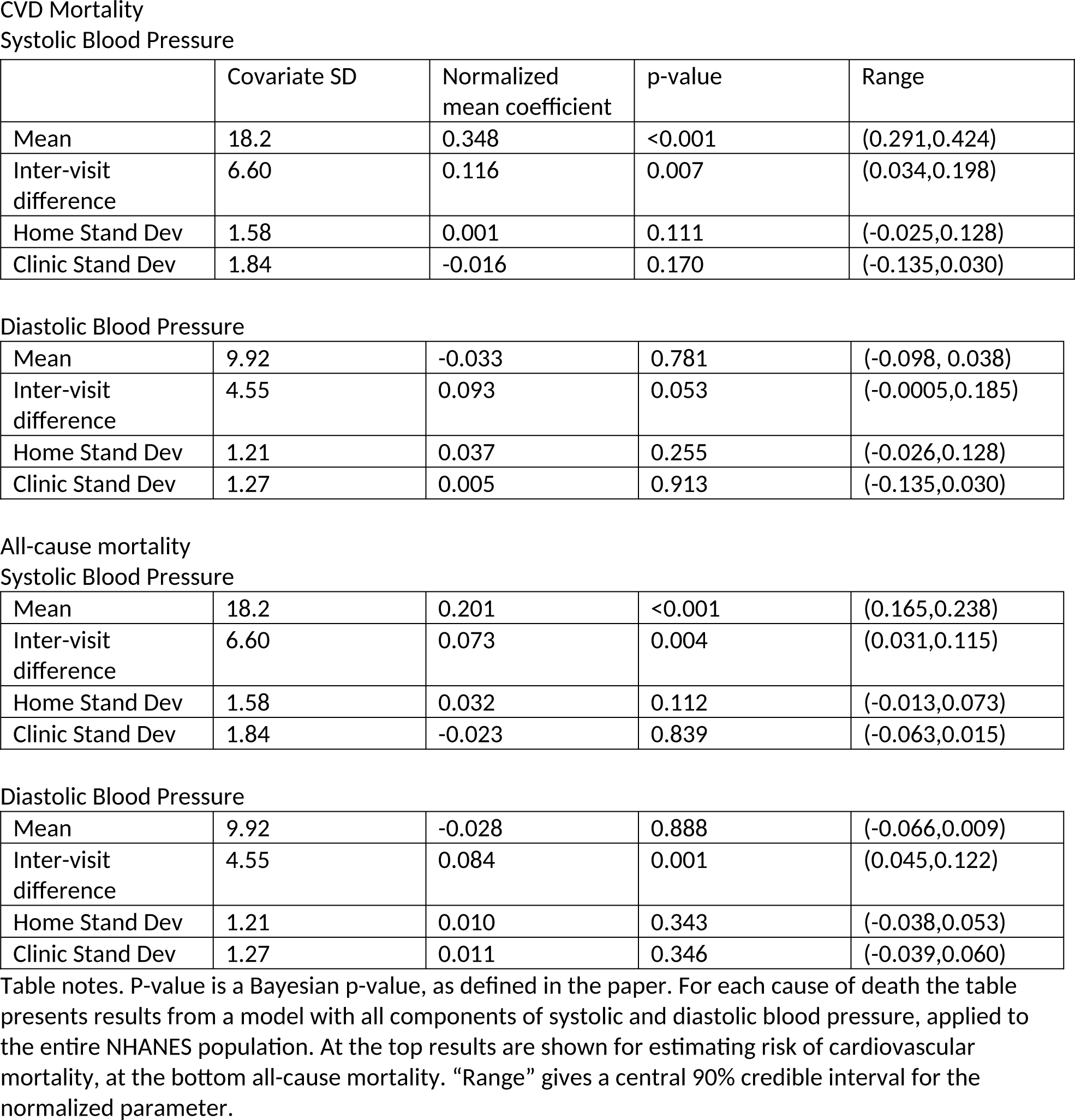
Parameter estimates for CVD and all-cause mortality, NHANES III, Framingham risk score population.

Systolic Blood Pressure
|  | Covariate SD | Normalized mean coefficient | p-value | Range |
| --- | --- | --- | --- | --- |
| Mean | 18.2 | 0.348 | <0.001 | (0.291,0.424) |
| Inter-visit difference | 6.60 | 0.116 | 0.007 | (0.034,0.198) |
| Home Stand Dev | 1.58 | 0.001 | 0.111 | (-0.025,0.128) |
| Clinic Stand Dev | 1.84 | -0.016 | 0.170 | (-0.135,0.030) |
| Mean | 9.92 | -0.033 | 0.781 | (-0.098, 0.038) |
| Inter-visit difference | 4.55 | 0.093 | 0.053 | (-0.0005,0.185) |
| Home Stand Dev | 1.21 | 0.037 | 0.255 | (-0.026,0.128) |
| Clinic Stand Dev | 1.27 | 0.005 | 0.913 | (-0.135,0.030) |
| Systolic Blood Pressure |  |  |  |  |
| Mean | 18.2 | 0.201 | <0.001 | (0.165,0.238) |
| Inter-visit difference | 6.60 | 0.073 | 0.004 | (0.031,0.115) |
| Home Stand Dev | 1.58 | 0.032 | 0.112 | (-0.013,0.073) |
| Clinic Stand Dev | 1.84 | -0.023 | 0.839 | (-0.063,0.015) |
| Mean | 9.92 | -0.028 | 0.888 | (-0.066,0.009) |
| Inter-visit difference | 4.55 | 0.084 | 0.001 | (0.045,0.122) |
| Home Stand Dev | 1.21 | 0.010 | 0.343 | (-0.038,0.053) |
| Clinic Stand Dev | 1.27 | 0.011 | 0.346 | (-0.039,0.060) |

Table notes. P-value is a Bayesian p-value, as defined in the paper. For each cause of death the table presents results from a model with all components of systolic and diastolic blood pressure, applied to the entire NHANES population. At the top results are shown for estimating risk of cardiovascular mortality, at the bottom all-cause mortality. "Range" gives a central 90% credible interval for the normalized parameter.

Applying the same model to all-cause mortality we find, unsurprisingly, that the coefficients that were statistically significant for predicting CV mortality have been reduced somewhat. In some cases, though, because of the increased power due to the much larger number of events forming the basis for the analysis, the statistical significance has increased. This is most notable for diastolic inter-visit difference, where the Bayesian p-value has gone down from 0.053 to 0.001.

When we look to the results for the whole population in Table 2 we see very similar coefficients, with an increase in significance for the systolic mean and Delta, and the diastolic Delta, due to the larger population size. There are two notable differences: an unexpected strongly negative coefficient for the effect of diastolic mean on cardiovascular and all-cause mortality, and a marginally significant negative influence of clinic diastolic SD on cardiovascular mortality. This could be seen as consistent with the known When mean BPs were removed from the model and replaced by FRS we obtained the results tabulated in Table 3. We see that the coefficients for systolic and diastolic Delta are both strongly positive, and larger than they were in the model that included only the BP means without the FRS. This shows that the inter-visit difference is giving information that is not included in the FRS. We also see in this model a marginally significant effect of short-term home-measure systolic BP variation. For predicting CV mortality the Bayesian p-value is 0.047, and for all-cause mortality it is 0.033. As this is an isolated non-null result among multiple hypothesis tests, it must be viewed with some skepticism, but it does suggest that more targeted exploration of a link between short-term SBP variability and mortality may be warranted.

**Table 2.**
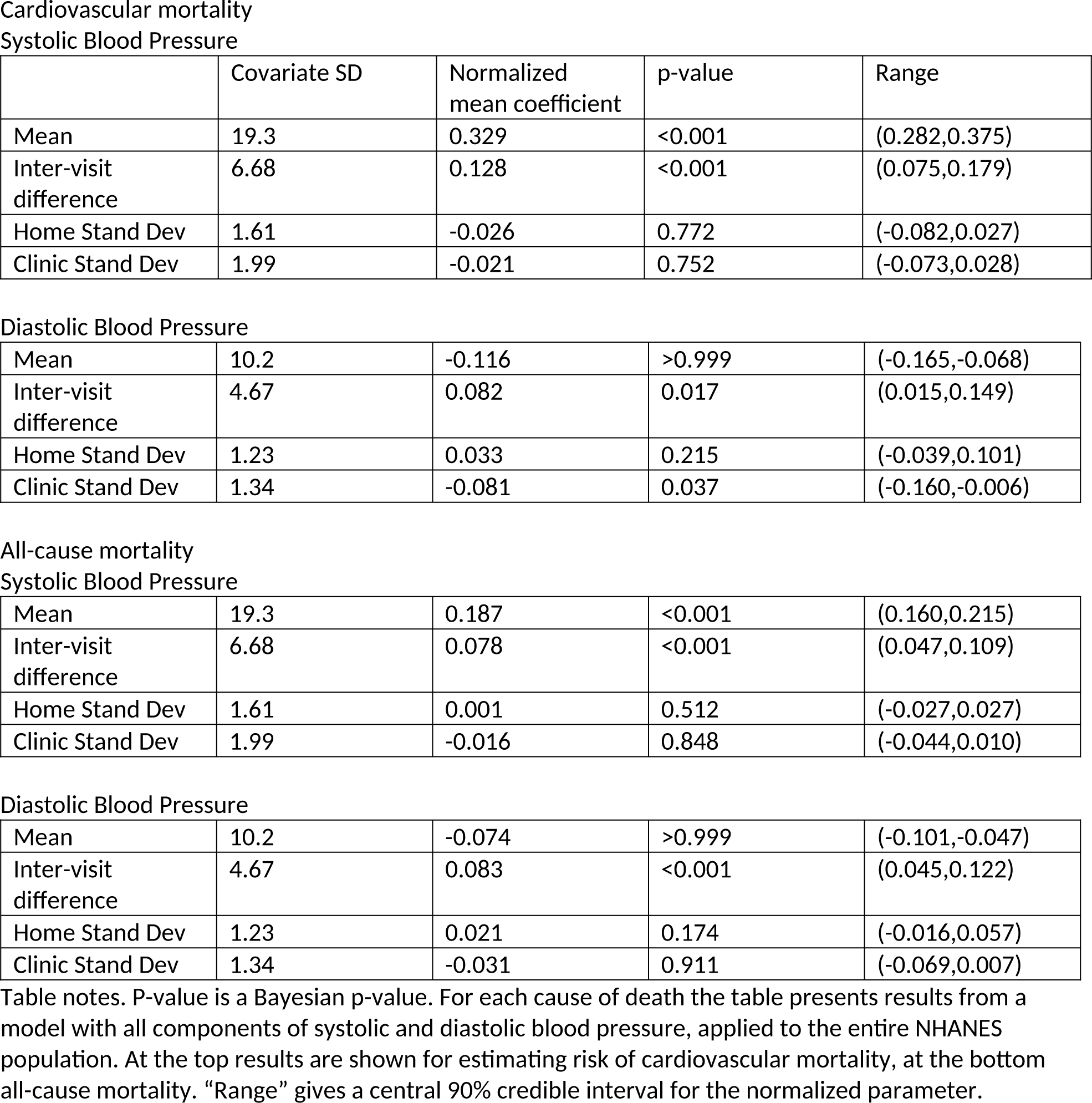
Parameter estimates for Cardiovascular and all-cause mortality, NHANES III, Full population.

Systolic Blood Pressure
|  | Covariate SD | Normalized mean coefficient | p-value | Range |
| --- | --- | --- | --- | --- |
| Mean | 19.3 | 0.329 | <0.001 | (0.282,0.375) |
| Inter-visit difference | 6.68 | 0.128 | <0.001 | (0.075,0.179) |
| Home Stand Dev | 1.61 | -0.026 | 0.772 | (-0.082,0.027) |
| Clinic Stand Dev | 1.99 | -0.021 | 0.752 | (-0.073,0.028) |
| Mean | 10.2 | -0.116 | >0.999 | (-0.165,-0.068) |
| Inter-visit difference | 4.67 | 0.082 | 0.017 | (0.015,0.149) |
| Home Stand Dev | 1.23 | 0.033 | 0.215 | (-0.039,0.101) |
| Clinic Stand Dev | 1.34 | -0.081 | 0.037 | (-0.160,-0.006) |
| Systolic Blood Pressure |  |  |  |  |
| Mean | 19.3 | 0.187 | <0.001 | (0.160,0.215) |
| Inter-visit difference | 6.68 | 0.078 | <0.001 | (0.047,0.109) |
| Home Stand Dev | 1.61 | 0.001 | 0.512 | (-0.027,0.027) |
| Clinic Stand Dev | 1.99 | -0.016 | 0.848 | (-0.044,0.010) |
| Mean | 10.2 | -0.074 | >0.999 | (-0.101,-0.047) |
| Inter-visit difference | 4.67 | 0.083 | <0.001 | (0.045,0.122) |
| Home Stand Dev | 1.23 | 0.021 | 0.174 | (-0.016,0.057) |
| Clinic Stand Dev | 1.34 | -0.031 | 0.911 | (-0.069,0.007) |

Table notes. P-value is a Bayesian p-value. For each cause of death the table presents results from a model with all components of systolic and diastolic blood pressure, applied to the entire NHANES population. At the top results are shown for estimating risk of cardiovascular mortality, at the bottom all-cause mortality. "Range" gives a central 90% credible interval for the normalized parameter.

**Table 3.**
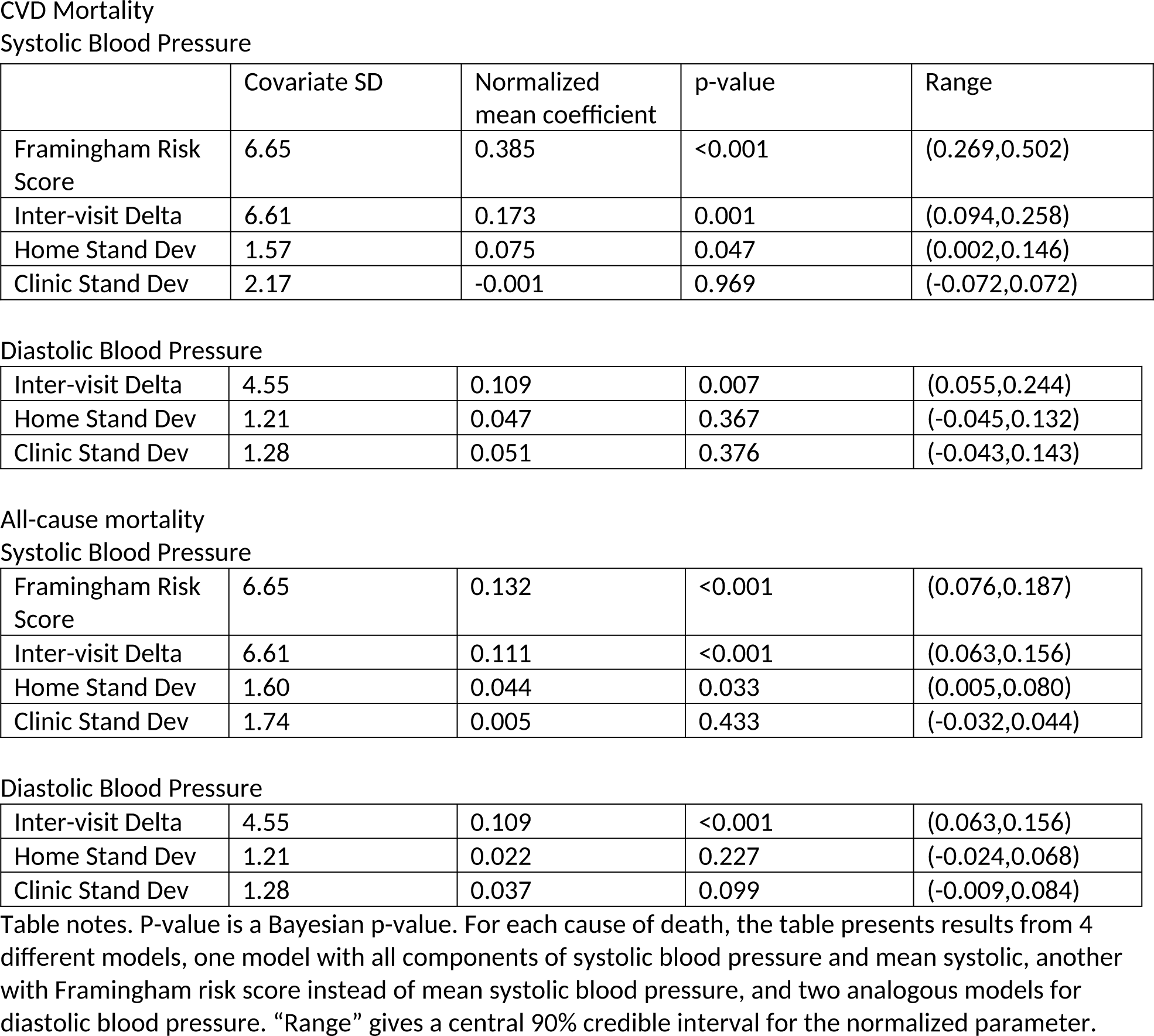
Parameter estimates for CVD and all-cause mortality, NHANES III, model including Framingham Risk Score.

|  | Covariate SD | Normalized mean coefficient | p-value | Range |
| --- | --- | --- | --- | --- |
| Framingham Risk Score | 6.65 | 0.385 | <0.001 | (0.269,0.502) |
| Inter-visit Delta | 6.61 | 0.173 | 0.001 | (0.094,0.258) |
| Home Stand Dev | 1.57 | 0.075 | 0.047 | (0.002,0.146) |
| Clinic Stand Dev | 2.17 | -0.001 | 0.969 | (-0.072,0.072) |
| Inter-visit Delta | 4.55 | 0.109 | 0.007 | (0.055,0.244) |
| Home Stand Dev | 1.21 | 0.047 | 0.367 | (-0.045,0.132) |
| Clinic Stand Dev | 1.28 | 0.051 | 0.376 | (-0.043,0.143) |
| Framingham Risk Score | 6.65 | 0.132 | <0.001 | (0.076,0.187) |
| Inter-visit Delta | 6.61 | 0.111 | <0.001 | (0.063,0.156) |
| Home Stand Dev | 1.60 | 0.044 | 0.033 | (0.005,0.080) |
| Clinic Stand Dev | 1.74 | 0.005 | 0.433 | (-0.032,0.044) |
| Inter-visit Delta | 4.55 | 0.109 | <0.001 | (0.063,0.156) |
| Home Stand Dev | 1.21 | 0.022 | 0.227 | (-0.024,0.068) |
| Clinic Stand Dev | 1.28 | 0.037 | 0.099 | (-0.009,0.084) |

Table notes. P-value is a Bayesian p-value. For each cause of death, the table presents results from 4 different models, one model with all components of systolic blood pressure and mean systolic, another with Framingham risk score instead of mean systolic blood pressure, and two analogous models for diastolic blood pressure. "Range" gives a central 90% credible interval for the normalized parameter.

Rather than considering a single target percentile, we may choose to look at the whole risk distribution implied by the model, by plotting receiver-operator characteristic (ROC) curves, shown in figure 1. Within the context of a proportional hazards model, the plot shows how concentrated relative risk is. Within a population of a fixed age, the model predicts deaths to arise in proportion to the relative risk, so this may be thought of as a plot of true positive rate against false positive rate.(30) The ROC curves measure the overall performance of the model to predict, based on the corresponding BP measurements, which individuals (of the same sex, race, and age) will be the next to die. (A perfect oracle would appear in this plot as a right-angled bracket, with area 1 and AUC=1, as there would be no false-positives). We see here that a linear predictor including the |Δ| and FRS does increase the AUC from 0.66 to 0.69 for all-cause mortality, and from 0.71 to 0.73 for CVD mortality. Further discussion of ROC curves for empirical mortality is in section 3.3 of Appendix C.

**Figure 1.**
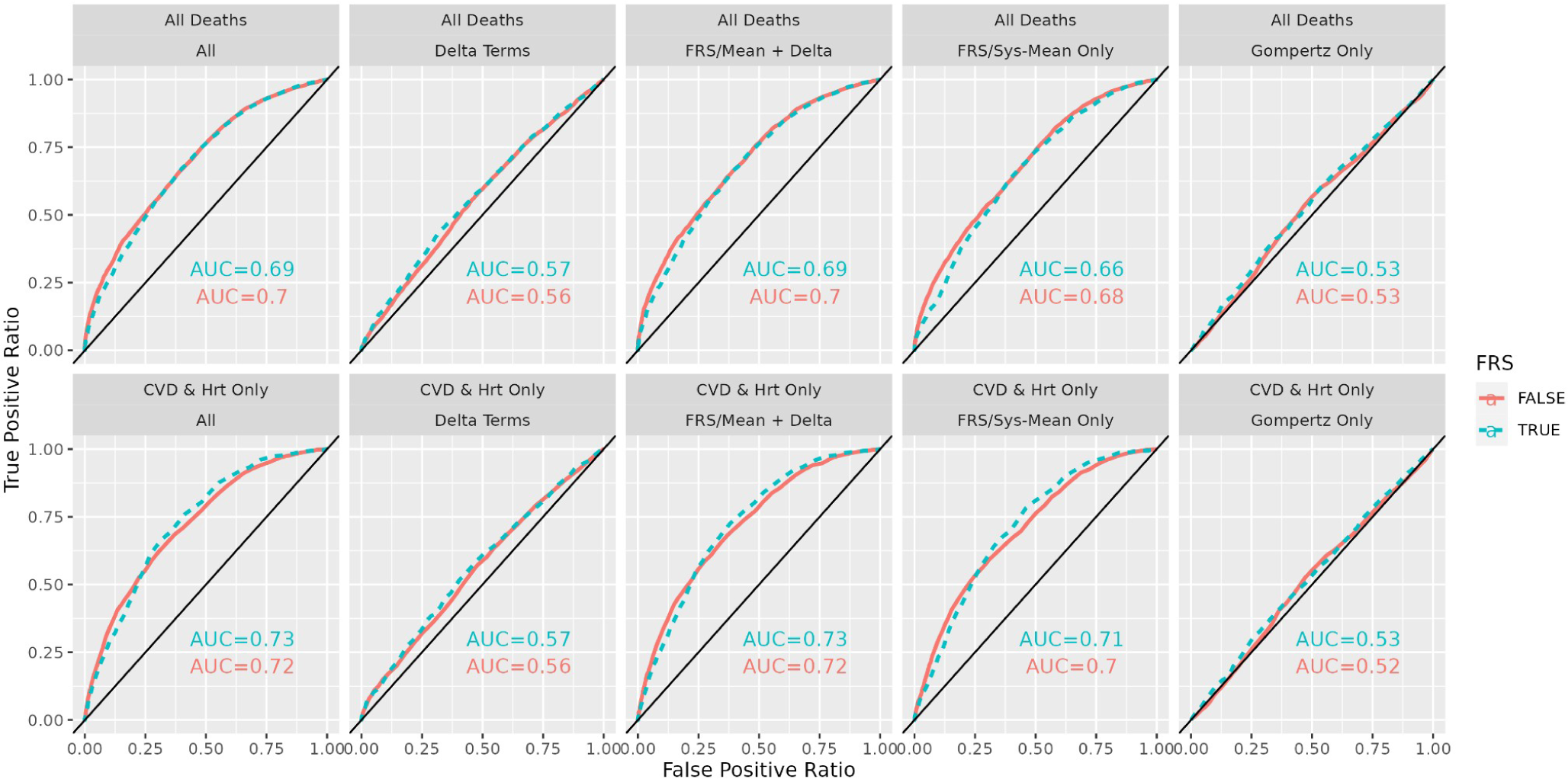
Model-based ROC curves for population with a Framingham risk score. The models shown in the first row was trained on all-death outcomes and the second row was trained specifically on CVD outcomes only. The type (solid/dashed) of the lines represents whether the model used the FRS or mean blood pressure (systolic and diastolic) in the linear predictor term. Finally, the columns differ in the choice of terms in the linear predictor, which is controlled by setting specific β-terms to be equal to zero. For example, the right-most column has all β=0. Note in particular that the predictor that including long-term variation in BP (Delta) as well as mean systolic BP (or FRS) – column 3 – increases AUC by 0.02 relative to the predictor that includes only the mean systolic BP (or FRS) (in column 4).

## Discussion

First, our findings are consistent with expectations based on known links between mean systolic BP and CV mortality(31). Our primary findings were that discrepancies between home and clinic measurements – our proxy here for medium-term BP variability – were associated with a meaningful amount of increased long-term risk of mortality, and particularly of mortality due to cardiovascular and cerebrovascular causes (CVD), and that this increased risk remained after controlling for mean BP or for Framingham risk score. There was suggestive evidence that very-short-term (1 minute apart) variability in measurements taken at home were also associated with increased long-term risk. While our results are from an observational study and are associational in nature, they suggest that variability of BP readings over several months, and possibly very-short-term variability as well, should receive increased attention as a potential predictor of increased CVD mortality risk. We also note that mean diastolic BP is found to have a highly significant negative link to all-cause mortality and to CVD mortality. This effect disappears in the primary analytic sample that has Framingham risk scores, perhaps because this subpopulation excludes the oldest subjects, which would be consistent with earlier observations that higher diastolic BP is associated with increased survival in the elderly.(32,33)

When the coefficients of the fitted models, the effect sizes, are scaled by the covariate standard deviation, we see the effect of systolic mean is about three times as large as that of the intervisit difference – about 0.33 vs 0.12 for CVD mortality, and 0.20 vs 0.07 for all-cause mortality. The association between mortality and intervisit variability is of similar strength between systolic and diastolic BP. This association is similar to that found in recent work using data from the English Clinical Practice Data Link (CPRD) between variability estimated across six visits, with 10 years of follow-up for CVD mortality; however, that study found no improvement to overall predictive value when adding variability to a risk score optimized for this data set (34).

Some of our findings appear to disagree with prior work that also examined BP variability using NHANES data(11). This prior study focused on different parts of the data, and thus interpreting these findings in light of the current analysis can improve our understanding of the relationships between different types of BP variability and cardiovascular mortality. The disagreements are only indirectly due to the differences in statistical methodology. One difference in our findings is that we find a highly significant positive effect of inter-visit variability, both systolic and diastolic, on both CVD and all-cause mortality. They found no significant effect of diastolic inter-visit variability. The most likely explanation for this is that this prior work used a different measure of inter-visit variability, based on multiple clinic measurements. This creates a less noisy proxy for LT variability, but drastically reduces the sample size. Instead of the 14,654 subjects included in our analysis (and included in subsequent analysis of within-visit variability(19)), they could only consider fewer than 1000 subjects, those who had three sets of clinic measurements.

NHANES has a complex sampling design, including oversampling of various population groups. Thus, the effect sizes estimated here cannot be expected to reflect exactly the effect sizes that prevail in the whole population. Correction of Bayesian hierarchical models for study design is a still underexplored problem in statistics, and we hope to improve our estimates in this direction in a future work.

We have not interrogated the causal implications of our results. It is possible that what we label “medium-term variation” could be a proxy for an unmeasured factor or one obscured by overlooked covariates. Our statistical analysis stratifies baseline mortality by sex, ethnicity, and age, suggesting such a hidden factor must be something less obvious. In any case, there does seem to be a factor, either inter-visit BP variability or something associated with it, that shows a notable correlation with long-term mortality.

## Data Availability

All data produced are available online at https://www.cdc.gov/nchs/nhanes/index.htm

