## Appendix A for "Short-term and Mid-term Blood Pressure Variability and long-term Mortality: evidence from the Third National Health and Nutrition Examination Study"

### NHANES Blood Pressure-Based Mortality Risk - Appendix

27/11/2023

**References** **47**

### 1 Appendix A – The data

#### 1.1 Exclusions

There were 19592 subjects in the initial data set. Of these 4573 were excluded because they had missing data or were not followed up, or belonged to the “Other” ethnic group. This left 15019 subjects for further consideration. A small number of subjects were excluded because their blood pressure measurements were outside the normal range, as described below in section 1.3.3. As our method depends on estimating the mortality rates for each demographic group (ethnicity and sex), we removed the small number of subjects whose ethnic group was given as “Other” (n=751). (The three included ethnic groups were Mexican American

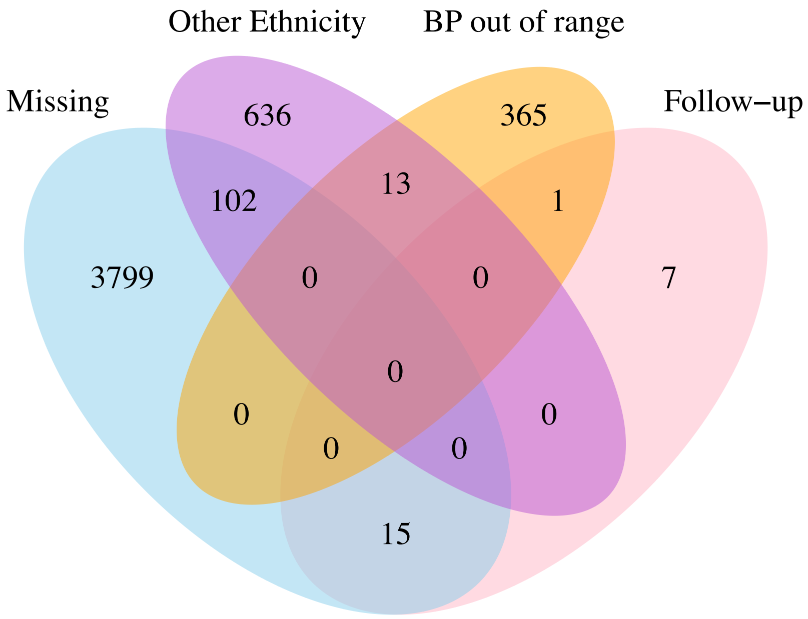

Figure 1: Venn diagram of subjects excluded from the analysis.

(n=5150), Black (n=5336), and White (n=8355). In the end there were 14654 subjects in the analysis data set. A Venn diagram of the different causes of exclusion is given in figure 1. We will refer to this as the "full population". Of these, 9008 had a computable FRS score. We call this the "FRS population' '.

#### 1.2 Exploratory data analysis

The empirical means of the home and clinic measures in population B are tabulated in Table 1. We note that the home measures are systematically higher than the clinic measures, within every demographic group, with greater differences for subjects who are white or Mexican, and female. The average difference is about 2.2 for diastolic and 2.7 for systolic, which is small compared with the general range of the differences, which have SD of 10.5 (diastolic) and 14.8 (systolic).

##### 1.2.1 Correlations between measurements

In Figure 2 see that there is relatively little correlation between empirical SD and empirical mean SD for the different BP types and places. This is reassuring, as it avoids the possibility of a collinearity effect confounding the sampling of mean and SD, which are being treated as independent covariates in the model.

In Table 2 we show the correlations between overall mean and absolute difference ($|\Delta|$) between clinic and home measurements. The results are given as a $2 \times 2$ table, showing correlations within systolic and diastolic BP, and between the two. The only moderately high correlation is between Systolic mean and Diastolic absolute Delta, which would correspond to a Variance Inflation Factor of 1.14. While this is not directly relevant to the present Bayesian methodology, it suggests that this correlation should not substantially affect the estimation of the model coefficients.

In Table 3 we show the correlations between mean and standard deviation for the three BP measures, considering all pairs of (Clinic,Home) and (Systolic,Diastolic). Finally, Table 4 shows the correlations between systolic and diastolic, ranging over (Clinic,Home) and (Mean,SD). (Some of the numbers here of course duplicate those in Table 3.) Again, the correlations are too low to require any special treatment.

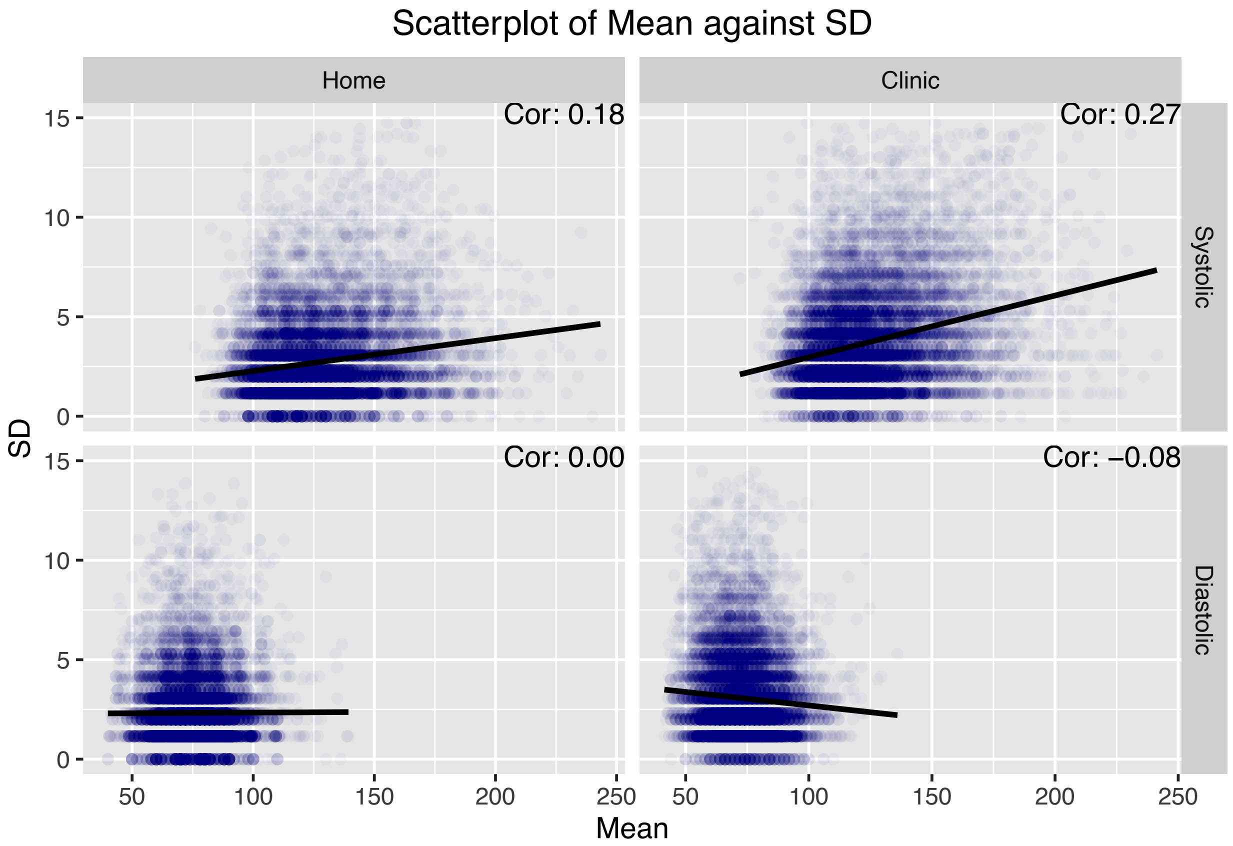

Figure 2: Scatterplot of individual mean BP against individual SD of BP

Table 1: Summary data for blood pressure

| Place | Sys/Dias | Sex | Ethnicity | Mean | SD |
|---|---|---|---|---|---|
| Home | Systolic | Male | Black | 128.8 | 2.7 |
| Home | Systolic | Male | White | 131.2 | 2.8 |
| Home | Systolic | Male | Mexican | 127.1 | 2.5 |
| Home | Systolic | Female | Black | 123.2 | 2.7 |
| Home | Systolic | Female | White | 127.2 | 2.9 |
| Home | Systolic | Female | Mexican | 121.2 | 2.7 |
| Home | Diastolic | Male | Black | 78.7 | 2.4 |
| Home | Diastolic | Male | White | 77.1 | 2.3 |
| Home | Diastolic | Male | Mexican | 76.8 | 2.4 |
| Home | Diastolic | Female | Black | 75.0 | 2.4 |
| Home | Diastolic | Female | White | 73.7 | 2.3 |
| Home | Diastolic | Female | Mexican | 72.0 | 2.4 |
| Clinic | Systolic | Male | Black | 127.7 | 3.5 |
| Clinic | Systolic | Male | White | 128.0 | 4.2 |
| Clinic | Systolic | Male | Mexican | 123.5 | 3.5 |
| Clinic | Systolic | Female | Black | 122.4 | 3.6 |
| Clinic | Systolic | Female | White | 123.5 | 4.1 |
| Clinic | Systolic | Female | Mexican | 117.8 | 3.4 |
| Clinic | Diastolic | Male | Black | 77.8 | 3.1 |
| Clinic | Diastolic | Male | White | 75.4 | 3.1 |
| Clinic | Diastolic | Male | Mexican | 74.9 | 3.2 |
| Clinic | Diastolic | Female | Black | 72.4 | 3.0 |
| Clinic | Diastolic | Female | White | 70.5 | 3.0 |
| Clinic | Diastolic | Female | Mexican | 69.2 | 3.0 |

Table 2: Correlation between mean and Delta. Rows correspond to type of Delta, columns to type of mean.

| | SysMean | DiasMean |
|---|---|---|
| SysDelta | 0.137 | 0.014 |
| DiasDelta | 0.347 | 0.128 |

Table 3: Correlation between mean and SD. Rows correspond to type and location of SD, columns to type and location of mean.

| | Clinic Sys Mean | Home Sys Mean | Clinic Dias Mean | Home Dias Mean |
|---|---|---|---|---|
| Clinic Sys SD | 0.269 | 0.254 | 0.095 | 0.106 |
| Home Sys SD | 0.157 | 0.177 | 0.047 | 0.073 |
| Clinic Dias SD | 0.041 | 0.030 | -0.076 | -0.037 |
| Home Dias SD | 0.045 | 0.060 | 0.010 | 0.003 |

#### 1.3 Errors in blood pressure measurement or recording

The blood pressure measurement or recording errors were found particularly in the home measurements. While these did not destroy the usefulness of the home measurements, they did require some attention and decisions for how to work with these defects. We also consider them inherently interesting, and worth

Table 4: Correlation between diastolic and systolic summary statistics. Rows correspond to variables and locations for diastolic, columns to variables and locations for systolic.

| | Clinic Sys SD | Clinic Sys Mean | Home Sys SD | Home Sys Mean |
|---|---|---|---|---|
| Clinic Dias SD | 0.150 | 0.041 | 0.007 | 0.030 |
| Clinic Dias Mean | 0.095 | 0.497 | 0.047 | 0.378 |
| Home Dias SD | 0.022 | 0.045 | 0.224 | 0.060 |
| Home Dias Mean | 0.106 | 0.405 | 0.073 | 0.547 |

registering for future researchers working on these or similar data. In particular, the problem we have called "dependent replication" was entirely unexpected, although not unprecedented, and is of particular concern to researchers trying to estimate individual variation in clinically relevant measures.

###### 1.3.1 Last-digit preference

Mild tendency for observers to prefer certain last digits in reporting BP measurements has been reported in other studies, though an analysis of the 1999 wave of NHANES reported no last-digit preference (Ostchega et al. 2003).

The last-digit preference in NHANES III, on the other hand, is substantial, with about 26.7% of all the clinic-measured systolic BP measurements ending in 0, but only about 31.9% ending in 4 or 6. Because the shifts due to last-digit preference are presumably small, we expect them to have little effect on the main effects that we are examining in this paper, but they do increase the probability of two measurements being rounded to the same value, something that needs to be taken into account in examining the problem of dependent replication.

Table 5: Summary data for BP end digits

| Place | Sys/Dias | 0 | 2 | 4 | 6 | 8 |
|---|---|---|---|---|---|---|
| Home | Systolic | 0.240 | 0.199 | 0.159 | 0.169 | 0.233 |
| Home | Diastolic | 0.186 | 0.179 | 0.198 | 0.217 | 0.219 |
| Clinic | Systolic | 0.267 | 0.188 | 0.160 | 0.159 | 0.226 |
| Clinic | Diastolic | 0.192 | 0.189 | 0.209 | 0.212 | 0.198 |

###### 1.3.2 Dependent replicates

While the protocol calls for each subject to have three independent BP measures taken, it is not impossible that the observers may have been influenced by one measure in recording the next. This could happen in either direction: later measurements could be pulled closer to the first, or there could be an inclination to avoid repeated measures. This is relevant, because erroneously repeated measures would artificially decrease the variance of the three measurements, and avoiding repeated measures would have the opposite effect.

The end-digit bias may be expected to have an effect here, since it influences the probability of two measurements being rounded to the same value. We begin by noting the standard deviations for measurements of individual subjects as given in the column 'Mean of SD' in Table 6. The column 'Prob all rep' gives the theoretical probability that two of the three measurements for a subject would have the same value, if the measurements were independent and normally distributed with the given standard deviation (adjusted for the rounding), and assuming that rounding to particular digits is done in proportion to the fractions listed in Table 5. The column 'Prob 2 rep' gives the probability that two of the three measurements would have the same value, under the same conditions. The column 'Frac all rep' gives the observed fraction of subjects for whom all three measurements were equal, and 'Frac 2 rep' gives the fraction for whom two of the three measurements were equal. The observed fractions for three equal measurements are all very close to the theoretical probabilities, but the observed fractions for two equal measurements are substantially lower than the theoretical probabilities. (For comparison, a 95% probability range for the fraction of subjects with two equal measurements is about $\pm 0.008$.)

In Figure 3, we show the fraction of subjects with two equal measurements, by examiner, blocked by place and type. We see that the fraction of subjects with two equal measurements varies substantially by examiner, and that the variation is greater for the systolic than for the diastolic measurements.

Table 6: Summary data for repeated measures

| Place | Sys/Dias | Mean of SD | Frac all rep | Prob all rep | Frac 2 rep | Prob 2 rep |
|---|---|---|---|---|---|---|
| Home | Systolic | 2.739 | 0.050 | 0.048 | 0.432 | 0.510 |
| Home | Diastolic | 2.343 | 0.063 | 0.061 | 0.488 | 0.564 |
| Clinic | Systolic | 3.775 | 0.024 | 0.028 | 0.355 | 0.400 |
| Clinic | Diastolic | 3.082 | 0.028 | 0.036 | 0.414 | 0.459 |

We show the fraction of subjects with two equal measurements in Figure {fig:examinerPlot}, split by examiner, blocked by place and type. We see that the fraction of subjects with two equal measurements varies substantially by examiner, and that the variation is greater for the systolic than for the diastolic measurements.

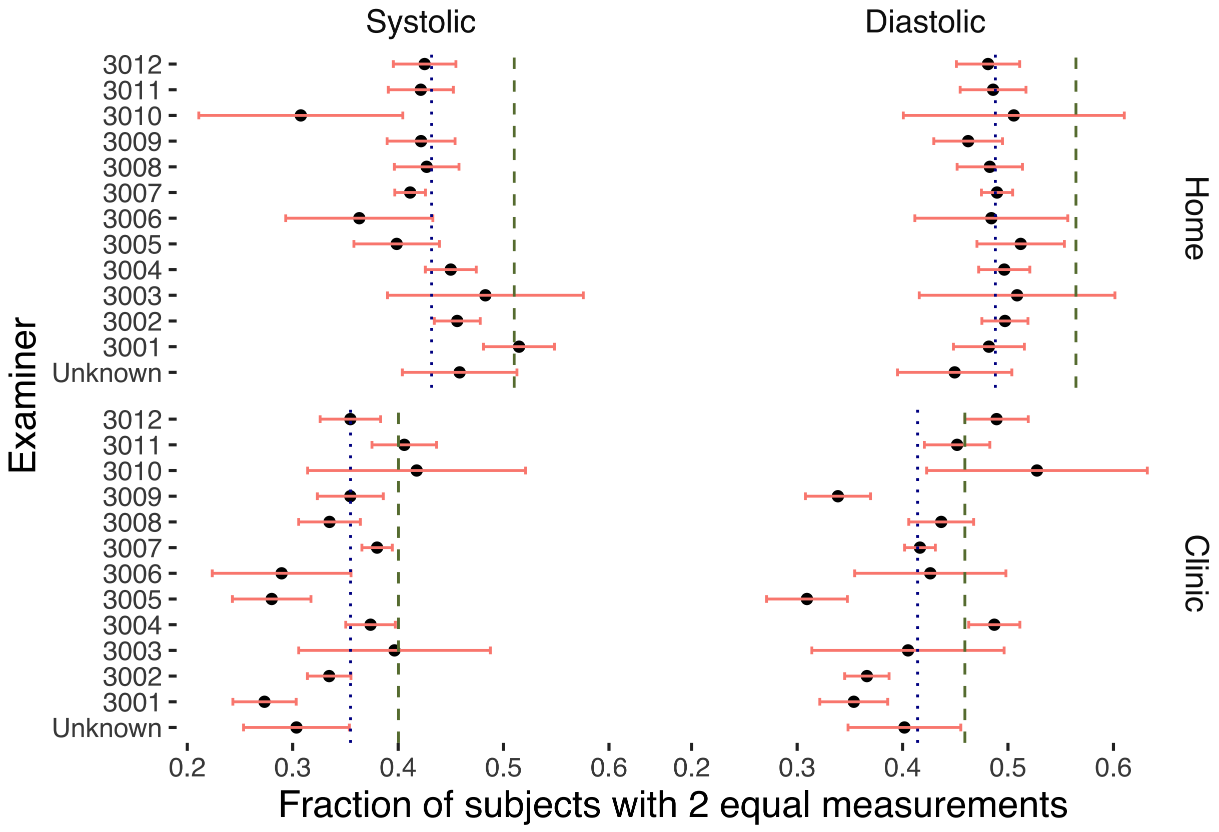

Figure 3: Number of subjects with 2 equal measurements by examiner, blocked by place and type. Red band shows 95% probability range. Vertical green dashed line shows expected fraction; blue dotted line shows observed fraction over all examiners.

In Figure 4, we show the fraction of subjects with three equal measurements, by examiner, blocked by place and type. Relative to the expected random fluctuations, we see that there is even more variation among

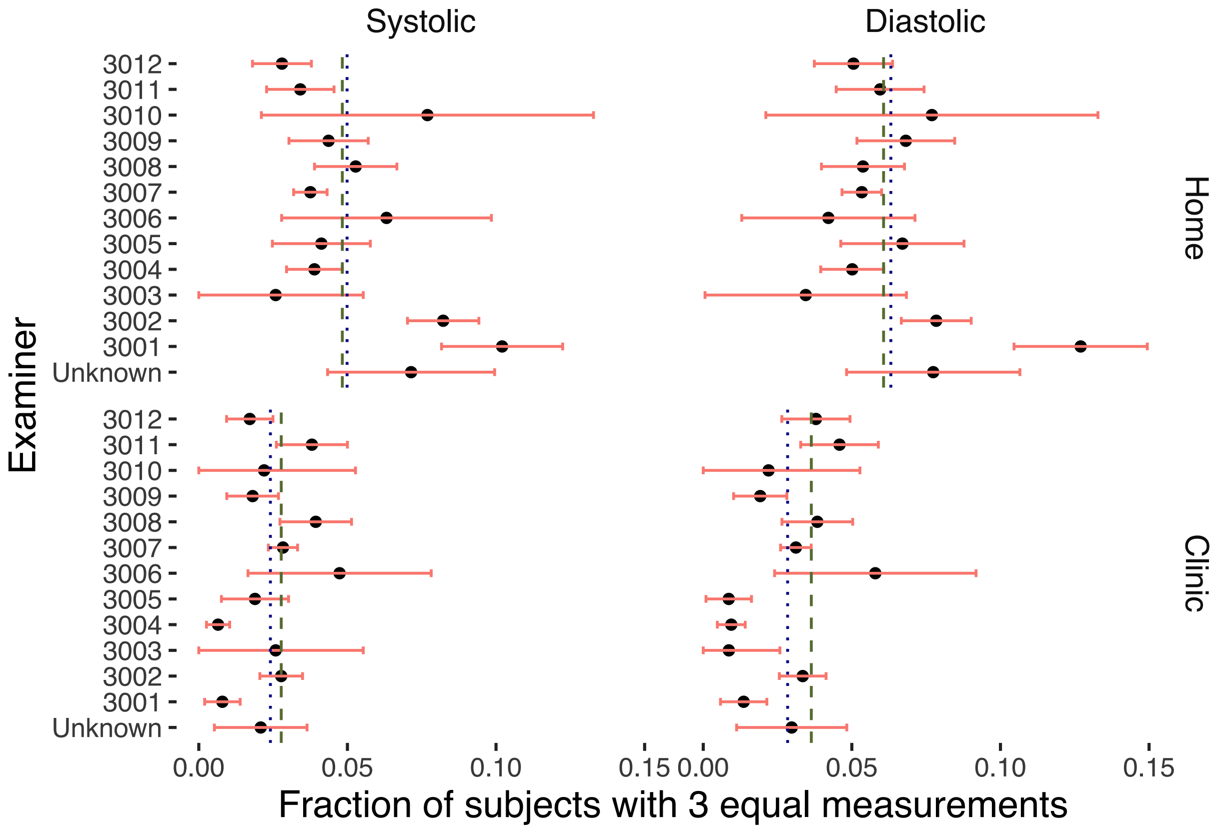

Figure 4: Number of subjects with 3 equal measurements by examiner, blocked by place and type. Red band shows 95% probability range. Vertical green dashed line shows expected fraction; blue dotted line shows observed fraction over all examiners.

the examiners. One examiner (3001) produced consistently excessive numbers of triple repeats in Home measurements, and a deficit of triple repeats in Clinic measurements.

One further point to explore is the position of the two equal measures in a group of three. If there are three independent measures, with two equal, each of the three has equal probability of being the odd one out. On the other hand, if there is a trend in the measurements, then the second is least likely to be the odd one out.

In fact, what we observe is that it is the third measurement that is least likely to differ from the other two, while the first is most likely. This is what we would expect if examiners sometimes either intentionally copied the second measurement into the space for the third, or unintentionally allowed themselves to be influenced into observing the same number. The proportions are listed in Table 7, together with chi-squared tests for difference from the expected equal proportions for each site and type. On the other hand, if there is a trend in the measurements, then the second is least likely to be the odd one out, which is also not what we see.

We see that there is a huge deviation from the expected proportions in the Home measurements, but less in the Home measurements, and more deviation in Systolic than in Diastolic measurements.

Table 7: Chi-square test for difference between observed proportions (all examiners), stratified by place and type

| Place | Sys/Dias | Freq1 | Freq2 | Freq3 | ChiSq | p-value |
|---|---|---|---|---|---|---|
| Home | Systolic | 2657 | 2149 | 1522 | 306.0 | 3.57e-67 |
| Home | Diastolic | 2864 | 2541 | 1746 | 278.0 | 4.3e-61 |
| Clinic | Systolic | 1905 | 1702 | 1594 | 28.8 | 5.57e-07 |
| Clinic | Diastolic | 2172 | 1992 | 1906 | 18.2 | 1.12e-04 |

To explore this further, we can look at the proportions of first, second and third measurements from each examiner that are different from the other two. The results of a chi-squared test for each examiner (stratified by site and type of BP) for difference from the expected equal proportions are shown in Figure 5. The dashed line represents a p-value of 0.001. Here we see that the Home measurements are extremely variable, while the Clinic measurements are quite consistent with the expected proportions, with the single exception of examiner 3004, who is far from the expected equal proportions in all categories of measurement.

Given that the position of the differing measure clearly differs from the expected equal proportions, we might ask whether the examiners agree on a common proportion, suggesting that there might be some underlying systematic (observer-independent) reason for the differing measurements. In Table 8 we show the results of a chi-squared test for equality of observed proportions among the examiners, stratified by place and type. Interestingly, we see here that the examiners are fairly consistent in their proportions for the Home measures, but not for the Clinic measures.

Table 8: Chi-square test for difference between observed proportions among the examiners, stratified by place and type

| Place | Sys/Dias | ChiSq | p-value |
|---|---|---|---|
| Home | Systolic | 32.8 | 1.69e-01 |
| Home | Diastolic | 38.9 | 5.01e-02 |
| Clinic | Systolic | 60.0 | 1.67e-04 |
| Clinic | Diastolic | 110.0 | 3.05e-12 |

Looking at a ternary plot Figure 6 for the proportions from the 13 different examiners, we see very clearly the bias toward having the last two measures agree, for almost all examiners, and examiner 3004 (marked larger) standing out as a clear outlier.

Overall, we can only conclude that there are clearly some irregularities in the BP measurement process, but

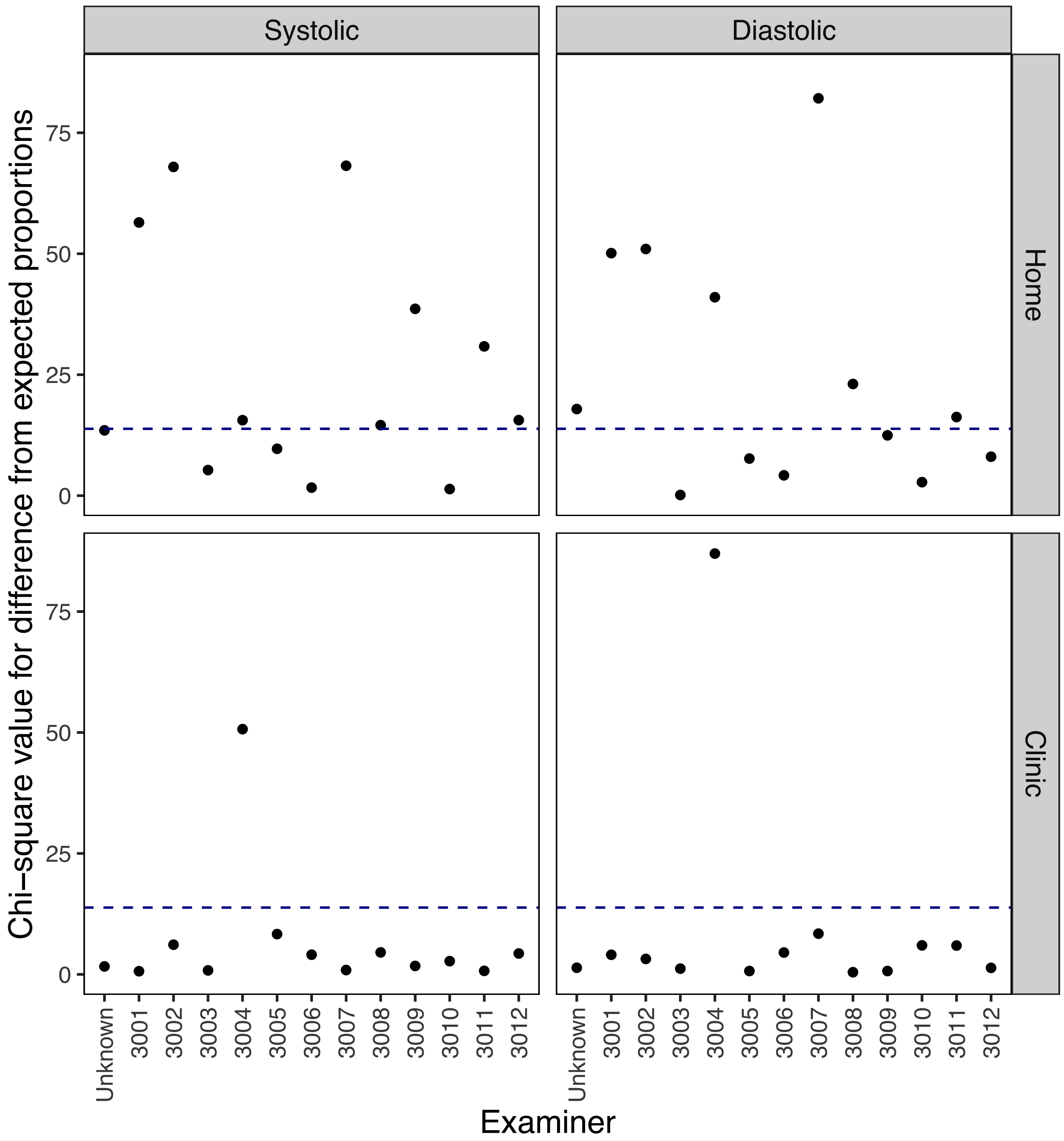

Figure 5: Proportions of first, second and third measurements from each examiner that are different from the other two, by place and type. Chi-squared value for difference from expected proportions. Dashed line represents p-value 0.001.

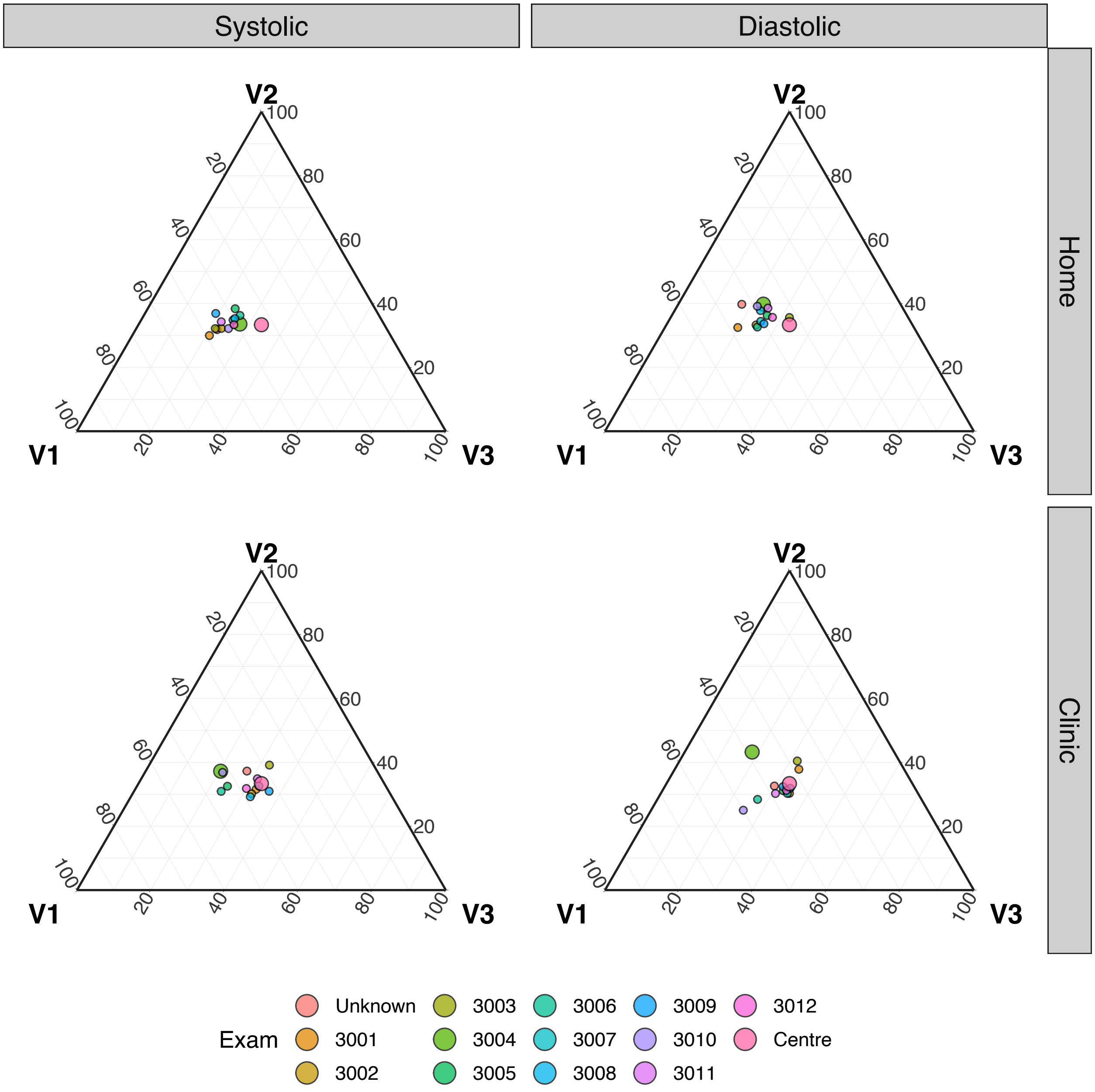

Figure 6: Ternary plot of the position of the measurement that is unique, among subjects with 2 equal measurements. V1 is the fraction with the first distinct, V2 is the fraction with the second distinct, V3 is the fraction with the third distinct.

we cannot identify a specific structure to them, or propose a remedy. As the irregularities are not very large, we will proceed with the analysis without attempting to correct for them.

##### 1.3.3 Missing or implausible measurements

Some of the reported measures were extremely implausible, particularly for diastolic BP. NAsubjects had at least one diastolic BP measure recorded as 0, in addition to the 3916 subjects who were missing at least one measurement. We excluded all of these subjects, and indeed any subject who had at least one measurement recorded outside the ranges (40,140) for diastolic and (60,250) for systolic BP, as recommended by the CDC (Littman et al. 2012). There was just one subject with systolic BP measures that were too low, but NA subjects with low diastolic BP (in addition to those with measures recorded as 0). One subject was excluded for diastolic BP 156, and three were excluded for systolic BP that was too high, with the maximum being 264.
