## Appendix B for "Short-term and Mid-term Blood Pressure Variability and long-term Mortality: evidence from the Third National Health and Nutrition Examination Study"

### 2 Appendix B – Model details

This appendix aims to add more detail about the numerical modelling than was provided in the article. This is to ensure that the research methods are transparent and entirely reproducible. The numerical modelling presented in this paper was performed using R combined with Rstan. More detail will be provided here about the model, about the specific methodology used to parameterize the model, and more results are provided that were not included in the main text.

#### 2.1 The Statistical Model

The model used in this research is built from the theory of joint modelling of longitudinal and time-to-event data. This will be described in detail later on in this section, however, in brief, this allows the simultaneous modelling of both longitudinal observation data (in this article, this is blood pressure measurements) and also the time-to-event outcome. In this research the event of interest is either death from any cause, or death from specifically cardiovascular or cerebrovascular causes. We henceforth will refer to this latter mortality as CVD. In the latter case, death from a different cause is treated as a noninformative censoring event.

##### 2.1.1 Survival Analysis (Time-to-Event)

The basic survival model is a Gompertz hazard rate with proportional hazards influences of the blood pressure covariates. The Gompertz equation

$$h_0(t) = B \exp\left(\theta(x + T)\right), \tag{1}$$

describes the baseline hazard of the population to a particular risk, which, for this article, investigates CVD mortality specifically, as well as studying mortality risk in general. $x \in \mathbb{N}^{\mathbb{N}}$ is the age of the individual at the initial interview time, for $N$ the number of individuals, and $T \in \mathbb{R}^{+,N}$ the time since the individual entered the survey. Note that both $B$ and $\theta$ have 6 different values, depending on the sex reported at the initial interview – female or male — or the race — black, white or 'other'. Note that 'other' in the race category is a combination of all non-black or non-white racial identities, such as Hispanic populations. The log-linear proportional hazards model links the covariates of the model (mean systolic blood pressure, variance in the diastolic blood pressure, etc) to the survival outcome of the individual via the equation

$$h(t) = h_0(t) \exp\left(\beta \cdot (X - \hat{X})\right), \tag{2}$$

where $X \in \mathbb{R}^{+,N \times d}$ is a vector of summary statistics of the blood pressure measurements of individual covariates in our model, $\hat{X} \in \mathbb{R}^{+,d}$ is the centering of the covariates such that the equation $\sum_i^N \exp\left(\beta \cdot (X - \hat{X})\right) = 0$ is approximately satisfied (more on this later), and $\beta \in \mathbb{R}^d$ implies the strength of the influence of the covariate on the mortality risk. The majority of mortality events are censored — not yet known at the time of data collection — the censoring indicator being notated as $\delta \in \{0, 1\}$. When CVD mortality is the event being analysed, deaths due to other causes are treated as noninformative censoring events. In this study, we explored the following covariates:

| Variable Name | Support | Description |
|---|---|---|
| $FRS-1998$ | $R^N$ | 1998 version of the FRS score |
| $FRS-ATP$ | $R^N$ | ATP version of the FRS score |
| $M_S$ | $R^{+,N}$ | Mean systolic blood pressure |
| $M_D$ | $R^{+,N}$ | Mean diastolic blood pressure |
| $\Delta_S$ | $R^{+,N}$ | Semi-difference between Home and Clinic mean systolic blood pressure |
| $\Delta_D$ | $R^{+,N}$ | Semi-difference between Home and Clinic mean diastolic blood pressure |
| $\sigma_{\{S,H\}}$ | $R^{+,N}$ | Standard deviation of the systolic blood pressure taken at home |
| $\sigma_{\{D,H\}}$ | $R^{+,N}$ | Standard deviation of the diastolic blood pressure taken at home |
| $\sigma_{\{S,C\}}$ | $R^{+,N}$ | Standard deviation of the systolic blood pressure taken at the clinic |
| $\sigma_{\{D,C\}}$ | $R^{+,N}$ | Standard deviation of the diastolic blood pressure taken at the clinic |
| $\tau_{\{S,H\}}$ | $R^{+,N}$ | Precision of the systolic blood pressure taken at home |
| $\tau_{\{D,H\}}$ | $R^{+,N}$ | Precision of the diastolic blood pressure taken at home |
| $\tau_{\{S,C\}}$ | $R^{+,N}$ | Precision of the systolic blood pressure taken at the clinic |
| $\tau_{\{D,C\}}$ | $R^{+,N}$ | Precision of the diastolic blood pressure taken at the clinic |

Please note that the last four elements of this list, the precision values, were only carried out to ensure model consistency with the use of standard deviation instead. Note as well that the $\Delta$ covariates, representing the medium-term variability, enter into the log relative risk sum as an **absolute value**.

For the parametrization of this model, we assume that the Gompertz parameters and the parameters in the linear predictor term have prior distributions as follows:

$$\begin{aligned} B &\sim \mathcal{N}(0, 2), \\ \theta &\sim \mathcal{N}(0, 2), \\ \beta &\sim \text{Cauchy}(0, 100). \end{aligned} \tag{3}$$

The likelihood for this Gompertz proportional hazards model, over all individuals in the census, is as follows:

$$L_S(v,\delta) = \prod_i^N f(v_i, \delta_i | B_i, \theta_i, \beta_i, X, \hat{X}) = \prod_i^N h(v_i | B_i, \theta_i, \beta_i, X, \hat{X})^{\delta_i} \exp\left(-\sum_i^N H(v_i | B_i, \theta_i, \beta_i, X, \hat{X})\right), \tag{4}$$

with $H(v) = \int_0^v h(w)\mathrm{d}w$ the cumulative hazard.

##### 2.1.2 Longitudinal Modelling

The mortality hazard rates are assumed to be influenced by individual-level blood pressure means and variability characteristics. These characteristics are not directly observed, but are inferred from their influence on the individual blood pressure measurements, which have been observed. Let $Y_i(t_j)$ be the observed blood pressure for patient $i$ at time $t_j$, for the individual $i \in 1, 2, ..., N$ and the number of blood pressure measurements per individual $j \in 1, 2, ..., k$. Due to the fact that the blood pressure measurement data was taken at both the home and clinic (written using subscripts H and C, respectively), with approximately 6 months between these two measurements, we model the blood pressure using the following model, assuming the diastolic $Y_i^D$ and systolic $Y_i^S$ blood pressure to be Gaussian-distributed:

$$\begin{aligned} (Y_i^D)_H &\sim \mathcal{N}(M_i^D + \Delta_i^D, (\sigma_i^D)_H), \\ (Y_i^D)_C &\sim \mathcal{N}(M_i^D - \Delta_i^D, (\sigma_i^D)_C), \\ (Y_i^S)_H &\sim \mathcal{N}(M_i^S + \Delta_i^S, (\sigma_i^S)_H), \\ (Y_i^S)_C &\sim \mathcal{N}(M_i^S - \Delta_i^S, (\sigma_i^S)_C), \end{aligned} \tag{5}$$

where superscripts $D$ and $S$ refer to diastolic and systolic blood pressure, respectively.

The blood pressure characteristics — the individual-level parameters — are themselves distributed according to a hierarchical model, determined by population-level parameters (also called "hyperparameters' '):

$$\begin{aligned} M_i^{\{D,S\}} &\sim \mathcal{N}(\mu_M^{\{D,S\}}, \sigma_M^{\{D,S\}}), \\ \Delta_i^{\{D,S\}} &\sim \mathcal{N}(\mu_D^{\{D,S\}}, \sigma_D^{\{D,S\}}), \\ \sigma_{i,C}^{\{D,S\}} &\sim \Gamma(r_C^{\{D,S\}}, \lambda_C^{\{D,S\}}), \\ \sigma_{i,H}^{\{D,S\}} &\sim \Gamma(r_H^{\{D,S\}}, \lambda_H^{\{D,S\}}). \end{aligned} \tag{6}$$

The longitudinal outcome modelling therefore aims to infer these hyperparameters

$$\Theta = \left\{\mu_M^{\{D,S\}}, \mu_D^{\{D,S\}}, \sigma_M^{\{D,S\}}, \sigma_D^{\{D,S\}}, r_C^{\{D,S\}}, \lambda_C^{\{D,S\}}, r_H^{\{D,S\}}, \lambda_H^{\{D,S\}}\right\}, \tag{7}$$

and to use the implied uncertainty about the individual-level parameters to inform the inference about the survival parameters. The likelihood for the longitudinal measurements is therefore (combining the systolic and diastolic into a single parameter for simplicity):

$$L_L(\Theta|Y) = \prod_{i=1}^{N} \left( \prod_{j=1}^{k} f(y_{ij}|M_i, \Delta_i, \sigma_i) \right) f(M_i|\mu_M, \sigma_M) f(\Delta_i|\mu_D, \sigma_D) f(\tau_{i,C}|r_C, \lambda_C) f(\tau_{i,H}|r_H, \lambda_H) \tag{8}$$

##### 2.1.3 Combined Hierarchical Model

Combining the longitudinal outcome and time-to-event partial likelihoods, and for a given parameter space value of $\Omega = \{\beta, B, \theta\} \cup \Theta$, the joint likelihood is

$$\begin{aligned} L(\Omega|Y) = \prod_{i=1}^{N} \left( \prod_{j=1}^{k} f(y_{ij}|M_i, \Delta_i, \sigma_i) \right) & f(v_i, \delta_i|B_i, \theta_i, \beta_i, X, \hat{X}) f(M_i|\mu_M, \sigma_M) \\ & f(\Delta_i|\mu_D, \sigma_D) f(\tau_{i,C}|r_C, \lambda_C) f(\tau_{i,H}|r_H, \lambda_H). \end{aligned} \tag{9}$$

One approach to estimating the complete set of hyperparameters

$$\Omega_H = \{\mu_B, \sigma_B, \mu_\theta, \sigma_\theta, \mu_\beta, \sigma_\beta, \mu_M^{\{D,S\}}, \sigma_M^{\{D,S\}}, \mu_D^{\{D,S\}}, \sigma_D^{\{D,S\}}, r_C^{\{D,S\}}, \lambda_C^{\{D,S\}}, r_H^{\{D,S\}}, \lambda_H^{\{D,S\}}\} \tag{10}$$

is to impose a higher-level prior distribution, and use the machinery of Bayesian inference to produce posteriors for everything. This approach runs into computational difficulties, which have led us to a two-stage 'empirical Bayes' approach, where the hyperparameters for the longitudinal model are first fixed by a maximum-likelihood calculation, after which the remaining hyperparameters and individual-level parameters can be estimated with Bayesian machinery. For the time-to-event parameters we choose flat hyperpriors, selecting the hyperparameters $\mu_B = \mu_\theta = \mu_\beta = 0$, $\sigma_B = \sigma_\theta = 2$, and $\sigma_\beta = 100$.

##### 2.1.4 The modelling variants

In this article, we researched into 16 variants of the model-fitting problem, but focussed mainly on 8 of them. The 8 main models use the standard deviation, $\sigma$, as the measure of the influence of blood-pressure variability on mortality. We also produced the same 8 models but using precision, $\tau = 1/\sigma^2$, as the measure of the influence of blood-pressure variability on mortality. However, this was only to ensure that there were no differences between the use of one over the other. Throughout the remainder of this appendix, we refer to the 8 main models using the following run numbers:

1. All participants (14,654), using mean systolic and diastolic blood pressure (not FRS) in the linear predictor term, with the outcome data as death specifically from CVD.

2. All participants (14,654), using mean systolic and diastolic blood pressure (not FRS) in the linear predictor term, with the outcome data as all-causes of death.

3. Only participants that had data from which FRS values could be computed (N=9,008) — the "FRS population" but using mean systolic and diastolic blood pressure (not FRS) in the linear predictor term, with the outcome data as death specifically from CVD.
4. FRS population, but using mean systolic and diastolic blood pressure (not FRS) in the linear predictor term, with the outcome data as all-causes of death.
5. FRS population, and using the FRS ATP-III value in the linear predictor term, with the outcome data as death specifically from CVD.
6. FRS population, and using the FRS ATP-III value in the linear predictor term, with the outcome data as all-causes of death.
7. FRS population, and using the FRS 1998-version value in the linear predictor term, with the outcome data as death specifically from CVD.
8. FRS population, and using the FRS 1998-version value in the linear predictor term, with the outcome data as all-causes of death.

We also include Directed Acyclical Graph (DAG) sketches to help visualize the different models, as shown in figures 7 and 8. In order to read the DAGs, note that each square background layer that appears as a stack of layers represents different measured outcomes that were made in the first wave of the survey. The outcome variables measured are represented by a square-shaped text box, and a parameter of the model is represented by a circular-shaped text box. If either a square or circular text box is placed on top of a stacked rectangular layer, it means that multiple values of that variable (as many as there are layers to the stack) are either measured (for outcome variables) or simulated (for parameters of the model). Please note that the number of layers in the stack is written in the text box that does not contain a frame which is intentionally displayed on top of the stacked layer that it represents. For example, $i = 1, ..., N$. Finally, the direction of the arrows implies causality assumed in the model.

The distribution of the blood pressure parameters in the population are derived from the model, and are summarised with other outputs of the model in Table 10 of Appendix C.

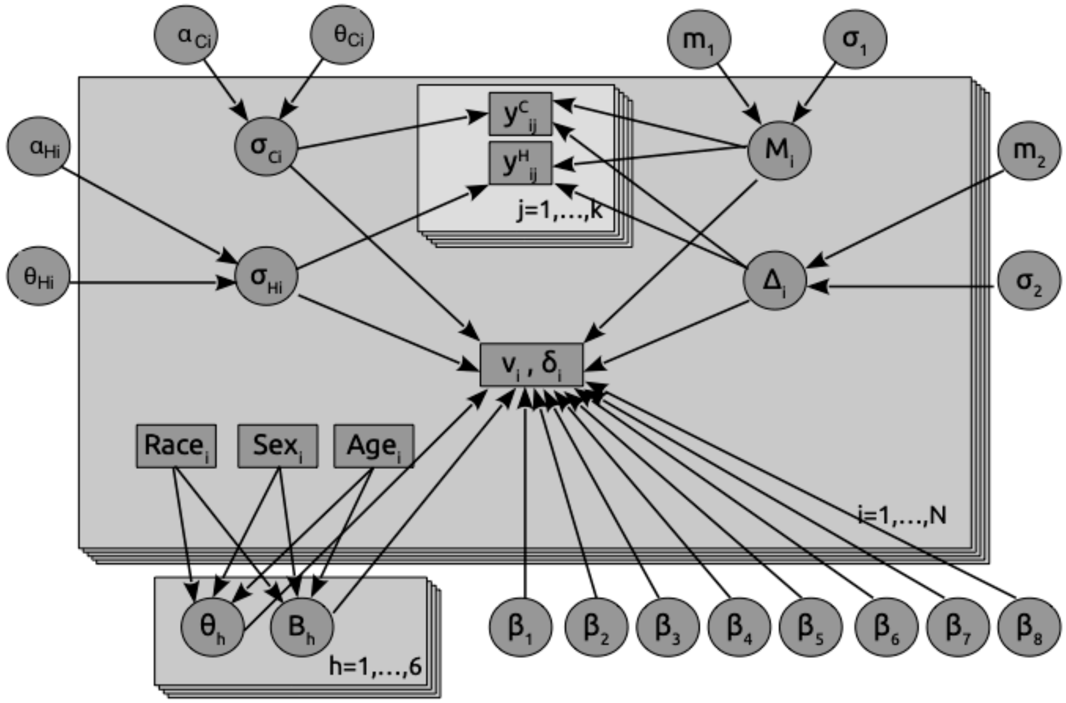


Figure 7: An illustration of the DAG of the mean blood pressure-based model presented in this article.

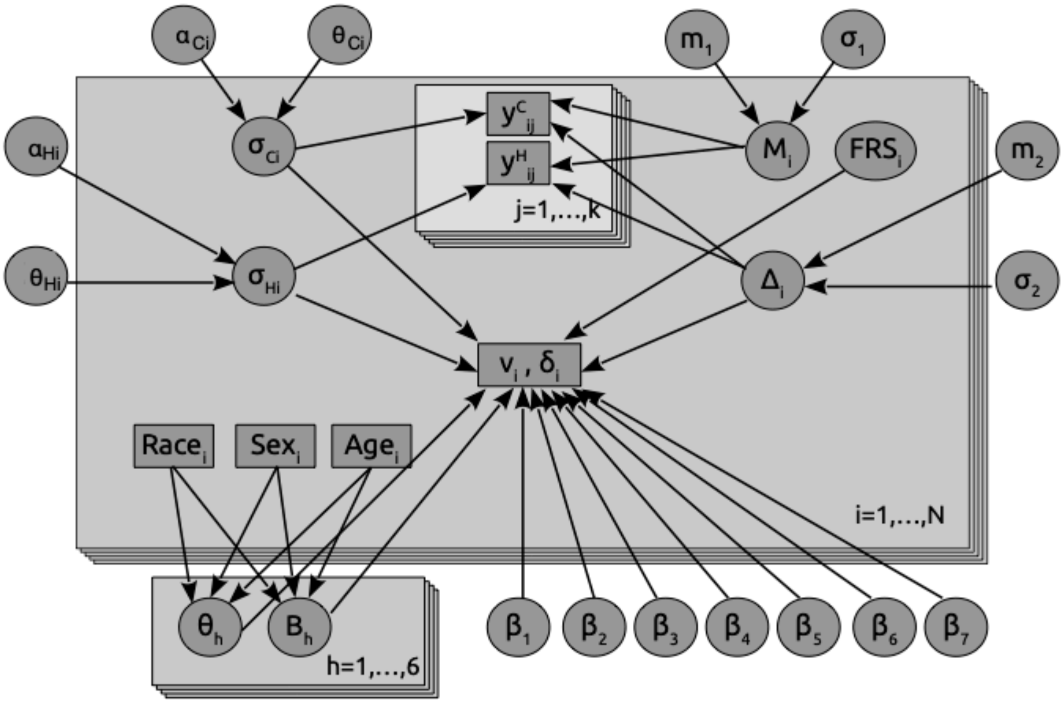


Figure 8: An illustration of the DAG of the FRS-based model presented in this article.

#### 2.2 Computational methodology

The methodology for this research can be split into three main sections: 1) calculating the empirical Bayes' parameters, 2) parameterizing the model using Hamiltonian Monte Carlo (HMC) and 3) re-centering the variables in the linear predictor equation. By applying empirical Bayes', Maximum Likelihood Estimates (MLEs) of some of the parameter distributions are provided. Note that the parameters estimated here are only the prior distribution of the global (not individual) blood pressure means and the variances, for both systolic and diastolic and home and clinic measurements. These estimates are then provided as prior distributions for the Stan MCMC simulations using HMC, where estimates can be made for all the parameter distributions of the model, given the specific centering applied. Finally, section (3) recalculates the centering values based on the previous MCMC iteration, and sets of the next iteration, while simultaneously checking for convergence in both the MCMC simulations and the centering values.

#### 2.3 Empirical Bayes Parameters

First, we extract the intervals for the digits in the blood pressure measurement recordings. Suppose the fractions of digits 0,2,4,6,8 are $b_0, b_2, b_4, b_6, b_8$. Letting $B_0 = 0$ and $B_k = 10\sum_{j=0}^{k-1} b_{2j}$ for $k = 1, \ldots, 5$, we want to choose a positive $a$ and place breaks at $-a + B_k$, so that measurements between $-a + B_k$ and $-a + B_{k+1}$ modulo 10 are assigned the final digit $2k$, for $k = 0, \ldots, 4$. We choose $a$ to minimise the total distance of the intervals from the rounded value:

$$\sum_{k=0}^{4} \int_{-a+B_k}^{-a+B_{k+1}} |x - 2k| \mathrm{d}x = \frac{1}{2} \sum_{k=0}^{4} (-a + B_k - 2k)^2 + (-a + B_{k+1} - 2k)^2,$$

as long as $2k$ is in the appropriate interval. This is minimized at

$$a = \frac{1}{5}(B_1 + B_2 + B_3 + B_4 - 15) = \sum_{j=0}^{3} (8 - 2j) b_{2j} - 3.$$

Next step, we fit the BP distribution parameters. We suppose that each individual has BP measures $\tilde{y}_{ij}^l$ for $i = 1, \ldots, n$, $j = 1, \ldots, k$ (default $k = 3$), and $l = 1, 2$, which are rounded versions of

$$y_{ij}^l \sim \mathcal{N}(\mu_i^l, (\tau_i^l)^{-1}),$$

where

$$\begin{aligned}
\mu_i^1 &= (M_i + \Delta_i)/2, \\
\mu_i^2 &= (M_i - \Delta_i)/2, \\
M_i &\sim \mathcal{N}(m_M, \sigma_M^2) \text{ and } \Delta_i \sim \mathcal{N}(m_\Delta, \sigma_\Delta^2) \text{ independent}, \\
\tau_i^l &\sim \text{Gamma}(\alpha^l, \alpha^l/\theta^l).
\end{aligned}$$

(Note that $\alpha^l$ is the usual shape parameter, while $\theta^l$ is the expectation.)

We wish to estimate the eight parameters

$$(m_M, m_\Delta, \sigma_M^2, \sigma_\Delta^2, \alpha^1, \theta^1, \alpha^2, \theta^2)$$

We begin by assuming $y_{ij}^l$ observed directly. We estimate by maximising the partial likelihood on the observations

$$\begin{aligned}
\bar{y}_{i+} &:= \frac{1}{2k} \sum_{j=1}^k (y_{ij}^1 + y_{ij}^2), \\
\bar{y}_{i-} &:= \frac{1}{2k} \sum_{j=1}^k (y_{ij}^1 - y_{ij}^2), \\
s_i^l &:= \frac{1}{k-1} \sum_{j=1}^k \Big( y_{ij}^l - \frac{1}{k} \sum_{j=1}^k y_{ij}^l \Big)^2.
\end{aligned}$$

Note that

$$(k-1) s_i^l \tau_i^l = \sum_{j=1}^k \Big( z_{ij}^l - \frac{1}{k} \sum_{j=1}^k z_{ij}^l \Big)^2.$$

where $z_{ij}^l$ are i.i.d. standard normal is independent of $\tau_i^l$, thus has a chi-squared distribution with $k-1$ degrees of freedom — hence $\frac{k-1}{2} \cdot s_i^l \tau_i^l$ is gamma distributed with parameters $(\frac{k-1}{2}, 1)$. Since $\frac{\alpha}{\theta} \tau_i^l$ is independent of $s_i^l \tau_i^l$, with $\text{Gamma}(\alpha, 1)$ distribution, we see that $\frac{(k-1)\theta}{2\alpha} s_i^l$ is the ratio of two independent gamma random variables, hence has beta-prime distribution with parameters $\big(\frac{k-1}{2}, \alpha\big)$, so log partial likelihood

$$\ell_{\text{Beta}}(\alpha, \theta; s_\cdot^l) = n\alpha \log \frac{\alpha}{\theta} + n \log \Gamma\left(\alpha + \frac{k-1}{2}\right) - n \log \Gamma(\alpha) + \frac{k-1}{2} \sum_{i=1}^n \log s_i^l - \left(\alpha + \frac{k-1}{2}\right) \sum_{i=1}^n \log\left(s_i^l + \frac{\alpha}{\theta}\right).$$

Note as well that these quantities $(k-1)s_i^l$ should correspond to empirically observed individual variances; hence we will compare these empirical variances (with imputed fractional parts) divided by the normalization factor $2\alpha/(k-1)\theta$ to the beta-prime distribution below as a goodness-of-fit test.

The partial Fisher Information has entries

$$\begin{aligned}
-\frac{\partial^2 \ell}{\partial \alpha^2} &= n\psi_1(\alpha) - n\psi_1\left(\alpha + \frac{k-1}{2}\right) - \frac{n}{\alpha} + \sum_{i=1}^n \frac{2\theta s_i^l + \alpha - (k-1)/2}{(\theta s_i^l + \alpha)^2} \\
-\frac{\partial^2 \ell}{\partial \theta^2} &= -\frac{n\alpha}{\theta^2} + \frac{\alpha}{\theta^2}\left(\alpha + \frac{k-1}{2}\right) \sum_{i=1}^n \frac{2\theta s_i^l + \alpha}{(\theta s_i^l + \alpha)^2} \\
-\frac{\partial^2 \ell}{\partial \theta \partial \alpha} &= \frac{n}{\theta} - \frac{1}{\theta} \sum_{i=1}^n \frac{\alpha^2 + 2\alpha\theta s_i^l + \frac{k-1}{2}\theta s_i^l}{(\theta s_i^l + \alpha)^2}.
\end{aligned}$$

where $\psi_1$ is the trigamma function.

Let $(\hat{\alpha}^l, \hat{\theta}^l)$ be the maximum partial likelihood estimators. Conditioned on $(\tau_i^l)$ we have

$$\bar{y}_{i+} \sim \mathcal{N}\left(m_M, \sigma_M^2 + \frac{1}{4k}\left(\frac{1}{\tau_i^1} + \frac{1}{\tau_i^2}\right)\right),$$
$$\bar{y}_{i-} \sim \mathcal{N}\left(m_\Delta, \sigma_\Delta^2 + \frac{1}{4k}\left(\frac{1}{\tau_i^1} + \frac{1}{\tau_i^2}\right)\right).$$

We would then have MLEs

$$\hat{m}_M = \frac{1}{n}\sum_{i=1}^n \bar{y}_{i+},$$
$$\hat{m}_\Delta = \frac{1}{n}\sum_{i=1}^n \bar{y}_{i-},$$

which are approximately normally distributed, with means $m_M$ and $m_\Delta$ respectively, and conditional on $\tau_i^l$ standard errors

$$\frac{\sigma_M^2}{n} + \frac{1}{4kn^2}\sum_{i=1}^n (\tau_i^1)^{-1} + (\tau_i^2)^{-1} \quad \text{and} \quad \frac{\sigma_\Delta^2}{n} + \frac{1}{4kn^2}\sum_{i=1}^n (\tau_i^1)^{-1} + (\tau_i^2)^{-1},$$

which we may approximate — with error on the order of $n^{-3/2}$ — replacing the mean of $(\tau_i^l)^{-1}$ by its expected value $\theta^l/(\alpha^l - 1)$ to obtain

$$\mathrm{Var}(\hat{m}_M) \approx \frac{\sigma_M^2}{n} + \frac{1}{4kn}\left(\frac{\theta^1}{\alpha^1 - 1} + \frac{\theta^2}{\alpha^2 - 1}\right)$$
$$\mathrm{Var}(\hat{m}_\Delta) \approx \frac{\sigma_\Delta^2}{n} + \frac{1}{4kn}\left(\frac{\theta^1}{\alpha^1 - 1} + \frac{\theta^2}{\alpha^2 - 1}\right)$$

Finally, conditioned on the $\tau_i^l$ we have that the random variables $\bar{y}_{i+}$ are normal with variance

$$\sigma_M^2 + \frac{1}{4k}\left((\tau_i^1)^{-1} + (\tau_i^2)^{-1}\right),$$

so the unconditional variance is the expected value, or

$$\sigma_M^2 + \frac{1}{4k}\left(\frac{\theta_1}{\alpha_1 - 1} + \frac{\theta_2}{\alpha_2 - 1}\right).$$

This yields the estimators

$$\hat{\sigma}_M^2 = \frac{1}{n-1}\sum_{i=1}^n \left(\bar{y}_{i+} - n^{-1}\sum_{i=1}^n \bar{y}_{i+}\right)^2 - \frac{1}{4k}\left(\frac{\hat{\theta}_1}{\hat{\alpha}_1 - 1} + \frac{\hat{\theta}_2}{\hat{\alpha}_2 - 1}\right),$$
$$\hat{\sigma}_\Delta^2 = \frac{1}{n-1}\sum_{i=1}^n \left(\bar{y}_{i-} - n^{-1}\sum_{i=1}^n \bar{y}_{i-}\right)^2 - \frac{1}{4k}\left(\frac{\hat{\theta}_1}{\hat{\alpha}_1 - 1} + \frac{\hat{\theta}_2}{\hat{\alpha}_2 - 1}\right).$$

Using the delta method, and the fact that the correlation between $\hat{\alpha}$ and $\hat{\theta}$ is small, we see that the variance of $\hat{\theta}/(\hat{\alpha} - 1)$ is approximately

$$\frac{\sigma_\theta^2}{(\hat{\alpha} - 1)^2} + \frac{\hat{\theta}^2 \sigma_\alpha^2}{(\hat{\alpha} - 1)^4},$$

where $\sigma_\alpha$ and $\sigma_\theta$ are the standard errors for $\hat{\alpha}$ and $\hat{\theta}$ respectively. Define

$$\hat{\sigma}_{\alpha\theta}^2 := \frac{1}{16k^2}\left(\frac{\hat{\sigma}_{\theta_1}^2}{(\hat{\alpha}_1 - 1)^2} + \frac{(\hat{\theta}_1)^2 \hat{\sigma}_{\alpha_1}}{(\hat{\alpha}_1 - 1)^4} + \frac{\hat{\sigma}_{\theta_2}^2}{(\hat{\alpha}_2 - 1)^2} + \frac{(\hat{\theta}_2)^2 \hat{\sigma}_{\alpha_2}}{(\hat{\alpha}_2 - 1)^4}\right)$$

so the standard errors for $\hat{\sigma}^2_M$ and $\hat{\sigma}^2_\Delta$ are approximately

$$\mathrm{SE}\left(\hat{\sigma}^2_M\right) \approx \left(\frac{2\hat{\sigma}^4_M}{n} + \hat{\sigma}^2_{\alpha\theta}\right)^{1/2},$$
$$\mathrm{SE}\left(\hat{\sigma}^2_\Delta\right) \approx \left(\frac{2\hat{\sigma}^4_\Delta}{n} + \hat{\sigma}^2_{\alpha\theta}\right)^{1/2}.$$

Table 9: Results of estimating parameters from simulated data from the whole population. First column on top is the average parameter estimate from the simulations, second is the true parameter from which the simulations were made, third is the relative error. On bottom are the standard errors for the parameters: True is the theoretically computed standard error, SimAverage is the SD of the simulated parameter estimates, and RelError is the relative error.

| | Systolic | | | Diastolic | | |
|---|---|---|---|---|---|---|
| | SimAverage | True | RelError | SimAverage | True | RelError |
| **Parameter estimates** | | | | | | |
| $m_M$ | 123.3 | 123.3 | 2.24e-05 | 72.33 | 72.31 | 0.0002955 |
| $m_\Delta$ | 1.332 | 1.349 | -0.01244 | 1.114 | 1.1 | 0.01326 |
| $\sigma^2_M$ | 376 | 376.8 | -0.002022 | 104.1 | 104 | 0.001289 |
| $\sigma^2_\Delta$ | 46.36 | 46.42 | -0.001326 | 21.92 | 21.9 | 0.000857 |
| $\alpha_H$ | 2.146 | 2.158 | -0.005432 | 2.365 | 2.322 | 0.01864 |
| $\theta_H$ | 0.1501 | 0.1496 | 0.003854 | 0.1933 | 0.1945 | -0.005856 |
| $\alpha_C$ | 2.576 | 2.569 | 0.002662 | 2.727 | 2.756 | -0.01057 |
| $\theta_C$ | 0.07507 | 0.07515 | -0.001023 | 0.1097 | 0.1093 | 0.003751 |
| **Parameter SE estimates** | | | | | | |
| $m_M$ | 0.1608 | 0.1425 | 0.1282 | 0.08507 | 0.1073 | -0.2073 |
| $m_\Delta$ | 0.05795 | 0.06752 | -0.1417 | 0.04036 | 0.03555 | 0.1353 |
| $\sigma^2_M$ | 4.394 | 4.34 | 0.0125 | 1.218 | 1.511 | -0.1937 |
| $\sigma^2_\Delta$ | 0.551 | 0.6702 | -0.1778 | 0.2651 | 0.2495 | 0.06249 |
| $\alpha_H$ | 0.05593 | 0.0723 | -0.2264 | 0.06586 | 0.0524 | 0.257 |
| $\theta_H$ | 0.002128 | 0.002877 | -0.2605 | 0.002697 | 0.001951 | 0.3823 |
| $\alpha_C$ | 0.07608 | 0.04467 | 0.7034 | 0.08412 | 0.07422 | 0.1334 |
| $\theta_C$ | 0.001033 | 0.001119 | -0.07645 | 0.0015 | 0.001246 | 0.2041 |

Now we compute the combined variance. For a parameter like $\alpha$ we estimate the variance of $\hat{\alpha}$ by

$$\mathrm{Var}(\hat{\alpha}) = \mathbb{E}\left[\mathrm{Var}\left(\hat{\alpha} \mid I\right)\right] + \mathrm{Var}\left(\mathbb{E}\left[\hat{\alpha} \mid I\right]\right).$$

Here $I$ represents the randomly imputed fractional part. We can estimate the first term by averaging the estimated variance (from Fisher Information) over all random imputations. We estimate the second term by the variance of the $\alpha$ estimates over imputations. Note that this is not quite right, since what we really want the variance of is $\alpha_0(I)$ — effectively, the "true" parameter consistent with the imputation. This is a plug-in estimate, as is the Fisher Information estimate of the variance.

##### 2.3.1 Estimates for whole population

The estimates of the empirical Bayes parameters together with their standard errors are given in the column labelled "True" in Table 9.

Finally, we check the variance distribution empirically, to check whether the continuous distribution we have fit for individual variances describes the true distribution of variances in the population reasonably well. The first thing we do is to compare the empirical variances (with fractional parts imputed according to the observed proportions for the unequal digit preference, as discussed in section 1.3.1) to the theoretical beta-prime distribution. To match the standard distribution, the variances are normalized by being divided by the factor $\alpha/\theta$. We show histograms of these "unrounded" empirical variances and the theoretical beta-prime distribution in Figure 9. Note that the distribution has a very long tail, and we have truncated about 2% of the data to make the figures more readable.

In Figure 10 we show essentially the same data in the form of Q–Q plots. Here we have extended the plot far out into the tails of the distribution, including values in the range $[0, 10]$, covering around 99.7% of the data. We generate data from the inferred model that mimic the true data, with three systolic and three diastolic BP measures per person. As before, we impute the fractional parts to the real data. This gives us a set of true variances and a set of simulated variances, which we hope will have approximately the same distribution. We see some deviation here, but it is slight, and quite deep into the tails. Furthermore, the deviation is in the direction of the simulated data having slightly fatter tails than the true data, which is the direction we would wish to err in for the sake of making conservative inferences.

The estimates of the empirical Bayes parameters together with their standard errors are given in the column labelled "True" in Table 9. These parameters (and SEs) are accompanied by the results of 10 estimates of data simulated from the model with the parameters inferred from the data, and then fitted by the same procedure. Note that the errors for the estimates are consistent with the stated standard errors ($\pm\sqrt{\mathrm{SE}}$), and the relative errors for the SE are small, confirming that the estimation procedure is reliable.

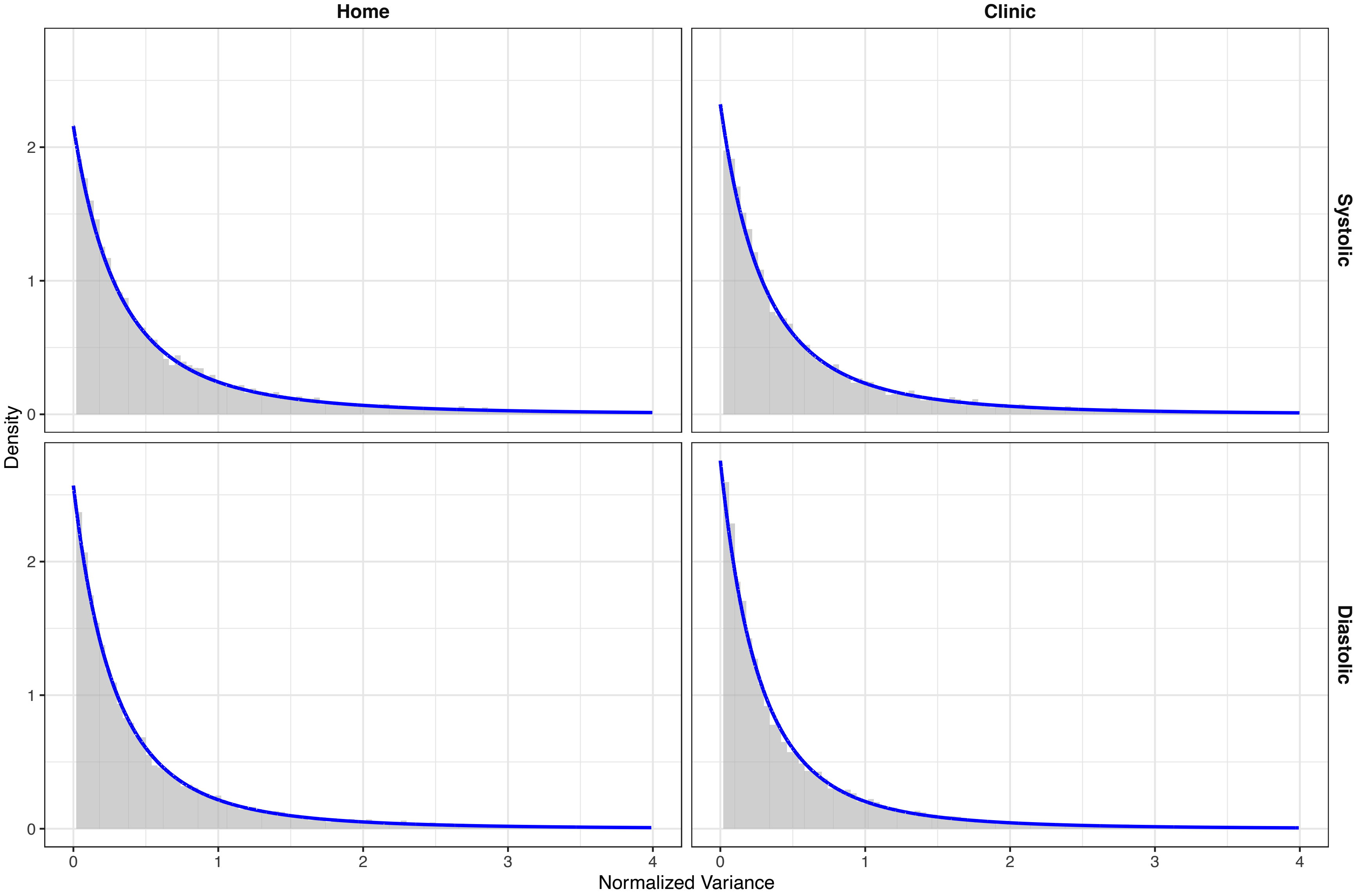


Figure 9: Comparison of the distribution of empirical variances, normalized by dividing by $\beta = \alpha/\theta$, to the fitted beta-prime distribution.

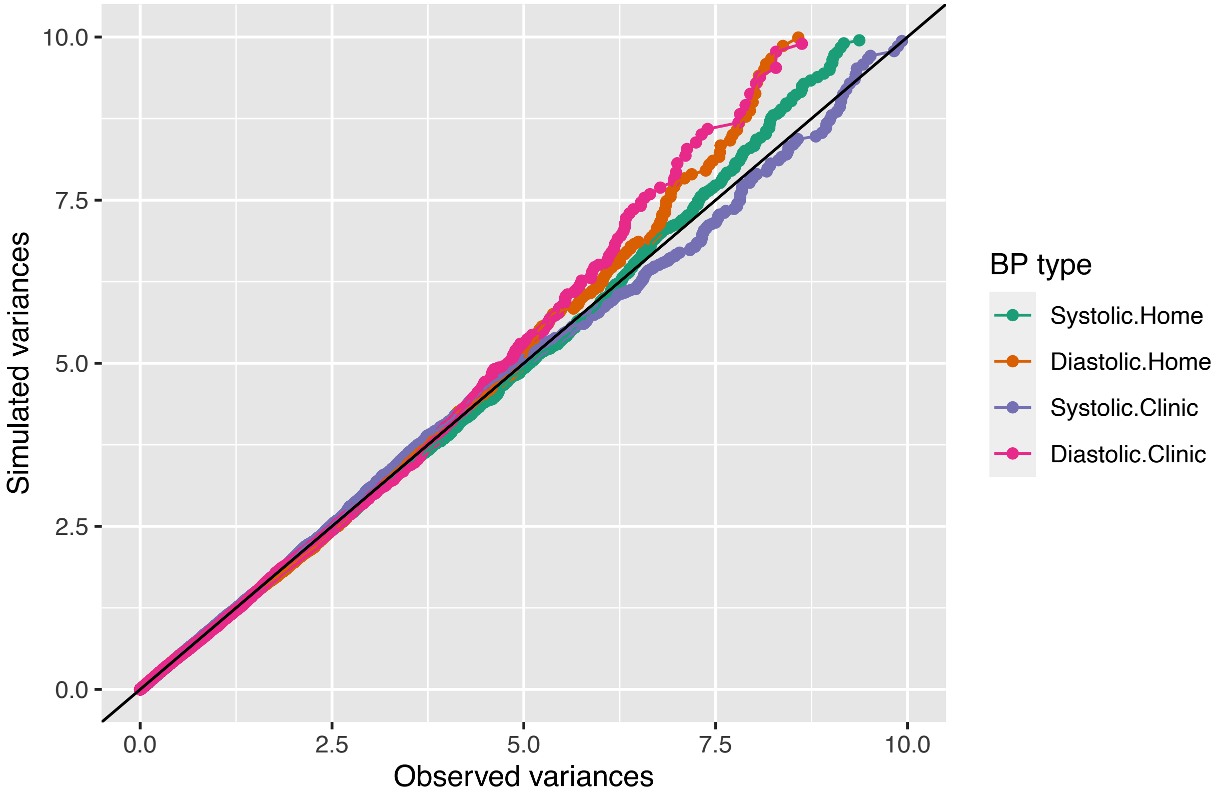


Figure 10: Q–Q plots of the variances of the observed data with imputed fractional parts (x-axis) against the variances of the simulated data (y-axis).

#### 2.4 Hamiltonian Monte Carlo (HMC)

The model, as described in the article, is a Bayesian hierarchical model. In order to parameterize such an intricate model, traditional Maximum Likelihood Estimation methods can no longer be applied. Therefore, we apply the Hamiltonian Monte Carlo (HMC) method. HMC is a form of Markov Chain Monte Carlo methods, which samples potential parameter space values of the model, then calculates directly the likelihood function based on that choice of parameters. The derivative of the likelihood function, $\phi$, guides parameter space exploration in $\theta$ towards the modal value of the joint posterior distribution. This method is ideal for complicated, non-Gaussian distribution forms. The three steps of HMC are:

1. Draw a sample of the derivative $\phi$ using the posterior distribution of $\phi$, which is the same as its prior.
2. Update the values of $\theta^*$ and $\phi^*$ using

$$\theta^* \leftarrow \theta + \epsilon M^{-1}\phi, \tag{11}$$

   and

$$\phi \leftarrow \phi + \epsilon \frac{1}{2} \frac{\mathrm{d}\log\{p(\theta|y)\}}{\mathrm{d}\theta}, \tag{12}$$

   where $M$ is the Jacobian of the parameters. This can be set to a diagonal matrix for no correlation between parameters, and is updated pointwise throughout the calculation. This is the leapfrog method, whereby $\epsilon$ dictates the scale size of the step to ensure convergence on the correct point is made, and L is the number of steps to be 'leaped'.
3. Compute the rejection parameter:

$$r = \frac{p(\theta^*|y)p(\phi^*)}{p(\theta^{t-1}|y)p(\phi^{t-1})} \tag{13}$$

4. Set $\theta^t$ to $\theta^*$ with probability $\min\{1, r\}$, or otherwise keep $\theta^{t-1}$.

The tuning parameters $\epsilon$ and L should be chosen according to a desired acceptance rate. The No-U-Turn Sampler of Stan automates the calculation of these tuning parameters. A more detailed overview of HMC and the NUTS algorithm integrated into the Stan package, see (Hoffman, Gelman, et al. 2014).

##### 2.4.1 Centering the Linear Predictor

During the MCMC simulations, the centering values play a non-negligible role in shaping the model parameterization. If the centering parameters are held constant throughout all of the MCMC simulations, then the equation $\sum_i^N \exp\left(\beta \cdot (X - \hat{X})\right) = 0$ is no longer guaranteed. However, automatically defining the centering values based on the model parameters sampled at the current MCMC iteration is not advisable as it can lead to poor parameter convergence. This is because it modifies the likelihood function at every MCMC iteration. Therefore, we iterate the MCMC algorithm multiple times. At every iteration, we recalculate the centering parameters to satisfy the requirement that the average of the linear predictor term going to zero, based on the posterior distributions of the previous MCMC simulation. This iteration is carried out until the centering parameters converge. Convergence is defined by optimising on two factors. The first is that the sum of the linear predictor term across all MCMC samples needs to tend to negligible values (we define this as the average difference being less than $10^{-7}$), see figure 11. The second convergence criteria is that the average Root Mean-Squared Error (RMSE) of the model predictions on the survival outcomes in the MCMC simulations needs to also decrease towards zero, see figure 11 (top). For the second criteria, we stopped the simulations when either the difference in the RMSE stopped decreasing (below a threshold of 1%), or the RMSE value was less than 20, see figure 11 (bottom). Illustration of the convergence is shown in figure 11.

#### 2.5 Code Description

The code will be made available, but detailed references have been removed to preserve anonymity for the review process.
