## Appendix C for "Short-term and Mid-term Blood Pressure Variability and long-term Mortality: evidence from the Third National Health and Nutrition Examination Study"

### 3 Appendix C – Further Results

In this section, we add some additional detail to the results section covered in the article. Extra information is given to explain how convergence of the simulations was ensured, and to also include more visualisations of the converged model parameterizations. The authors feel that this is particularly useful to provide confidence in the model parameterization and the predictions.

#### 3.1 Convergence of Simulations

Convergence of the simulations required to parameterize the model presented in this work is required for the MCMC simulations performed by Stan, as well as convergence in the centering values that requires repeating the Stan calculations several times. Convergence of the latter is shown in figure 11. The upper plot in figure 11 illustrates convergence in the average Root Mean-Squared Error (RMSE) of the model predictions on the survival outcomes in the MCMC simulations. The lower plot in figure 11 illustrates convergence in the average sum of the linear predictor terms over all MCMC chain iterations.

With respect to convergence of the MCMC simulations, defining convergence first involves discarding the burn-in period of the simulations. When the time-evolution marker chain has a large number of samples, sequence thinning is used to reduce the amount of data storage - after convergence, take only the kth value of the simulations (after having discarded the burn-in phase values) and discard the rest. One measure of convergence is to bin similar markers and check that for each bin, the variation of the individual marker movement over a few time steps is larger than the variation of the ensemble markers in-between one-another. Other methods of convergence are stationarity and mixing. The former occurs by ensuring that the gradients of movements in the chains in time are in the same direction, the latter ensures that the amplitude of the movements in the chains are similar. To calculate the mixing and stationarity, one can do the following:

- Take the proposedly converged marker population, where there are N markers in total each of index length $\tau$ (thus of total physical time quantity $t\tau$). Split it k times, where k is a common denominator of $\tau$.
- Now you have $kN$ MCMC chains each of length $|\tau/k|$
- For the marker $\psi_{ij}$ with i and j the chain length (time) and marker number indices respectively, then the mean marker value over the chain length (time) is

$$\bar{\psi}_{|,j} = \frac{k}{\tau}\sum_{i=1}^{\tau/k}\psi_{ij} \tag{14}$$

  and the total average quantity of $\psi$ over all markers, over all chain lengths is therefore

$$\bar{\psi}_{||} = \frac{1}{kN}\sum_{j=1}^{kN}\bar{\psi}_{|j} \tag{15}$$

- Stationarity: compare the inter-marker variance (between sequence B):

$$B = \frac{\tau}{k(kN-1)}\sum_{j=1}^{kN}(\bar{\psi}_{|,j} - \bar{\psi}_{||})^2 \tag{16}$$

- Mixing: compare the variance along each markers chain length (within-sequence W):

$$W = \frac{1}{n(\tau-k)}\sum_{j=1}^{kN}\sum_{i=1}^{\tau/k}(\psi_{i,j} - \bar{\psi}_{|j})^2 \tag{17}$$

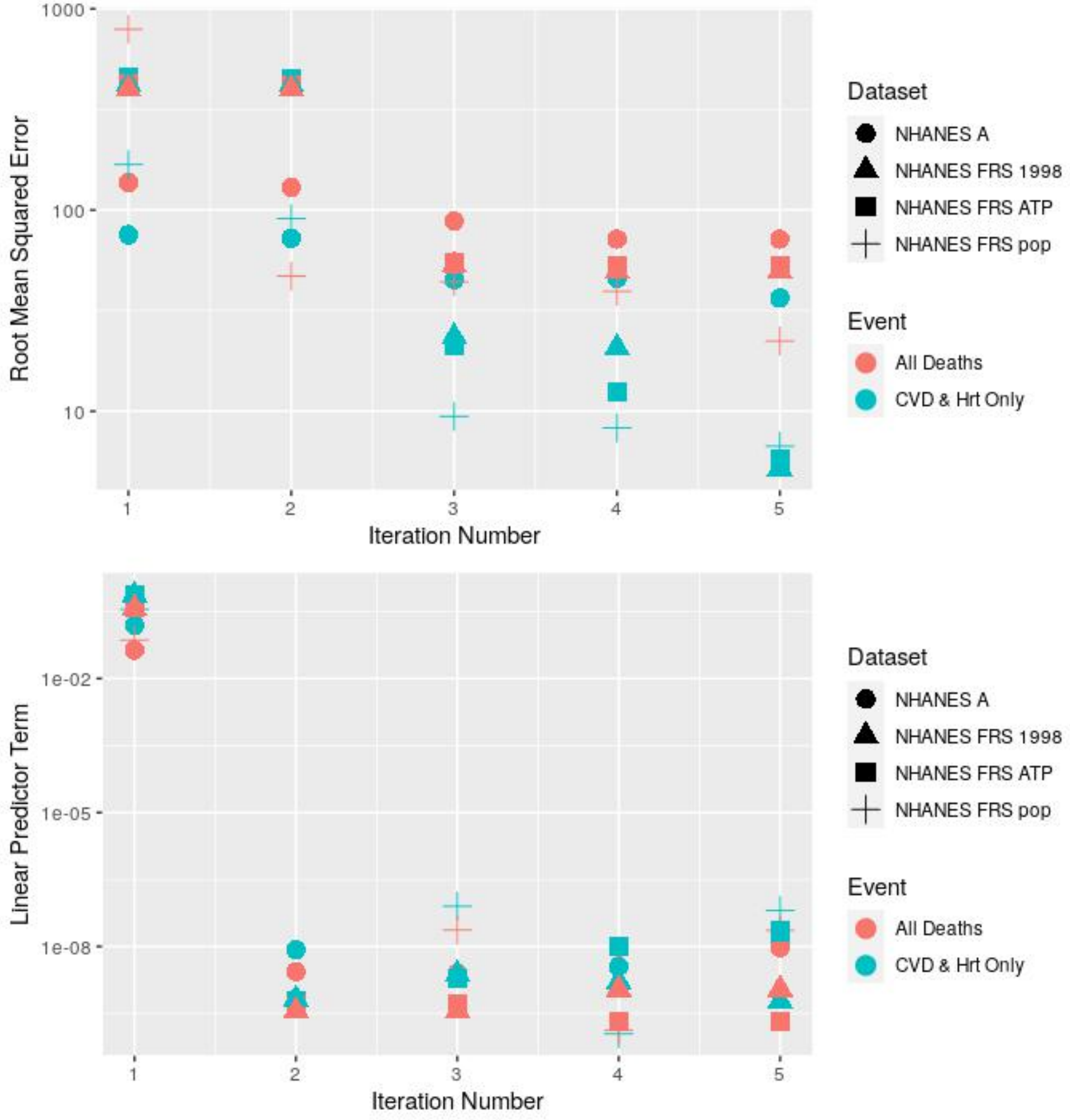


Figure 11: Illustration of the convergence of the centering parameters of the model.

- Therefore, to estimate the marginal posterior variance of $p(\psi|y)$, then we use a weighted average

$$\hat{\text{Var}}^{+}(\psi|y) = \frac{\tau - k}{N} W + \frac{1}{Nk} B \tag{18}$$

Note that this quantity overestimates the marginal posterior variance, but it is unbiased under stationarity: this can be used to infer convergence. When the varation in

$$\hat{R} = \sqrt{\frac{\hat{\text{Var}}^{+}(\psi|y)}{W}} \tag{19}$$

should approach close to 1 for converged simulations.

Another convergence parameter is the number of effective independent marker draws. Upon convergence, the time evolution of each marker should be uncorrelated and independent to previous time steps. To find the average time-correlation over all particles, we use the variogram $V_t$:

$$V_t = \frac{1}{Nk(\tau/k - \tilde{t})} \sum_{j=1}^{kN} \sum_{i=1}^{\tau/k} (\psi_{i,j} - \psi_{i-\tilde{t},j})^2, \tag{20}$$

where $\tilde{t} \in 1, 2, ..., \tau/k$ is a time index. Then we get the time-correlations:

$$\hat{\rho}_t = 1 - \frac{V_t}{2\hat{\text{Var}}^{+}} \tag{21}$$

This comes from the expectation of the variance $E[(\psi_i - \psi_{i-t})^2] = 2(1 - \rho_t)\text{Var}(\psi)$. This can be used to infer the effective number of independent marker draws:

$$\hat{n}_{eff} = \frac{mn}{1 + 2\sum_{\tilde{t}=1}^{T} \hat{\rho}_t} \tag{22}$$

Where T is the index at which the sum of the autocorrelation estimates $\hat{\rho}_{t'} + \hat{\rho}_{t'+1}$ is negative. As a general guide, we should have $\hat{n}_{eff} \sim 10N/k$ effective independent marker draws and that $\hat{R} \to 1 \sim 1.1$. In this research, we continued running the MCMC simulations until these two criteria were met (and went beyond: $\hat{R} < 1.05$ for all parameters in all models and that $\hat{n}_{eff} > 750$ for all parameters in all models).

#### 3.2 Results - Model Parameterization

We remind the reader of the list of numbers of the different models explored in this research, provided in the list found in section 'Proposed Models'. The authors will use the numbers in the list, referred to as the run number, in the following plots. One of the most important set of parameters of the model is the vector $\beta$ of covariates in the Cox' proportional hazards model. When the $\beta$ vector is normalised, the larger (in absolute terms) the value of $\beta$, the larger the correlation between that specific covariate and the risk of mortality. Positive values of $\beta$ imply a higher risk of mortality, and the inverse for negative values of $\beta$. As we can see from the violin plots of the MCMC posterior samples of the $\beta$ parameters in figure 12, the parameter that correlated the highest with both the mortality risk of CVD and for all mortalities, in absolute terms, was the 1998 version of the FRS score, shown in the top-right plot under run numbers 7 and 8. The FRS-1998 score correlated, on average over all the MCMC iterations, approximately 25% more with mortality risk of CVD than the (more recently developed) FRS ATP III score. A similar, but slightly weaker, correlation was found between the two FRS scores for all mortality-based risk. The middle-left plot in figure 12 shows that the mean diastolic blood pressure acts to decrease mortality risk. Finally, the influence of the longer-term difference in the mean blood pressure, displayed in the top-left and top-middle plots of figure 12, is also shown to increase mortality risk across all run numbers. The influence of the blood-pressure variability on mortality is illustrated to not be consistent across simulations, whereby the statistical significance of the effect is lower than for the other parameters in the linear predictor term.

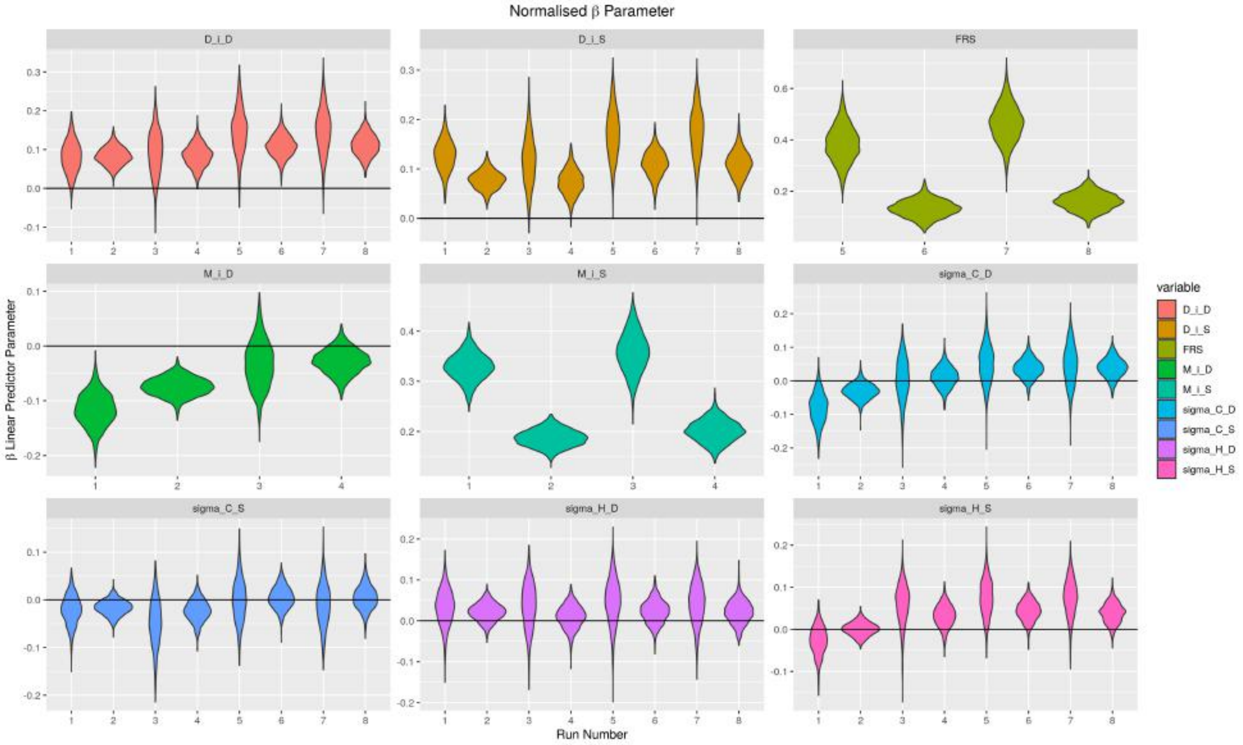


Figure 12: Violin plots of the normalised $\beta$ parameters of the different models.

With respect to the time-independent Gompertz parameter, denoted $B$ in this article, the results between all models that simulate CVD mortality risk, and all the models that simulation all-cause mortality risk are consistent with one-another. This is illustrated by the similarity between plots on the left hand side and the right hand side of figure 13. The consistency appears across sex assigned at birth and race.

Figure 14 reflects the same level of consistency for the Gompertz parameter that influences the temporal evolution of the mortality risk. It is worth noting that both figures 13 and 14 have inverse trends between the values of B and theta for each demographic group. This makes it difficult to imagine, based on these two plots, what the mortality risk is at different ages across demographics, yet it is evident that the form of the change in the mortality risk curve in time is different for each demographic group. Women are observed to have lower initial values of risk, but mortality risk later in life begins to increase much faster than for men. Additionally, hispanic populations are shown to have a larger initial mortality risk than black populations who are shown to have a larger initial mortality risk than white populations in the USA. However, mortality risk increases at a faster rate for white populations than for black populations, for which it increases faster than hispanic populations in the USA. For ease of comparison, we also present here tables of the mean and standard deviation values of the time dependent and independent Gompertz parameters in tables 11 to 13.

##### 3.3 Results - Model Performance

To measure the performance of the model to predict the survival outcome of individuals in the population, figure 15 shows, ordered by individual age, the cumulative hazard $H(t)$ predicted against the cumulative number of deaths in the populations, for each model explored in this research. Each model is shown to predict survival outcomes reliably, across the entire age range of the population.

A common metric that is used to evaluate the performance of models such as presented in this article is called the Receiver Operating Characteristic (ROC) curve. With continuous predictor values such as cumulative hazard $H(T_i)$, a threshold can be defined whereby any individual who has a cumulative risk larger than the threshold $H(T_i) > \epsilon$ is predicted to die. The ratio of the number individuals that were predicted to die

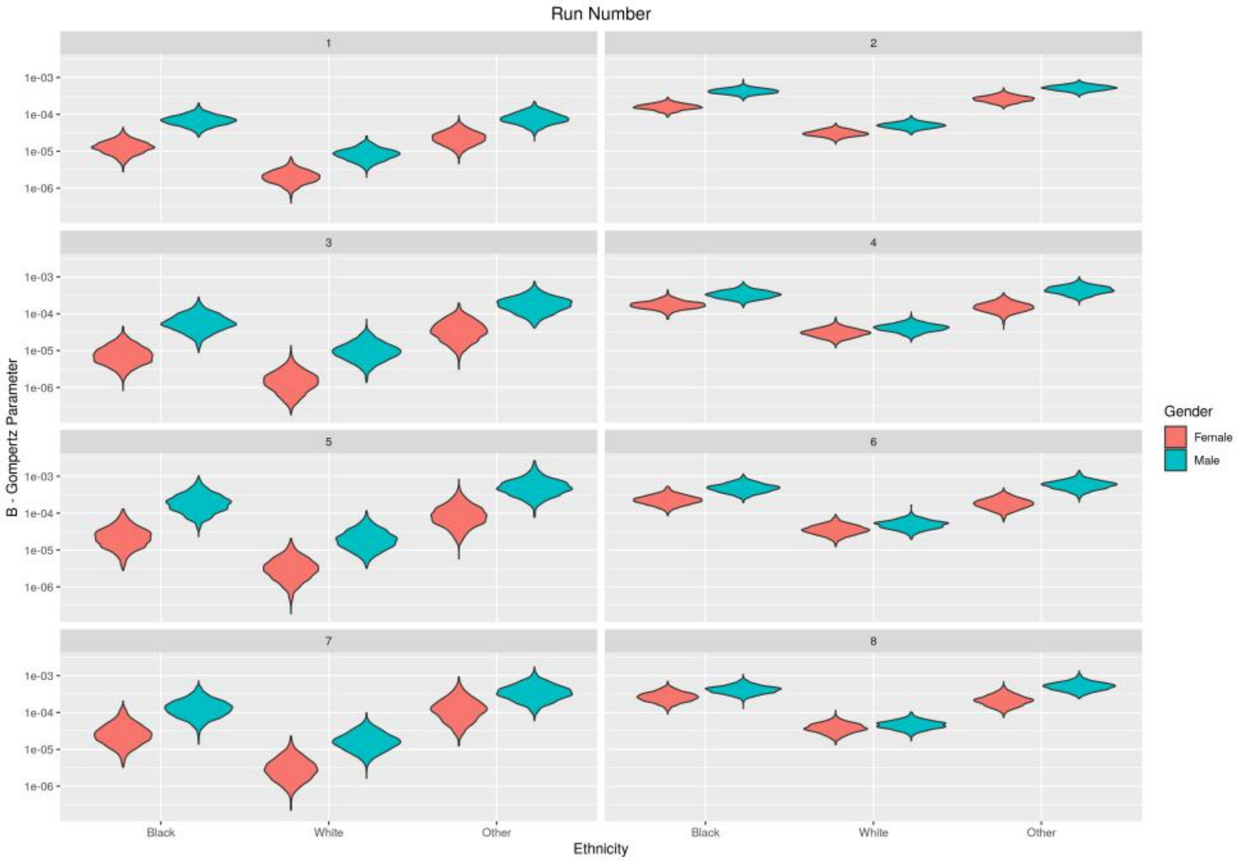


Figure 13: Violin plots of the normalised B parameter (from the Gompertz equation) of the different models.

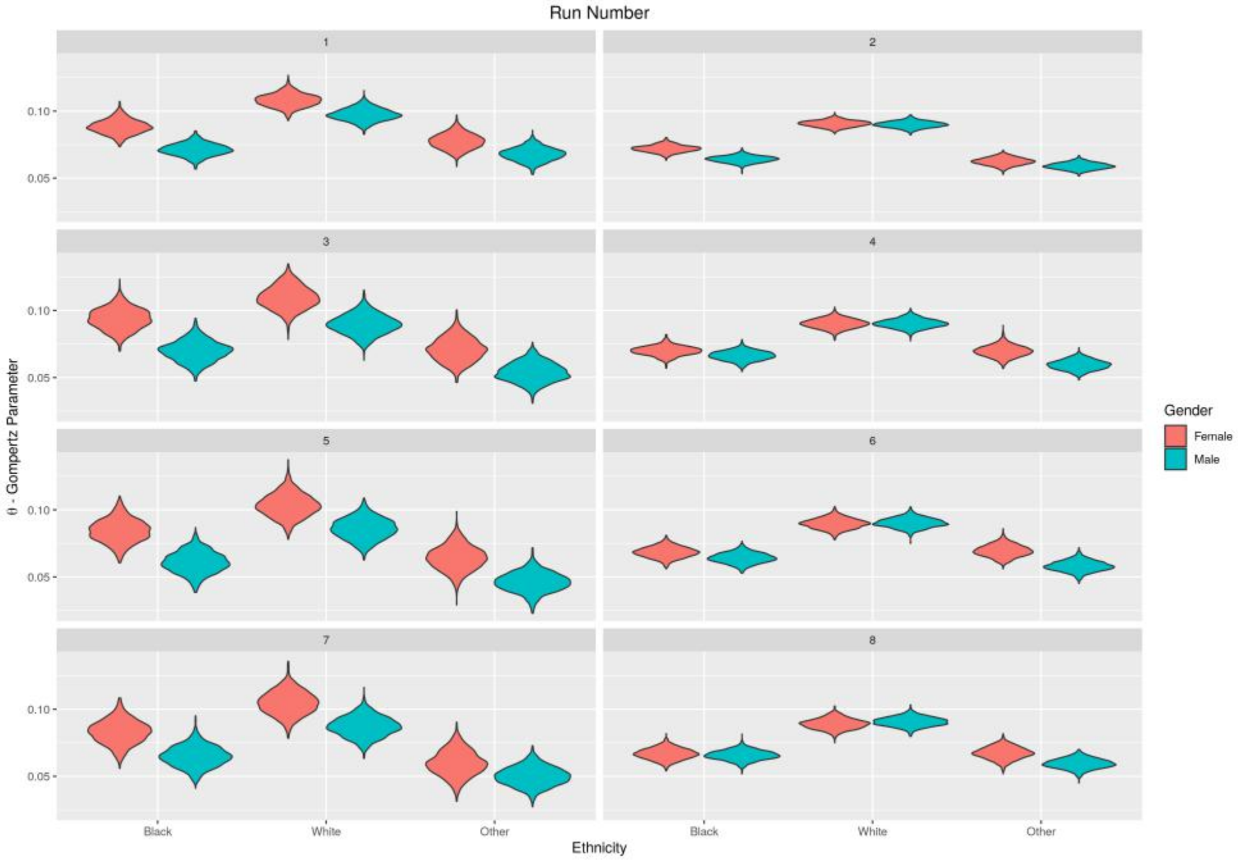


Figure 14: Violin plots of the normalised $\theta$ parameter (from the Gompertz equation) of the different models.

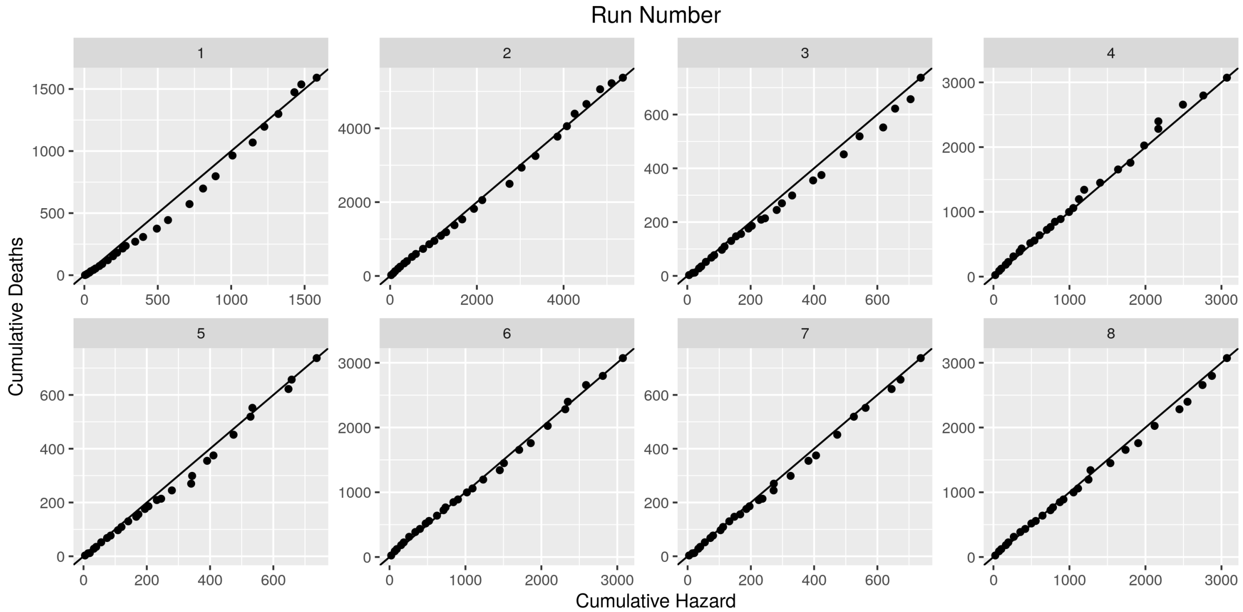


Figure 15: Predicted cumulative hazard against cumulative number of deaths in the population, ordered by the age of the individual.

Table 10: Summary of the posterior estimate of the blood pressure distribution.

| | Systolic | | Diastolic | |
|---|---|---|---|---|
| **Full population** | | | | |
| Overall Mean | 125.4 | 19.5 | 74.3 | 10.3 |
| $\lvert\Delta\rvert = \lvert\text{Home} - \text{Clinic}\rvert/2$ | 5.24 | 4.83 | 3.90 | 3.14 |
| Home Stand Dev | 2.74 | 2.05 | 2.34 | 1.75 |
| Clinic Stand Dev | 3.78 | 2.61 | 3.08 | 2.08 |
| **FRS population** | | | | |
| Overall Mean | 125.9 | 18.3 | 76.4 | 10.0 |
| $\lvert\Delta\rvert = \lvert\text{Home} - \text{Clinic}\rvert/2$ | 5.23 | 4.71 | 3.84 | 3.10 |
| Home Stand Dev | 2.78 | 2.06 | 2.28 | 1.71 |
| Clinic Stand Dev | 3.78 | 2.52 | 2.92 | 1.97 |

Table 11: Parameters for survival model, NHANES III, Full population, using the systolic and diastolic mean model.

| Sex | Race/Ethnicity | B–Mean | B–SD | $\theta$-Mean | $\theta$–SD |
|---|---|---|---|---|---|
| **All-Cause Mortality** | | | | | |
| Female | Black | 1.62e-04 | 3.14e-05 | 0.0720 | 0.00264 |
| Female | White | 3.12e-05 | 6.20e-06 | 0.0906 | 0.00238 |
| Female | Mexican | 2.66e-04 | 5.45e-05 | 0.0625 | 0.00279 |
| Male | Black | 4.29e-04 | 7.54e-05 | 0.0641 | 0.00243 |
| Male | White | 5.08e-05 | 9.70e-06 | 0.0900 | 0.00236 |
| Male | Mexican | 5.27e-04 | 8.88e-05 | 0.0590 | 0.00245 |
| **CVD Mortality** | | | | | |
| Female | Black | 1.36e-05 | 5.70e-06 | 0.0883 | 0.00534 |
| Female | White | 2.20e-06 | 9.00e-07 | 0.1080 | 0.00488 |
| Female | Mexican | 2.44e-05 | 1.07e-05 | 0.0775 | 0.00565 |
| Male | Black | 7.20e-05 | 2.49e-05 | 0.0712 | 0.00459 |
| Male | White | 8.70e-06 | 3.40e-06 | 0.0973 | 0.00466 |
| Male | Mexican | 8.17e-05 | 3.00e-05 | 0.0678 | 0.00498 |

compared to the total number who die corresponds is referred to as the True Positive Ratio (TPR)

$$TPR(\epsilon) = \frac{\sum_i \left(\mathbb{I}(H(T_i) > \epsilon \ \ \& \ \ \delta_i = 1)\right)}{\sum_i \left(\mathbb{I}(\delta_i = 1)\right)}. \tag{23}$$

Note that TPR is also referred to as the recall or sensitivity. Conversely, the ratio of the number of individuals predicted to die but survive compared to the total number of individuals that survived is referred to as the False Positive Ratio (FPR)

$$FPR(\epsilon) = \frac{\sum_i \left(\mathbb{I}(H(T_i) > \epsilon \ \ \& \ \ \delta_i = 0)\right)}{\sum_i \left(\mathbb{I}(\delta_i = 0)\right)}. \tag{24}$$

Note that the FPR is also referred to as $1-$specificity. The ROC curve is produced by varying the threshold value that is then used to calculate both the TPR and FPR, and plotting them against one another. The area under this curve is a metric that indicates performance of the model to predict survival outcomes. AUROC=1 implies perfect predictions and AUROC=0.5 implies the contrary. However, our model is formulated such that the variables age and time since starting the survey both form part of Cox's proportional hazard. Furthermore, the Gompertz model is stratified by demographic group. Therefore, in this work, we present a modified ROC curve, which calculates the individuals cumulative hazard at a given time since the start of

Table 12: Parameters for survival model, NHANES III, FRS-population only, using the systolic and diastolic mean model.

| Sex | Race/Ethnicity | B–Mean | B–SD | $\theta$-Mean | $\theta$–SD |
|---|---|---|---|---|---|
| **All-Cause Mortality** | | | | | |
| Female | Black | 1.77e-04 | 4.66e-05 | 0.0699 | 0.00359 |
| Female | White | 3.12e-05 | 8.80e-06 | 0.0902 | 0.00355 |
| Female | Mexican | 1.56e-04 | 4.83e-05 | 0.0696 | 0.00427 |
| Male | Black | 3.41e-04 | 8.54e-05 | 0.0662 | 0.00352 |
| Male | White | 4.27e-05 | 1.10e-05 | 0.0902 | 0.00335 |
| Male | Mexican | 4.51e-04 | 1.17e-04 | 0.0594 | 0.00369 |
| **CVD Mortality** | | | | | |
| Female | Black | 8.60e-06 | 5.40e-06 | 0.0937 | 0.00795 |
| Female | White | 1.70e-06 | 1.30e-06 | 0.1100 | 0.00844 |
| Female | Mexican | 4.06e-05 | 2.65e-05 | 0.0708 | 0.00879 |
| Male | Black | 6.36e-05 | 3.53e-05 | 0.0697 | 0.00765 |
| Male | White | 1.08e-05 | 6.50e-06 | 0.0896 | 0.00737 |
| Male | Mexican | 1.94e-04 | 9.69e-05 | 0.0539 | 0.00709 |

Table 13: Parameters for survival model, NHANES III, FRS-population only, using the 1998 FRS-based model.

| Sex | Race/Ethnicity | B–Mean | B–SD | $\theta$-Mean | $\theta$–SD |
|---|---|---|---|---|---|
| **All-Cause Mortality** | | | | | |
| Female | Black | 2.34e-04 | 7.08e-05 | 0.0684 | 0.00404 |
| Female | White | 3.66e-05 | 1.16e-05 | 0.0897 | 0.00392 |
| Female | Mexican | 1.88e-04 | 6.25e-05 | 0.0694 | 0.00442 |
| Male | Black | 4.94e-04 | 1.42e-04 | 0.0640 | 0.00384 |
| Male | White | 5.01e-05 | 1.46e-05 | 0.0903 | 0.00365 |
| Male | Mexican | 6.00e-04 | 1.74e-04 | 0.0580 | 0.00391 |
| **CVD Mortality** | | | | | |
| Female | Black | 2.62e-05 | 1.82e-05 | 0.0843 | 0.00859 |
| Female | White | 3.70e-06 | 2.70e-06 | 0.1030 | 0.00861 |
| Female | Mexican | 1.02e-04 | 7.46e-05 | 0.0641 | 0.00896 |
| Male | Black | 2.14e-04 | 1.34e-04 | 0.0608 | 0.00793 |
| Male | White | 2.22e-05 | 1.46e-05 | 0.0859 | 0.00778 |
| Male | Mexican | 5.58e-04 | 3.25e-04 | 0.0461 | 0.00720 |

the survey, $T_{surv} \in 5, 10, 15$ years, and calculates whether the model correctly predicted an event to occur before or after this time. Note that to do this, we split the ROC population by ages: 45-64 and 65-84. The modified TPR is then calculated via:

$$TPR(\epsilon) = \frac{\sum_i \left(\mathbb{I}(\delta_i = 1 \;\; \& \;\; H(T_i) \geq \epsilon \;\; \& \;\; T_i < T_{surv})\right)}{\sum_i \left(\mathbb{I}(\delta_i = 1 \;\; \& \;\; T_i < T_{surv})\right)}, \tag{25}$$

and the modified FPR:

$$FPR(\epsilon) = \frac{\sum_i \left(\mathbb{I}(H(T_i) \geq \epsilon \;\; \& \;\; T_i \geq T_{surv})\right)}{\sum_i \left(\mathbb{I}(T_i \geq T_{surv})\right)}. \tag{26}$$

Before presenting any ROC or AUC values, we first present the density distributions of the median posterior systolic $\Delta$ values for all individuals, split by demographic.

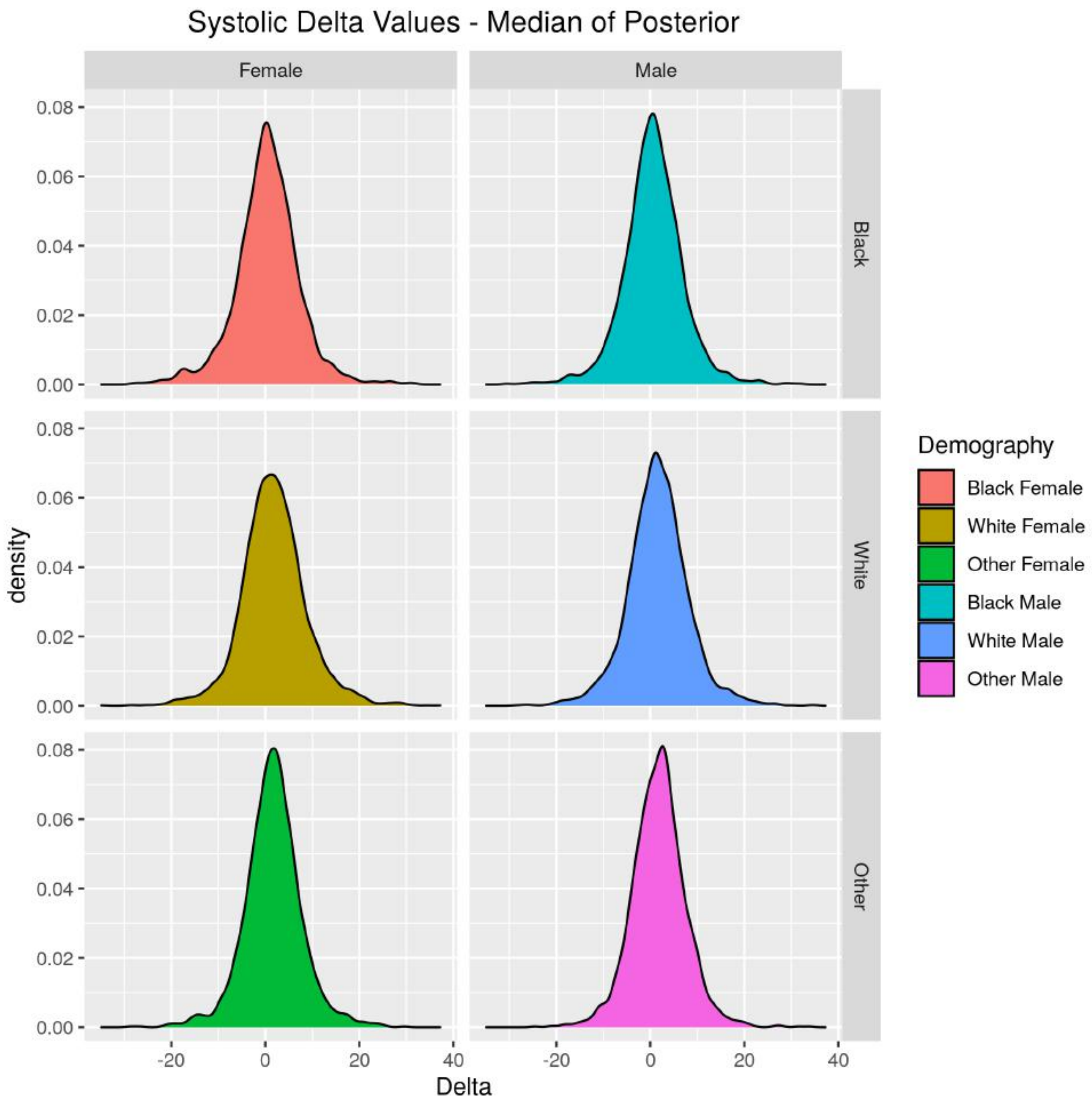


Figure 16: Density of the (median posterior) systolic $\Delta$ values, per demographic.

Using Welch's ANOVA test, we calculated that $p < 1 \times 10^{-9}$ for all demographics, including when split between the male and female populations. Figures 17 and 18 show the ROC curves (including the AUC values) of the model, the former using the mean systolic and diastolic blood pressure as covariates in the linear predictor term (in the Cox's proportional hazards component) and the latter using the FRS value instead. By making predictions of the 5, 10 and 15 year survival between the middle aged and old aged sub-groups, for the three different mortality causes, we start to build a picture of the performance of the model. For figure 17, we notice that the AUC value of the predictions for the middle aged compared to the older aged population is higher, independent of the survival year prediction or the different mortality causes. The highest AUC is for the 45-64 year old population with a focus on CVD and heart attack-related mortality, for all three survival year periods. We also note that the TPR seems to start increasing at a faster rate for the population aged 45-64 than the 65-84 group, implying that it is possible to choose a threshold level, $\epsilon$, for the survival predictions that could correctly identify people at risk without incorrectly predicting as many people to be at risk of mortality as for the group aged 65-84. The results also reflect that the influence of choosing a 5, 10 or 15 year prediction period does not seem to significantly influence the results. Figure 18 displays similar results to the mean systolic and diastolic model when using the FRS value instead, with the main difference that the predictions of the middle aged group for CVD and heart attack-related mortality for 5 year survival seems to be lower than the equivalent in the older group or as compared to the mean blood pressure model. This is caused by a reduced mortality before 5 years for the middle aged population that had their FRS value calculated, where 36, 83 and 213 CVD and heart attack-related deaths occurred before 5, 10 and 15 years in this sub-group, respectively. This can be compared to 141, 356 and 952 all-cause deaths in this same sub-group (red-curve in figure 18 and 17). Alternatively, when compared to the full-population (not just those who had the FRS value), the CVD and heart attack-related deaths for the middle aged population are 55, 115 and 282 over the 5, 10 and 15 year range, respectively.

Comparison of the ROC curves and AUC values is also presented for the different demographic groups, see figure 19. This figure shows the differences in the prediction performance (w.r.t. the ROC and AUC values) of the full-population model for the 45-64 age population for their 10 year survival outcome, using the mean systolic and diastolic blood pressure model. This plot illustrates that potentially only the all-cause mortality has enough outcomes in each demographic group to separate the ROC curves. The model seems to most accurately predict the 10 year survival outcome of the black and other ethnic groups, as well as the white female demographic as compared to the black female, other female and white male population. To provide insight into this, we also provide the frequency table of deaths for each demographic group, mortality cause and survival year, see table 14 and 15.

To finalise the section on the performance of the models using ROC and AUC values, we present a series of figures that provide ROC curves and AUC values for different linear predictor (Cox's proportional hazards model) covariate configurations. By setting the covariate-specific $\beta$ parameter values to zero, we can measure the additional prediction performance that adding different covariates provides to the model. In figures 20 to 25, we present three main formulations: using only the systolic and diastolic $\Delta$ terms, using the mean systolic and diastolic (figures 20-22) or the FRS value (figures 23-25) terms as well as the systolic and diastolic $\Delta$ terms (figures 20-25) and, finally, using the systolic mean (figures 20-22) or FRS value only (figures 23-25). Figures 20, 21 and 22 apply to the full-population with the models trained on CVD and heart-attack related mortality, all-cause, and other mortality, respectively. Figures 23, 24 and 25 apply to the population with an FRS value, with the models trained on CVD and heart-attack related mortality, all-cause, and other mortality, respectively. The first thing to note as a commonality between all these different figures is that the use of the long-term variability, $\Delta$, in the model has comparable performance with that of using the systolic mean or FRS values only. Additionally, where the number of deaths permits for prediction, the use of both the $\Delta$ and mean/FRS values results in higher AUC values than using models that use one or the other. Finally, the use of the FRS value consistently under-performs the mean diastolic and systolic blood pressure-based model.

#### 3.4 Results - Exploring $\Delta$ Directionality

This section of the appendix is to explore whether the directionality of the difference in clinic-home blood pressure (represented through the non-absolute value of the $\Delta$ covariate) may have an influence on the

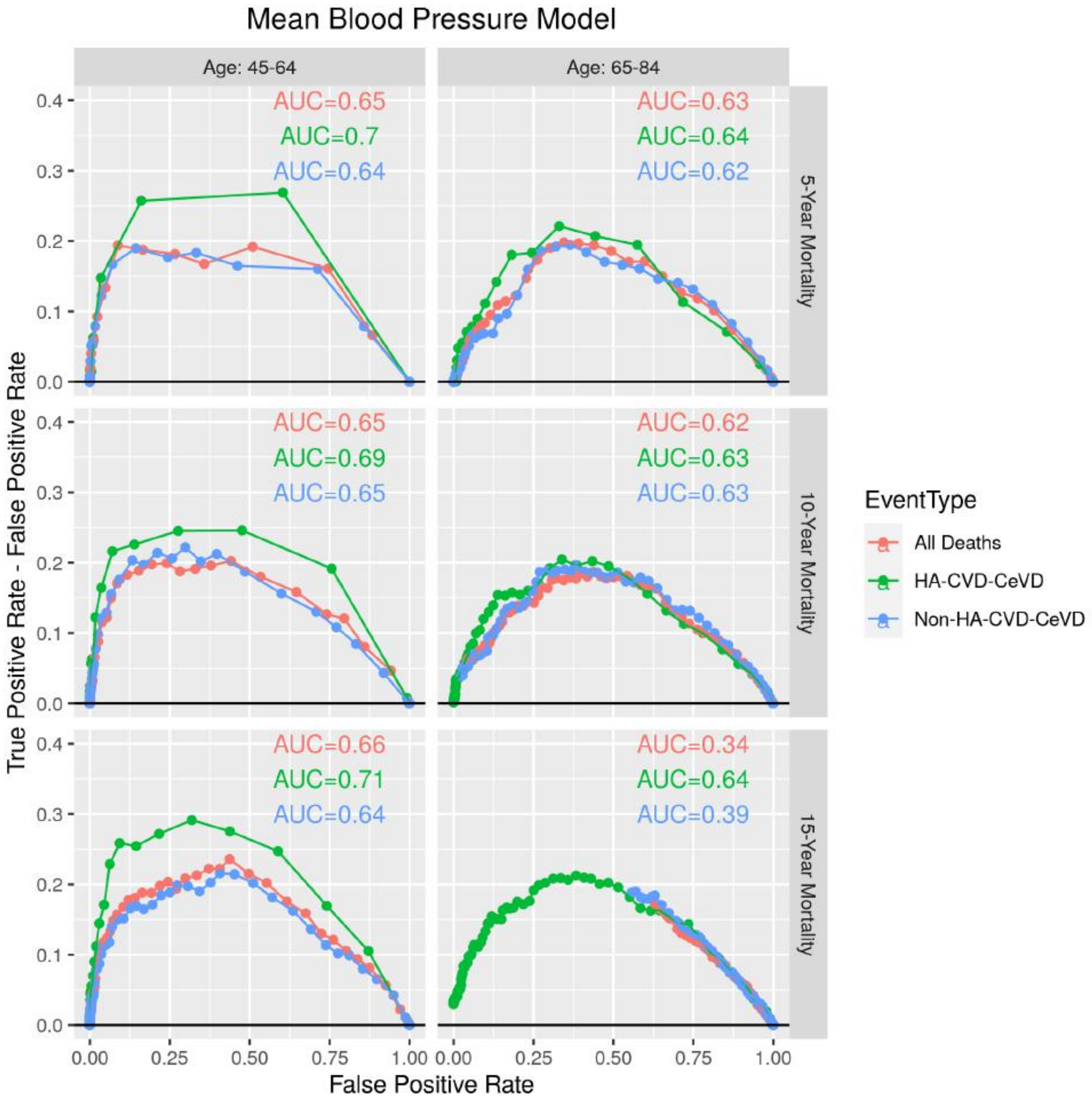


Figure 17: ROC curves for the model that used mean systolic and diastolic blood pressure as covariates in the linear predictor, stratified by the event type (cause of mortality). The columns split two groups in the population: those who start the survey aged between 45 to 64 and 65-84 years old. The rows split the model predictions between 5, 10 and 2 year survival.

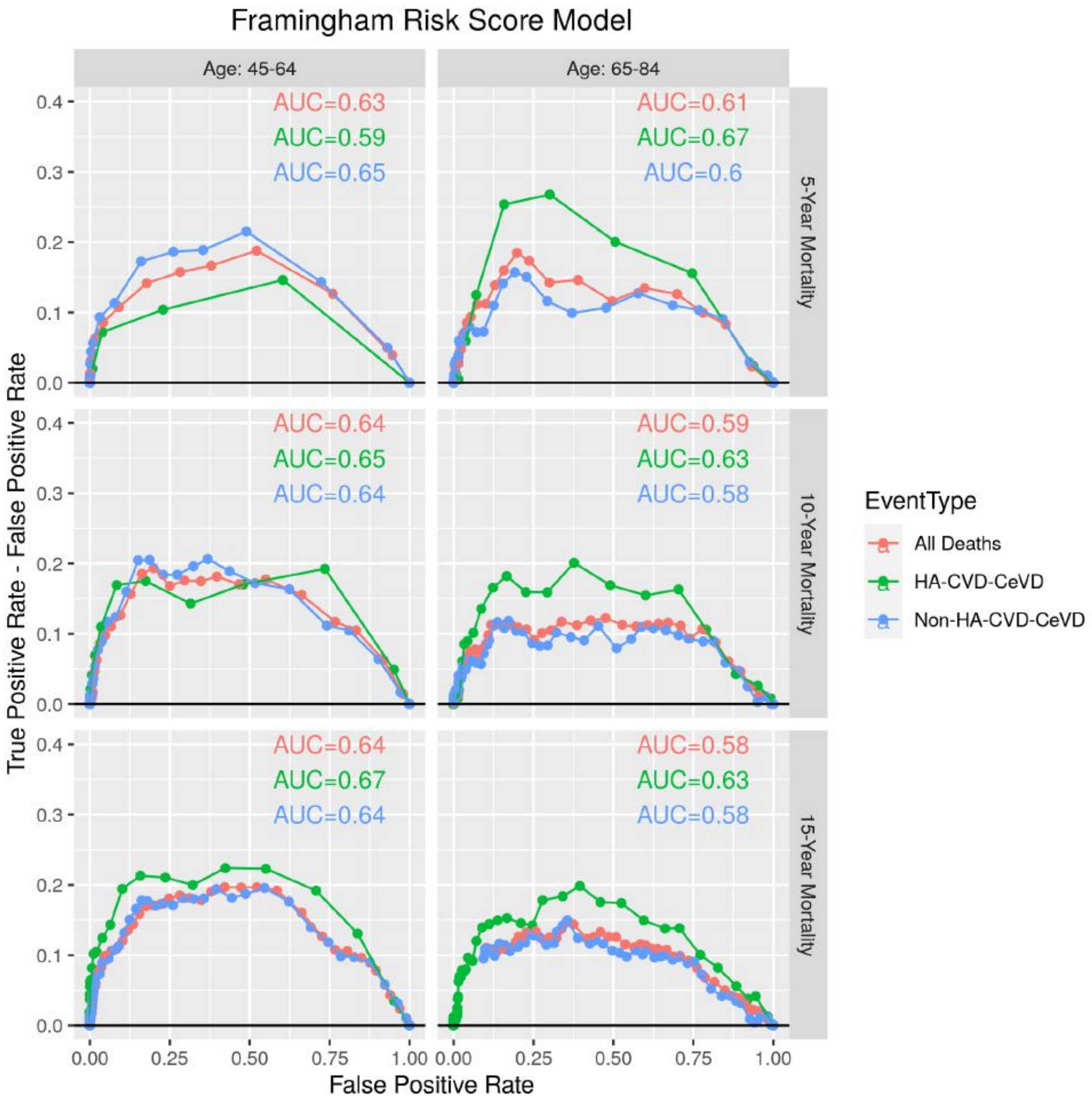


Figure 18: ROC curves for the model that used the FRS value as covariates in the linear predictor, stratified by age group and the number of years the survival outcome was predicted since participant starting the survey.

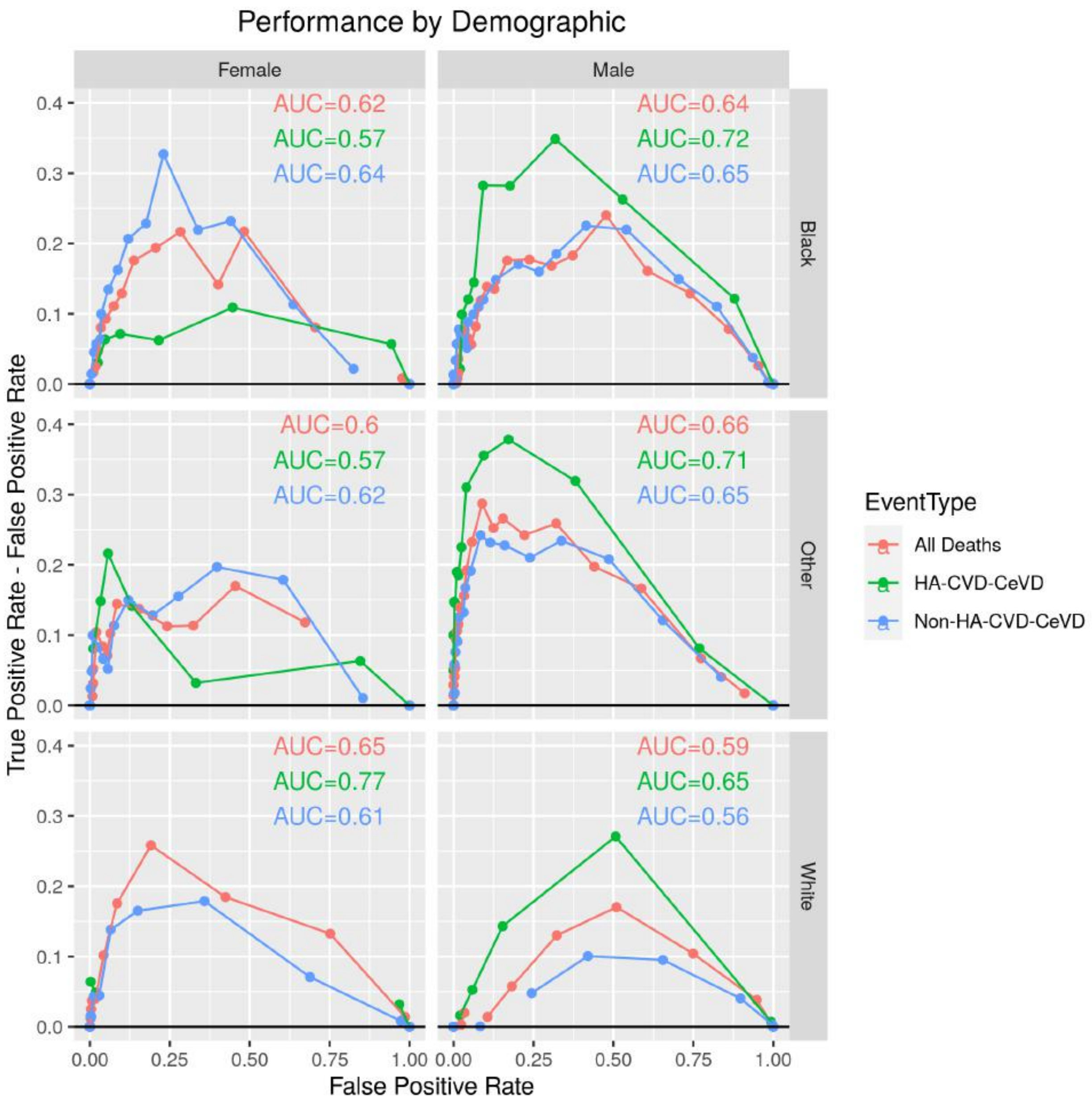


Figure 19: ROC curves stratified by the different demographic groups used in this research. The point and line colours represent the different event types that were used to predict on.

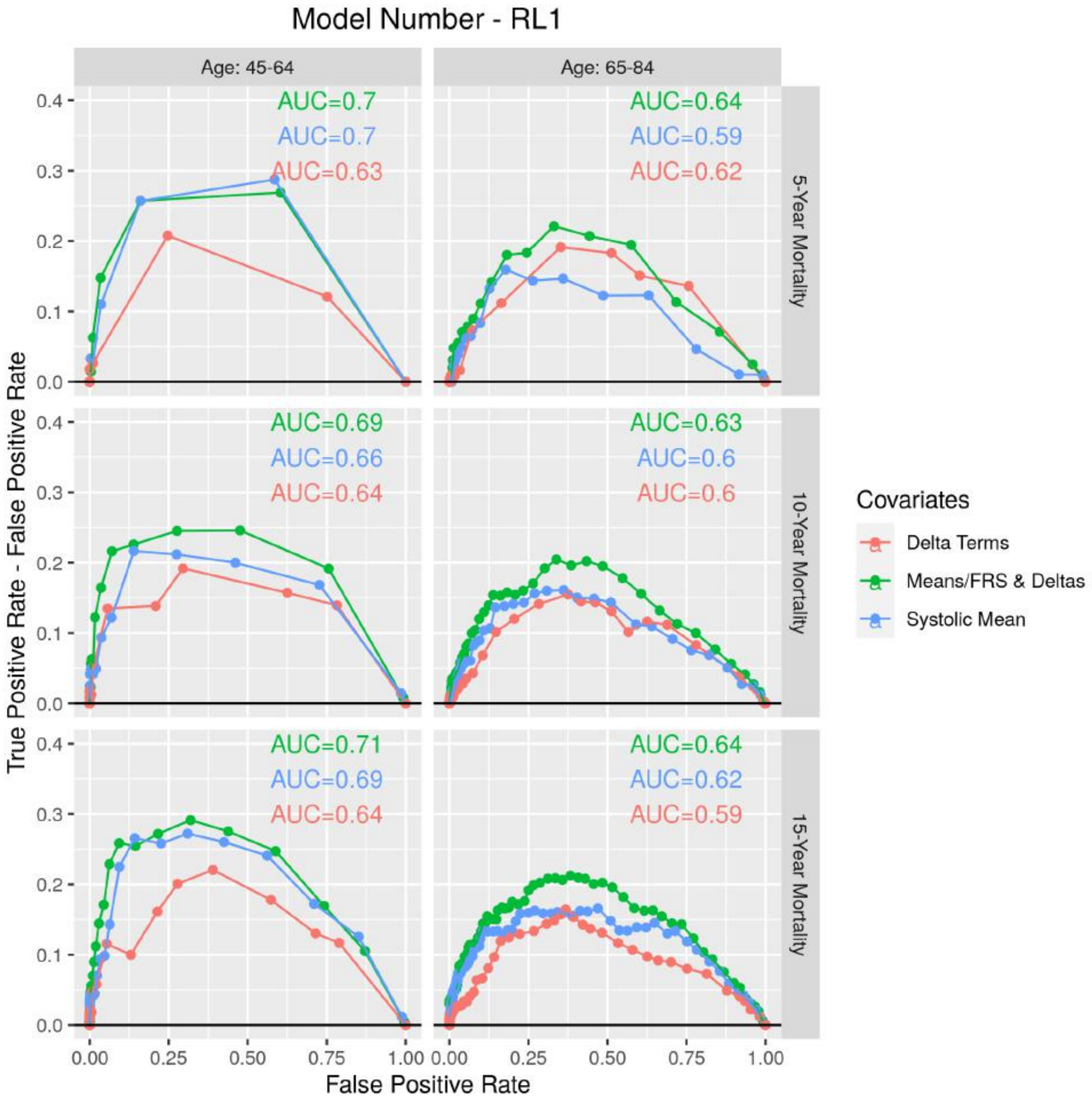


Figure 20: ROC curves for the mean systolic and diastolic model, looking specifically at CVD and heart attack-related deaths, stratified by age group and the number of years the survival outcome was predicted since participant starting the survey. The colour of the points and lines represents the different linear predictor covariate models possible.

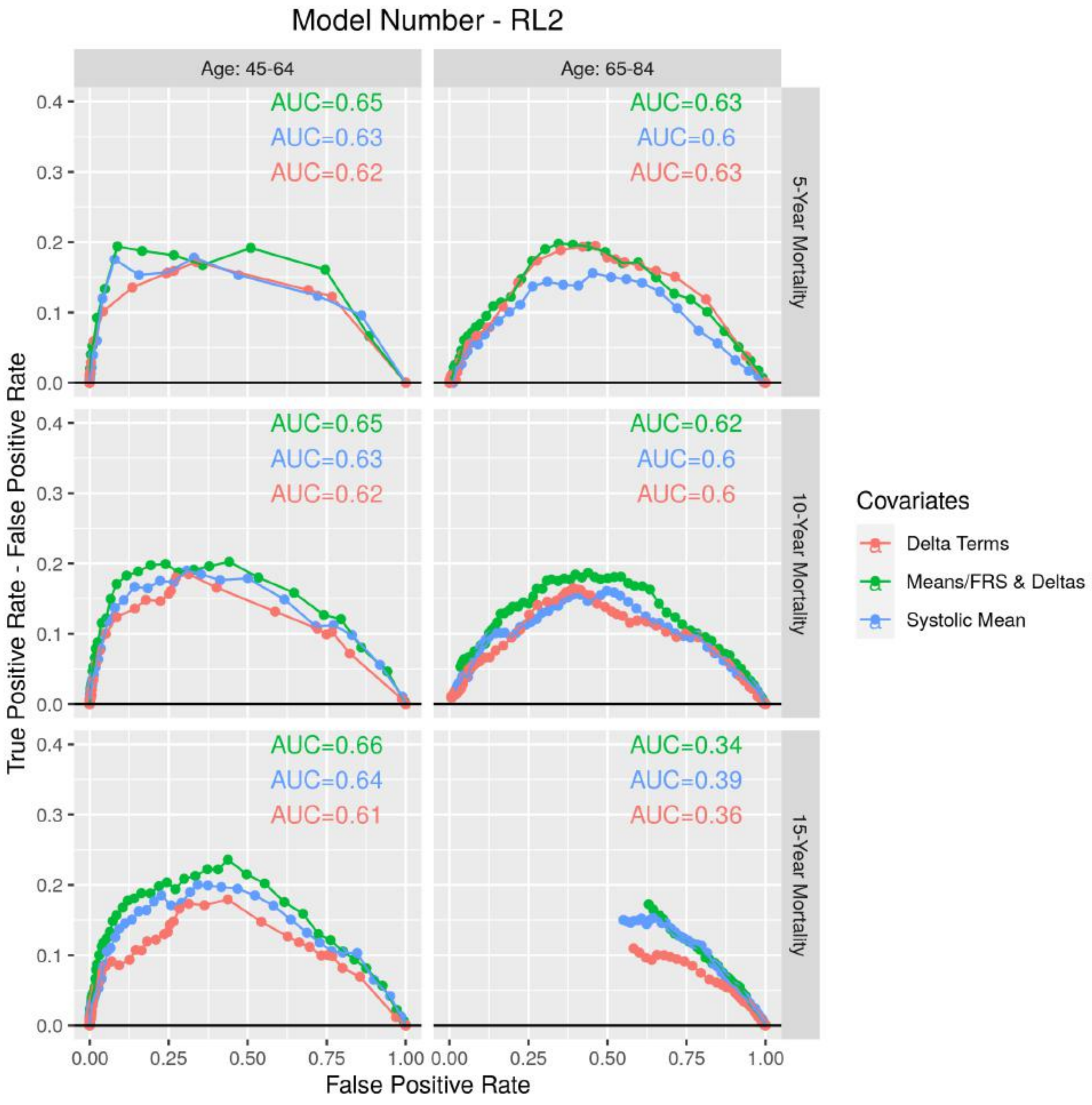


Figure 21: ROC curves for the mean systolic and diastolic model, looking at all-cause deaths, stratified by age group and the number of years the survival outcome was predicted since participant starting the survey. The colour of the points and lines represents the different linear predictor covariate models possible.

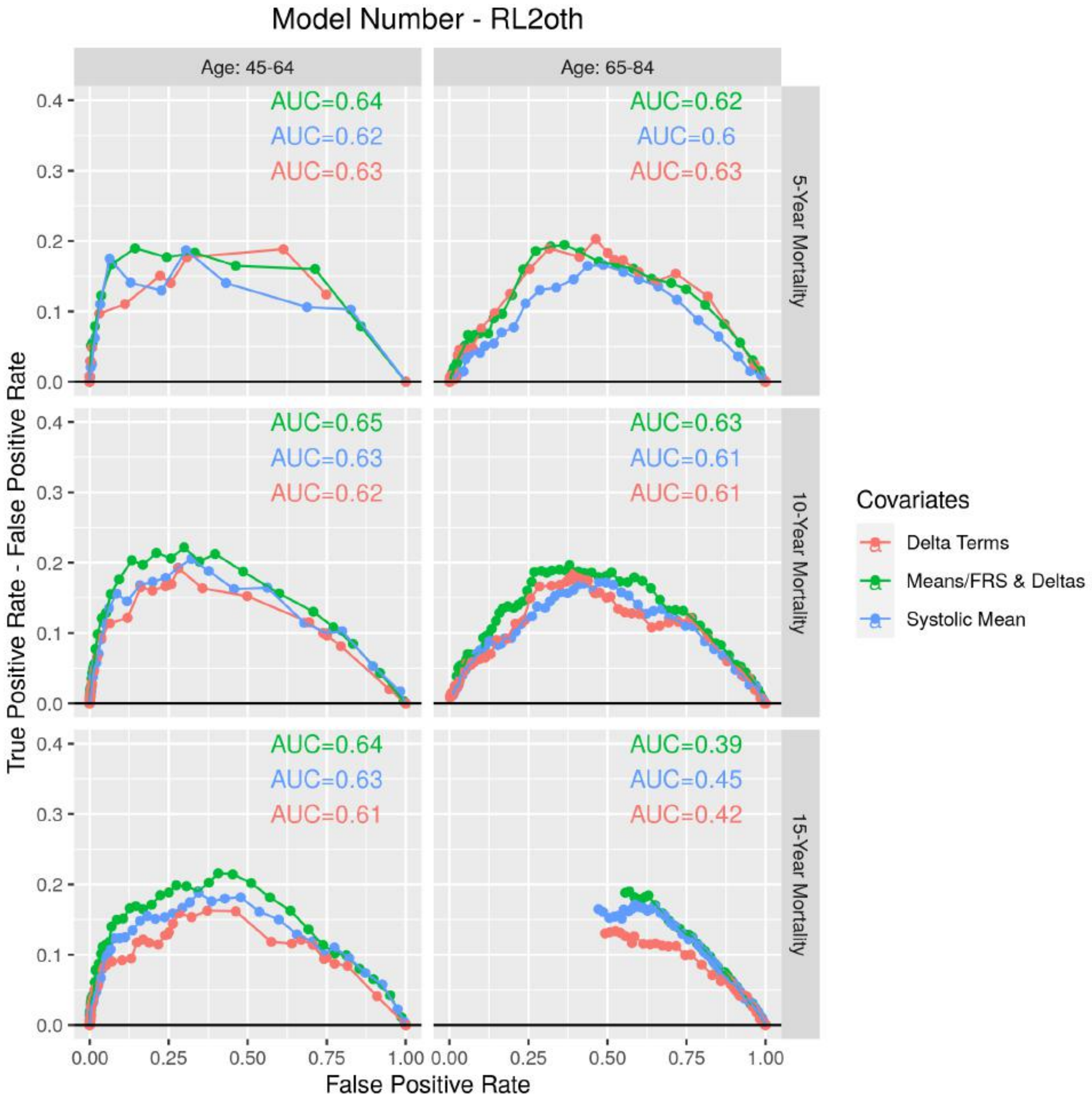


Figure 22: ROC curves for the mean systolic and diastolic model, looking non-CVD and heart attack-related deaths, stratified by age group and the number of years the survival outcome was predicted since participant starting the survey. The colour of the points and lines represents the different linear predictor covariate models possible.

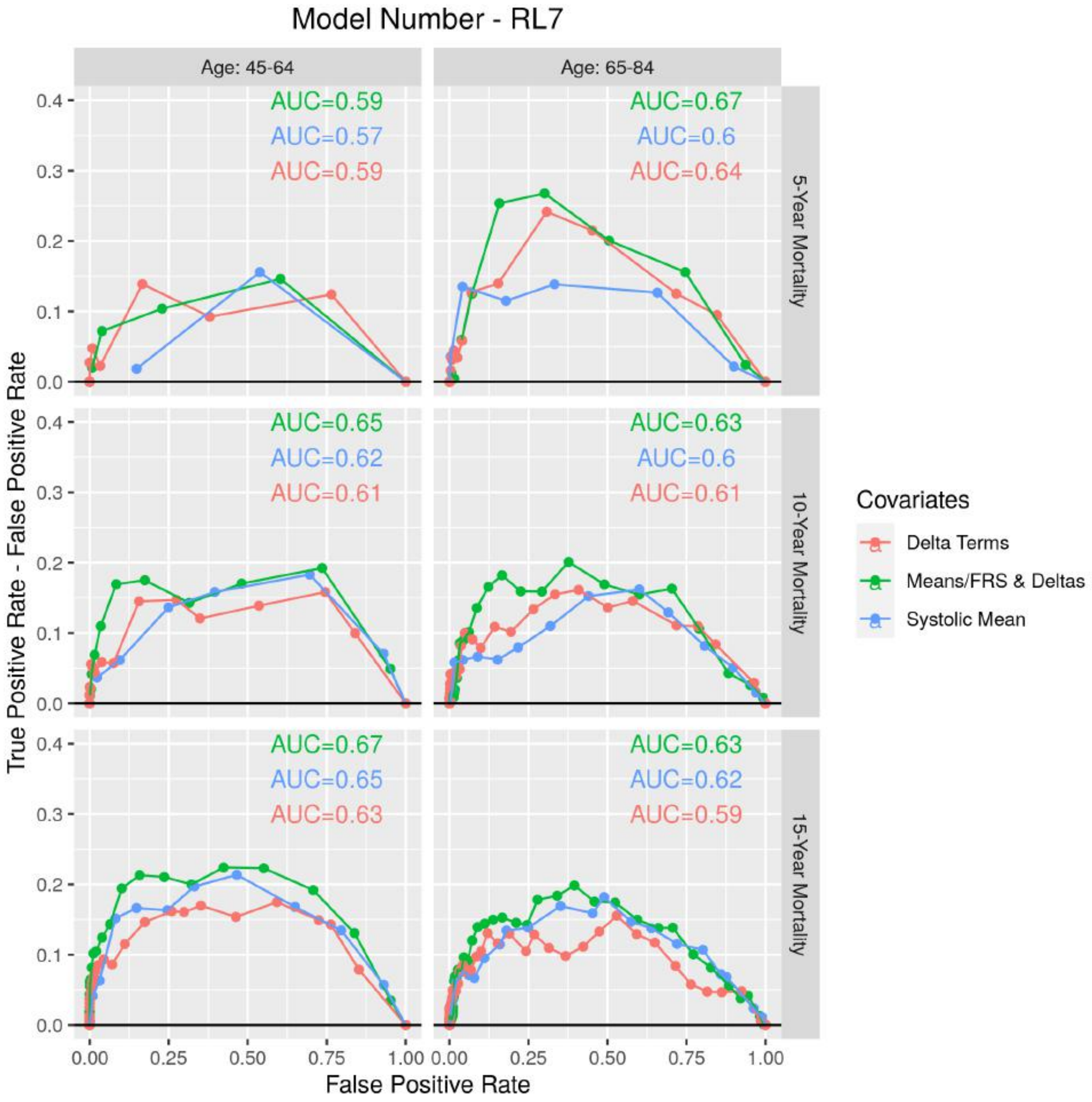


Figure 23: ROC curves for for the FRS-based model, looking specifically at CVD and heart attack-related deaths, stratified by age group and the number of years the survival outcome was predicted since participant starting the survey. The colour of the points and lines represents the different linear predictor covariate models possible.

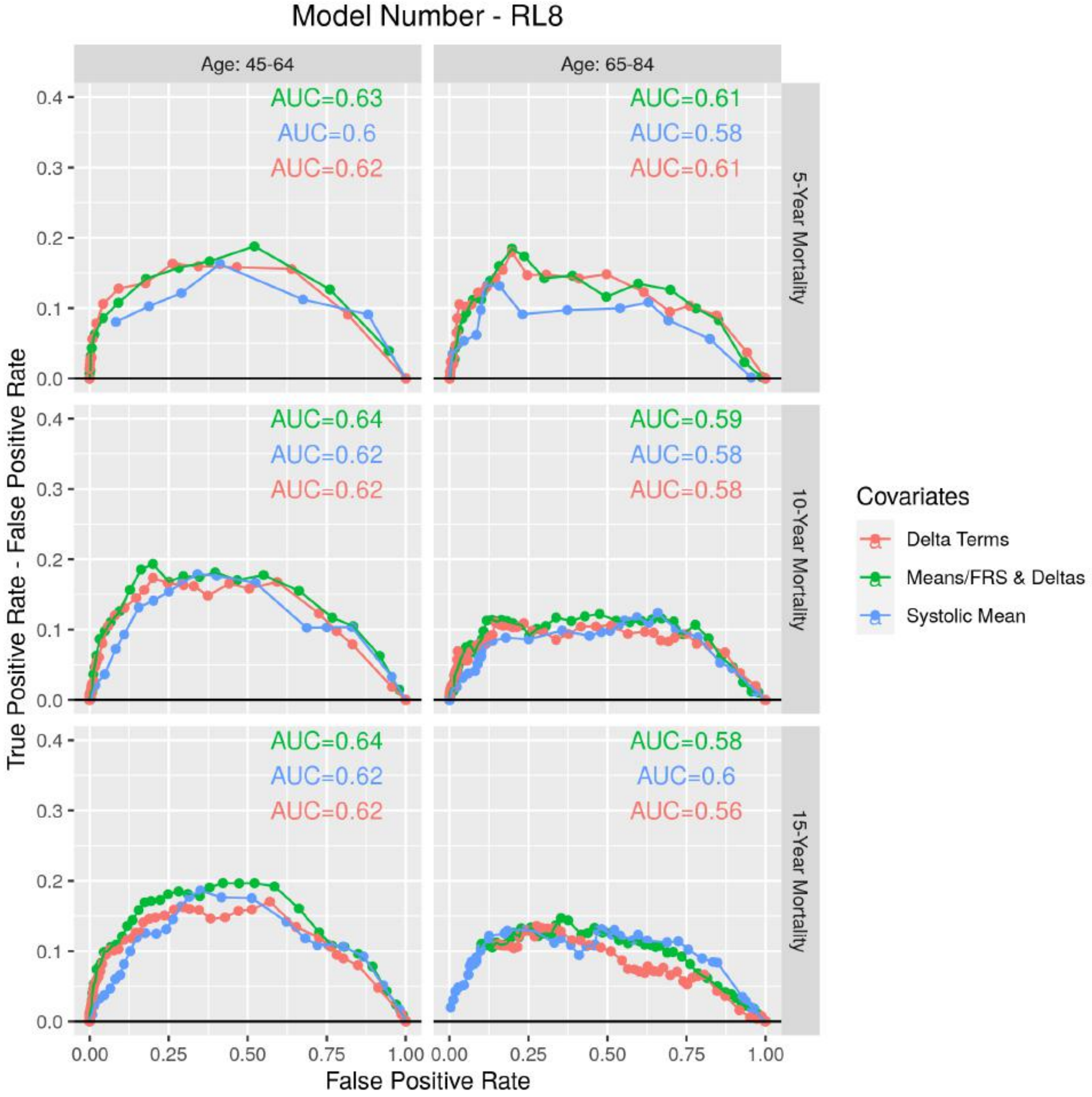


Figure 24: ROC curves for for the FRS-based model, looking at all-cause deaths, stratified by age group and the number of years the survival outcome was predicted since participant starting the survey. The colour of the points and lines represents the different linear predictor covariate models possible.

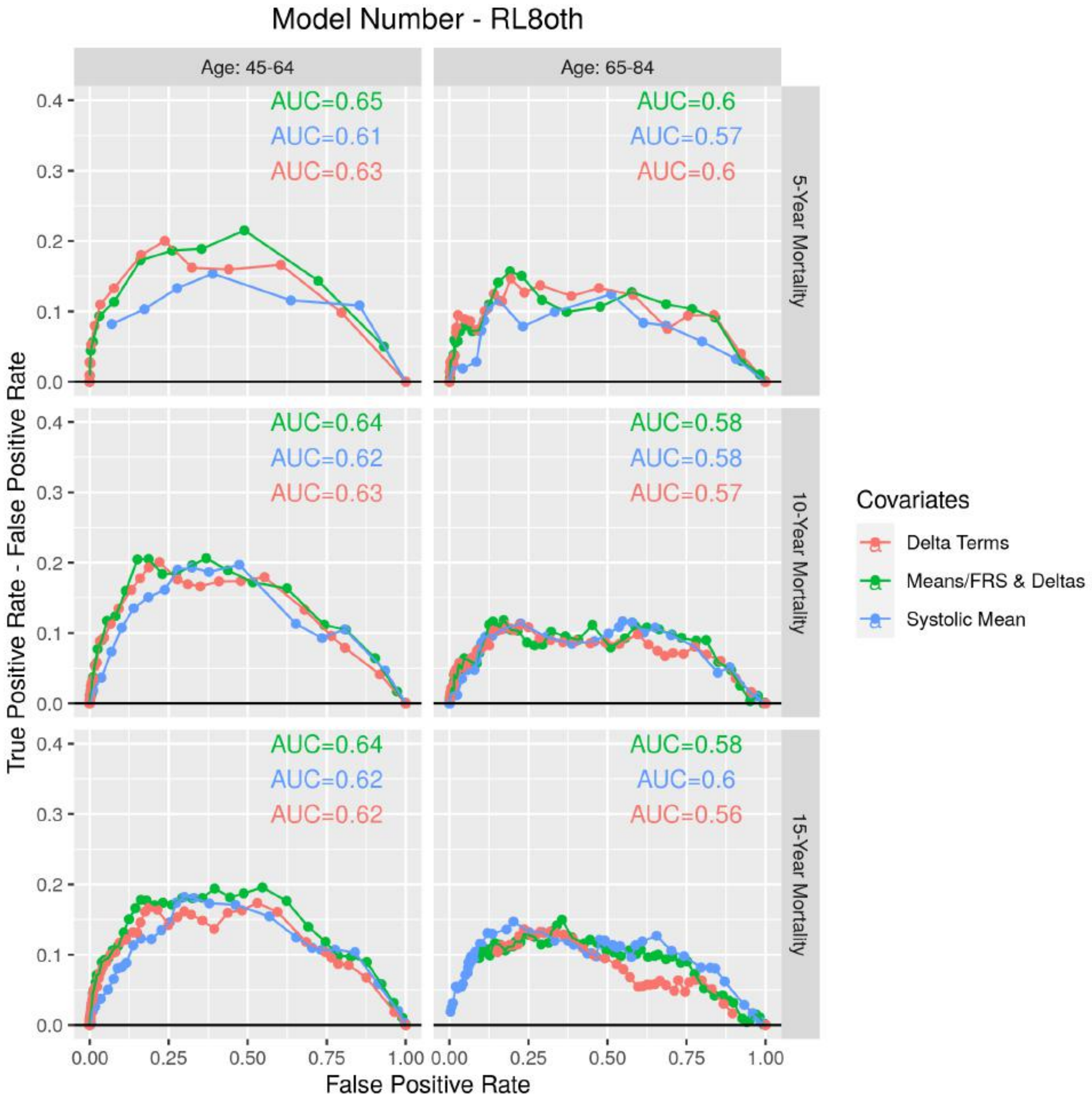


Figure 25: ROC curves for for the FRS-based model, looking non-CVD and heart attack-related deaths, stratified by age group and the number of years the survival outcome was predicted since participant starting the survey. The colour of the points and lines represents the different linear predictor covariate models possible.

Table 14: Frequency table of the population aged 45-64 for the N-year survival outcomes as separated by demographic group and mortality cause.

| Year | EventType | Ethnicity | Gender | Deaths |
|---|---|---|---|---|
| 5-Year Mortality | CVD | Black | Female | 15 |
| 5-Year Mortality | All Deaths | Black | Female | 44 |
| 5-Year Mortality | CVD | White | Female | 9 |
| 5-Year Mortality | All Deaths | White | Female | 33 |
| 5-Year Mortality | CVD | Mexican | Female | 9 |
| 5-Year Mortality | All Deaths | Mexican | Female | 24 |
| 5-Year Mortality | CVD | Black | Male | 6 |
| 5-Year Mortality | All Deaths | Black | Male | 25 |
| 5-Year Mortality | CVD | White | Male | 7 |
| 5-Year Mortality | All Deaths | White | Male | 24 |
| 5-Year Mortality | CVD | Mexican | Male | 6 |
| 5-Year Mortality | All Deaths | Mexican | Male | 21 |
| 10-Year Mortality | CVD | Black | Female | 23 |
| 10-Year Mortality | All Deaths | Black | Female | 87 |
| 10-Year Mortality | CVD | White | Female | 21 |
| 10-Year Mortality | All Deaths | White | Female | 67 |
| 10-Year Mortality | CVD | Mexican | Female | 19 |
| 10-Year Mortality | All Deaths | Mexican | Female | 63 |
| 10-Year Mortality | CVD | Black | Male | 17 |
| 10-Year Mortality | All Deaths | Black | Male | 67 |
| 10-Year Mortality | CVD | White | Male | 13 |
| 10-Year Mortality | All Deaths | White | Male | 62 |
| 10-Year Mortality | CVD | Mexican | Male | 10 |
| 10-Year Mortality | All Deaths | Mexican | Male | 44 |
| 15-Year Mortality | CVD | Black | Female | 37 |
| 15-Year Mortality | All Deaths | Black | Female | 132 |
| 15-Year Mortality | CVD | White | Female | 44 |
| 15-Year Mortality | All Deaths | White | Female | 139 |
| 15-Year Mortality | CVD | Mexican | Female | 35 |
| 15-Year Mortality | All Deaths | Mexican | Female | 105 |
| 15-Year Mortality | CVD | Black | Male | 26 |
| 15-Year Mortality | All Deaths | Black | Male | 113 |
| 15-Year Mortality | CVD | White | Male | 29 |
| 15-Year Mortality | All Deaths | White | Male | 117 |
| 15-Year Mortality | CVD | Mexican | Male | 18 |
| 15-Year Mortality | All Deaths | Mexican | Male | 67 |

survival outcome in the population. In the work presented in this article, $\Delta$ is the absolute value of the differences in the means of the blood pressure measurements at the clinic and at home, respectively, for both diastolic and systolic blood pressure. By 'directionality', we refer to whether the difference between the clinic and home mean measurements are positive or negative. Figure 26 shows the clinic-home directionalities, split by demographic group, indicating no significant difference between the different demographic groups. There is a general trend that the directionality for systolic and diastolic blood pressure is more likely to be the same than opposite.

In order to explore whether the directionality of the clinic-home measurements influences survival outcome, we will use a combination of Kaplan-Meier curves and Cox's proportional hazards regression. The latter will implement a simple Maximum Likelihood Estimation (MLE) method based on summary statistics of the

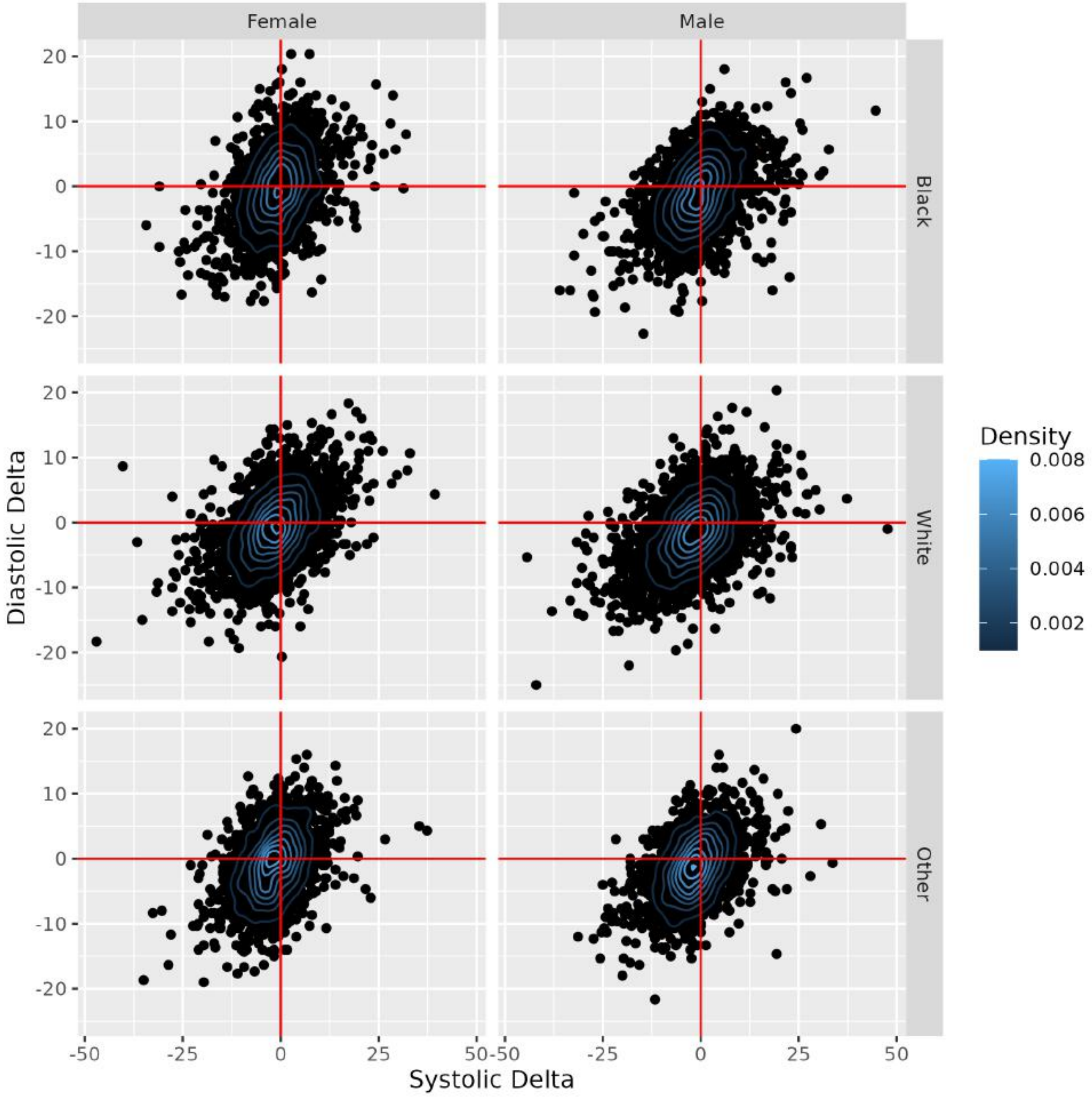


Figure 26: The range of the non-absolute Δ values in the systolic and diastolic blood pressure measurements, split by demographic group. This reflects the differences between the average measurements at the clinic and at home.

Table 15: Frequency table of the population aged 65-84 for the N-year survival outcomes as separated by demographic group and mortality cause.

| Year | EventType | Ethnicity | Gender | Deaths |
|---|---|---|---|---|
| 5-Year Mortality | CVD | Black | Female | 35 |
| 5-Year Mortality | All Deaths | Black | Female | 87 |
| 5-Year Mortality | CVD | White | Female | 92 |
| 5-Year Mortality | All Deaths | White | Female | 253 |
| 5-Year Mortality | CVD | Mexican | Female | 15 |
| 5-Year Mortality | All Deaths | Mexican | Female | 45 |
| 5-Year Mortality | CVD | Black | Male | 19 |
| 5-Year Mortality | All Deaths | Black | Male | 49 |
| 5-Year Mortality | CVD | White | Male | 55 |
| 5-Year Mortality | All Deaths | White | Male | 138 |
| 5-Year Mortality | CVD | Mexican | Male | 10 |
| 5-Year Mortality | All Deaths | Mexican | Male | 29 |
| 10-Year Mortality | CVD | Black | Female | 56 |
| 10-Year Mortality | All Deaths | Black | Female | 153 |
| 10-Year Mortality | CVD | White | Female | 187 |
| 10-Year Mortality | All Deaths | White | Female | 501 |
| 10-Year Mortality | CVD | Mexican | Female | 37 |
| 10-Year Mortality | All Deaths | Mexican | Female | 105 |
| 10-Year Mortality | CVD | Black | Male | 50 |
| 10-Year Mortality | All Deaths | Black | Male | 121 |
| 10-Year Mortality | CVD | White | Male | 144 |
| 10-Year Mortality | All Deaths | White | Male | 357 |
| 10-Year Mortality | CVD | Mexican | Male | 44 |
| 10-Year Mortality | All Deaths | Mexican | Male | 91 |
| 15-Year Mortality | CVD | Black | Female | 79 |
| 15-Year Mortality | All Deaths | Black | Female | 231 |
| 15-Year Mortality | CVD | White | Female | 251 |
| 15-Year Mortality | All Deaths | White | Female | 694 |
| 15-Year Mortality | CVD | Mexican | Female | 56 |
| 15-Year Mortality | All Deaths | Mexican | Female | 170 |
| 15-Year Mortality | CVD | Black | Male | 76 |
| 15-Year Mortality | All Deaths | Black | Male | 185 |
| 15-Year Mortality | CVD | White | Male | 225 |
| 15-Year Mortality | All Deaths | White | Male | 589 |
| 15-Year Mortality | CVD | Mexican | Male | 57 |
| 15-Year Mortality | All Deaths | Mexican | Male | 139 |

Bayesian posterior blood pressure values, not the Bayesian-HMC method applied elsewhere in this article. The Kaplan-Meier curve is a plot of the change in survival probability of a population in time since the start of a survey/census. The survival distribution is calculated using

$$\widehat{S}(t) = \prod_{t_j \leq t} \left(1 - \frac{d_j}{r_j}\right), \tag{27}$$

for $d_j$ the number of individuals who die within the time interval $t_j$ and $r_j$ the population that are alive (at risk of death) and not censored. Greenwood's formula is used to calculate the variance of the Kaplan-Meier estimation

$$\hat{\sigma}(t)^2 = \hat{S}(t)^2 \sum_{t_j \leq t} \left( \frac{d_j}{r_j(r_j - d_j)} \right). \tag{28}$$

The 100(1-$\alpha$)% confidence intervals of the Kaplan-Meier estimate are assumed to be normally distributed

$$\hat{S}(t) \pm z_{1-\alpha/2}\hat{\sigma}(t). \tag{29}$$

Figures 27 and 28 show the Kaplan-Meier estimates for the full NHANES population for CVD mortality, split by demographic group, for the age range 45-64 and 65-84, respectively. The survival probability of the older population decreases faster in time than the middle-aged (45-64) population.

By splitting the populations into the respective regions of the $\Delta$ directionality, we can use the different Kaplan-Meier plots to try to identify differences in the survival outcomes. Figures 29 and 30 show the Kaplan-Meier estimates for the full NHANES population for CVD mortality, split by $\Delta$ directionality and demographic group, for the age range 45-64 and 65-84, respectively. With the broad confidence intervals, all of the different $\Delta$ directionality regions overlap, for all demographic groups and both age range groups (where $\hat{S}(t) \neq 1$).

We further quantify this insignificant relationship between $\Delta$ directionality and survival outcome via the use of a Cox's Proportional Hazards (CPH) model. Via the use of the 'coxph' function in the 'survival' R package, we fit (using MLE) a CPH model. The covariates used in the model are $\Delta$ directionality region, gender, ethnicity and age. Table 16 shows the summary of the model fit, which reflects that being in different $\Delta$ directionality regions has a non-statistically significant influence on survival outcomes. As shown in the remainder of this article, ethnicity, gender and age are shown to have statistically significant effects on survival outcome.

Table 16: Parameters for distribution of blood pressure, for the full population

| covariate | coef | exp(coef) | se(coef) | z | Pr(>\|z\|) |
|---|---|---|---|---|---|
| DeltaRegionSys. -ve, Dys. +ve | 0.047 | 1.048 | 0.074 | 0.634 | 0.526 |
| DeltaRegionSys. +ve, Dys. -ve | -0.002 | 0.998 | 0.069 | -0.031 | 0.975 |
| DeltaRegionSys. +ve, Dys. +ve | -0.056 | 0.946 | 0.063 | -0.881 | 0.378 |
| GenderMale | -0.413 | 0.662 | 0.050 | -8.282 | 0.000 |
| EthnicityWhite | -0.248 | 0.780 | 0.061 | -4.043 | 0.000 |
| EthnicityMexican | -0.170 | 0.844 | 0.075 | -2.272 | 0.023 |
| age | 0.100 | 1.105 | 0.002 | 50.573 | 0.000 |

Finally, we wish to confirm that the perfomance of the model does not depend on the directionality of $\Delta$. Figure 31 plots the AUC values of 10-year CVD mortality for the all-covariate mean blood pressure-based model trained on the full NHANES population, split by the two age-ranges (45-64 and 65-84) and demographic groups. There is no clear trend between mode performance for the different regions of $\Delta$ directionality.

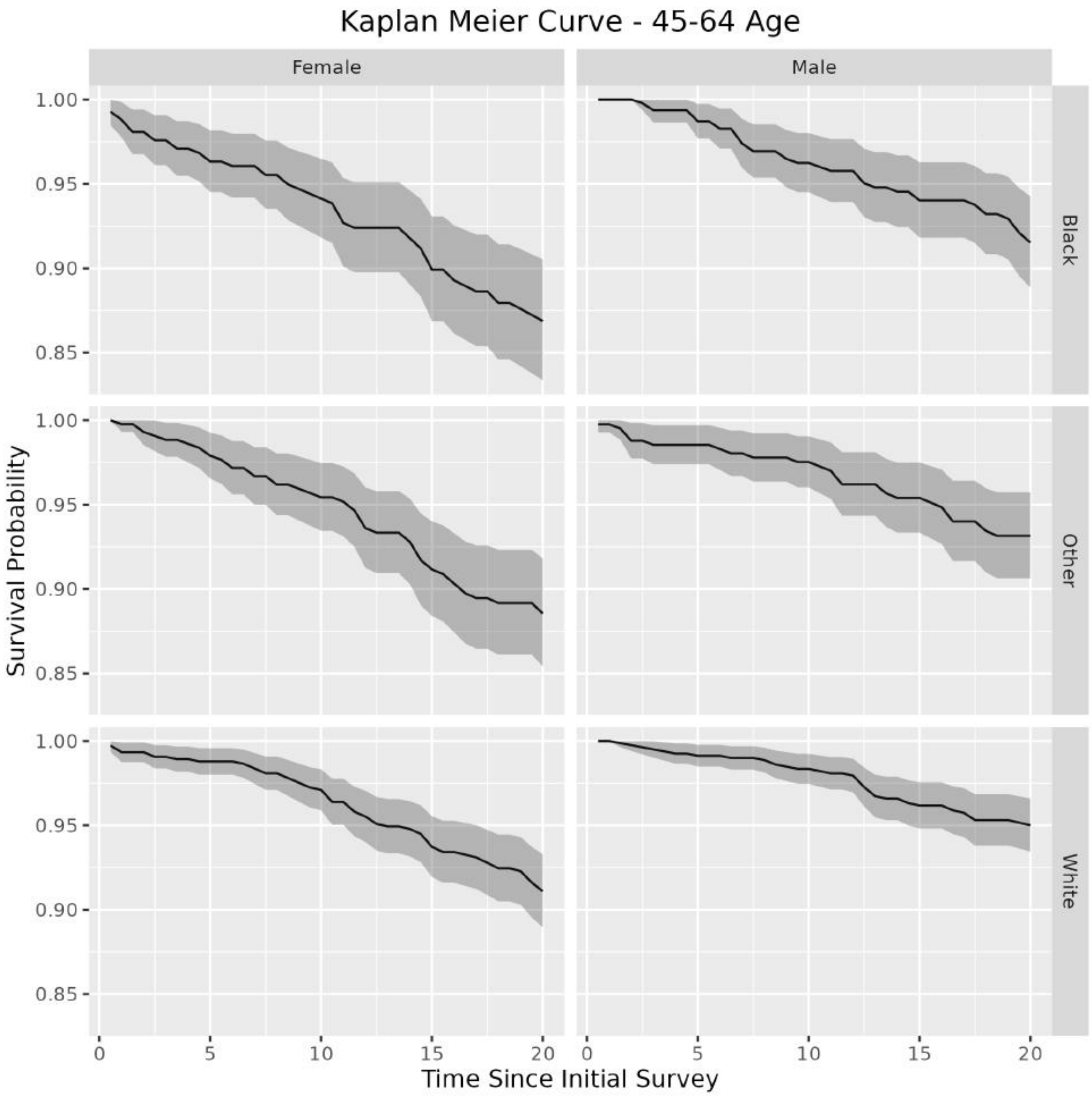


Figure 27: Kaplan-Meier plots of the full-population for CVD mortality, for ages between 45-64, split by demographic group.

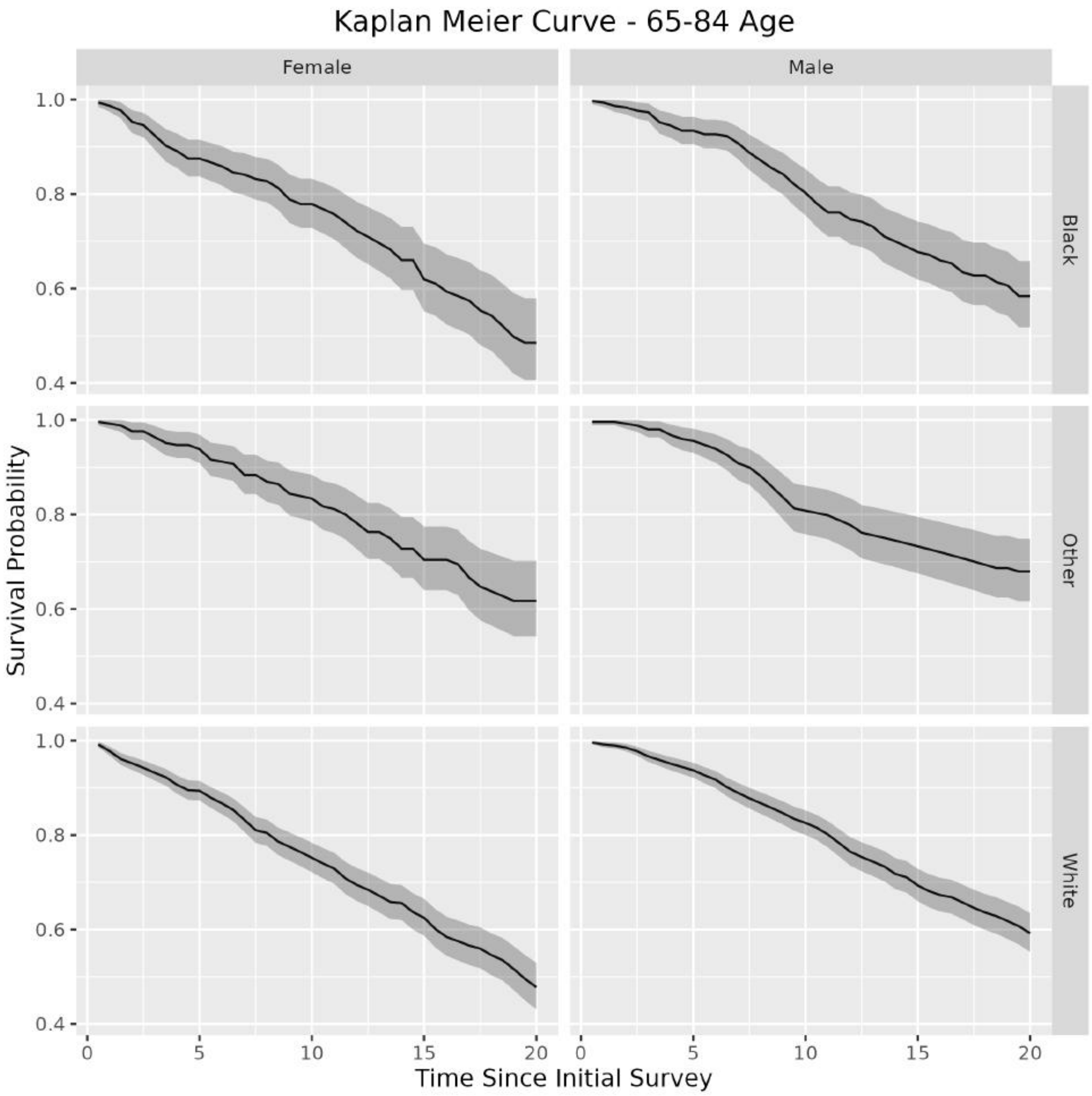


Figure 28: Kaplan-Meier plots of the full-population for CVD mortality, for ages between 65-84, split by demographic group.

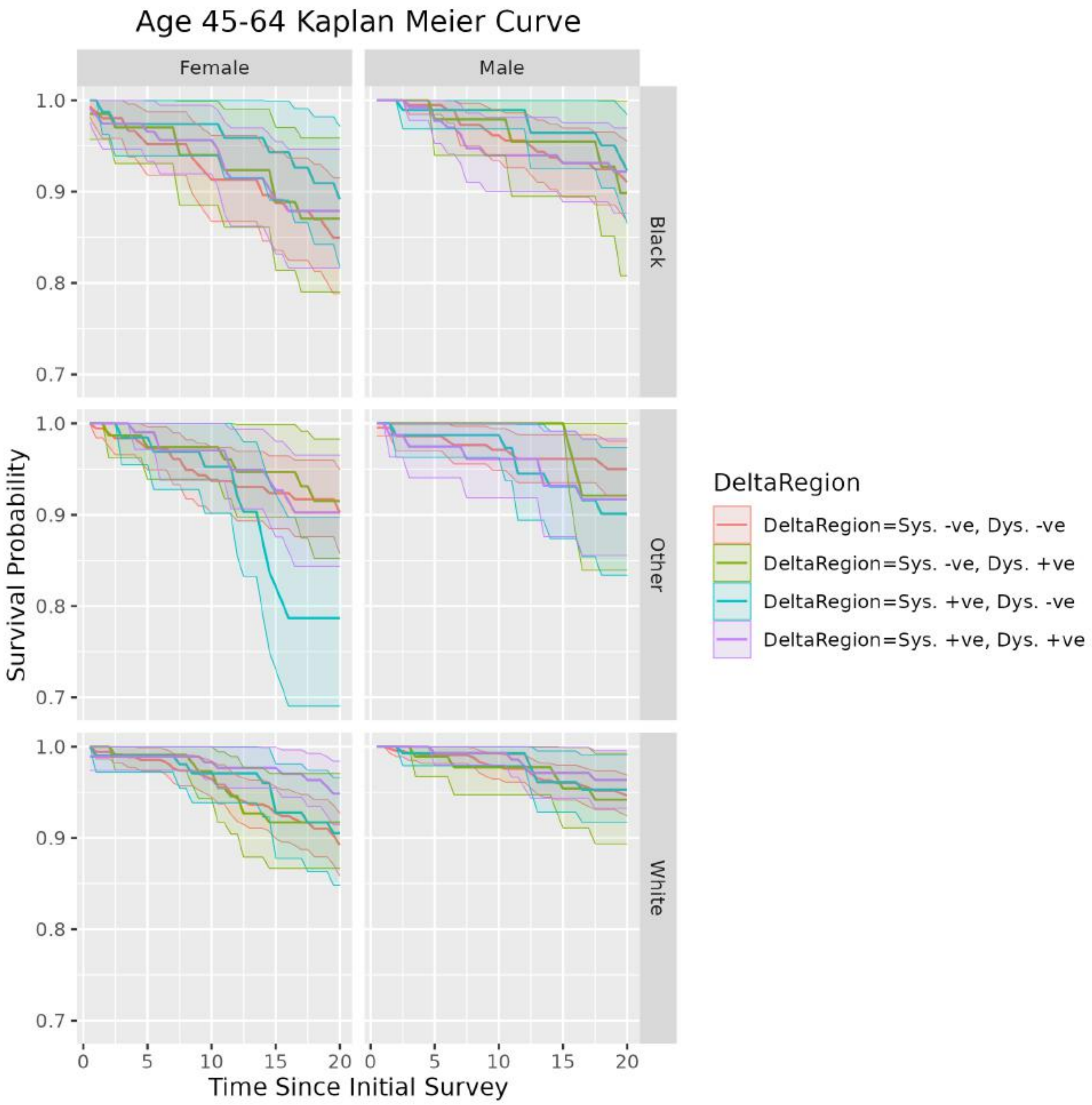


Figure 29: Kaplan-Meier plots of the full-population for CVD mortality, for ages between 45-64, split by demographic group and region in systolic-diastolic $\Delta$ space.

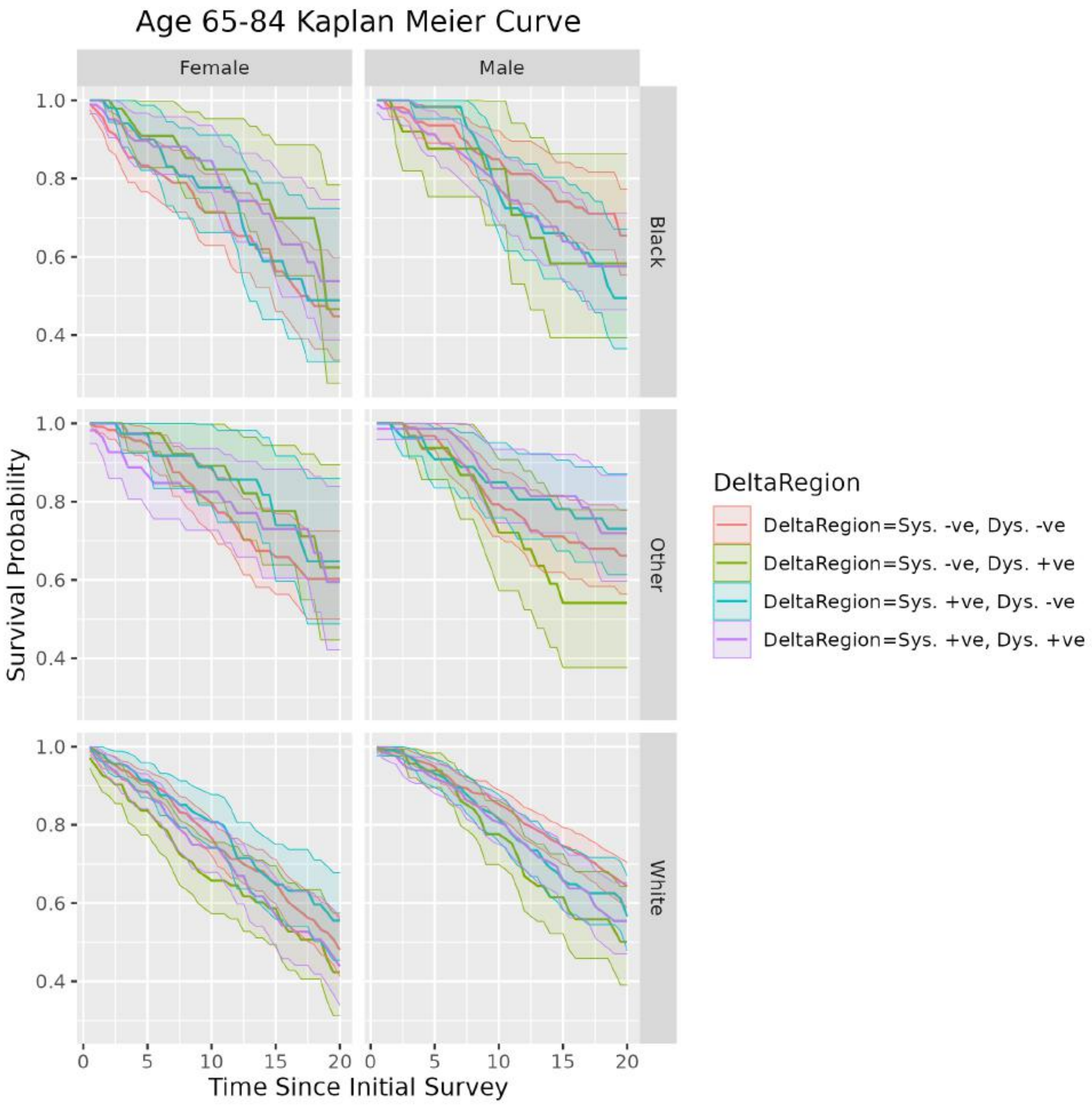


Figure 30: Kaplan-Meier plots of the full-population for CVD mortality, for ages between 65-84, split by demographic group and region in systolic-diastolic $\Delta$ space.

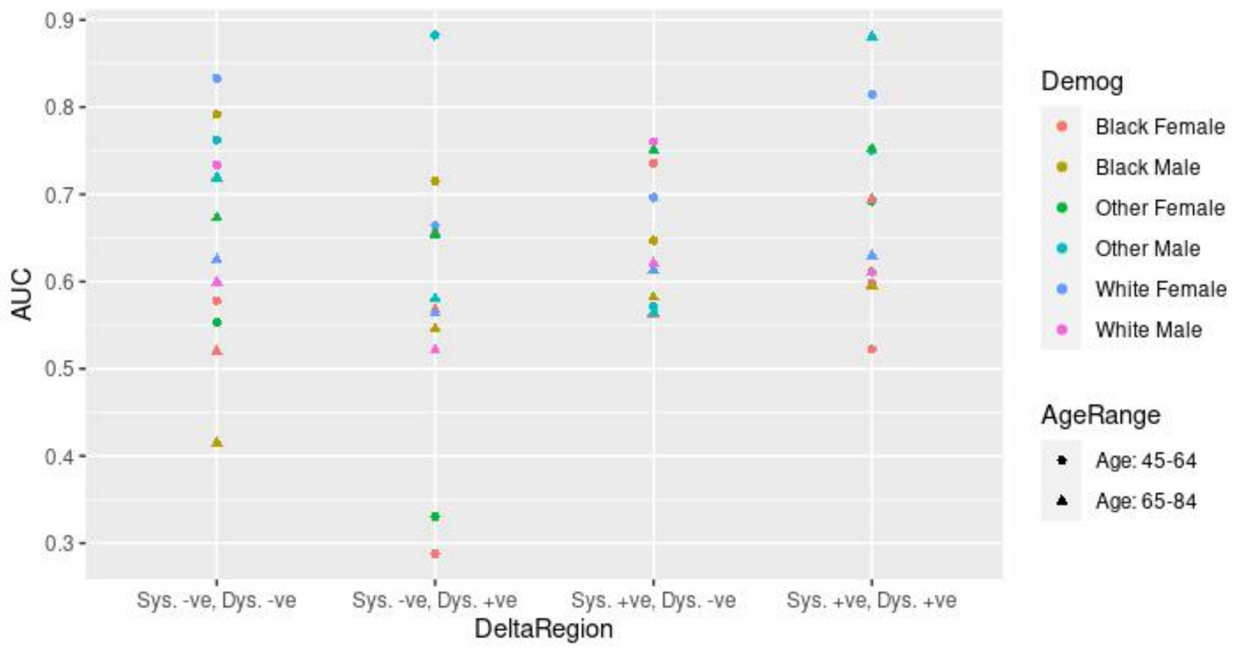


Figure 31: AUC values of the full-population, all covariate model with the mean blood pressure model, based on CVD 10-year mortality.
